# Can we predict the severe course of COVID-19 – a systematic review and meta-analysis of indicators of clinical outcome?

**DOI:** 10.1101/2020.11.09.20228858

**Authors:** Stephan Katzenschlager, Alexandra J. Zimmer, Claudius Gottschalk, Juergen Grafeneder, Alexander Seitel, Lena Maier-Hein, Andrea Benedetti, Jan Larmann, Markus A. Weigand, Sean McGrath, Claudia M. Denkinger

**Affiliations:** Department of Anesthesiology, Heidelberg University Hospital, Heidelberg, Germany; Departments of Epidemiology, Biostatistics and Occupational Health, McGill University, Montreal, Canada; Division of Tropical Medicine, Center for Infectious Diseases, Heidelberg University Hospital, Heidelberg, Germany; Department of Clinical Pharmacology, Medical University of Vienna, Vienna, Austria; Division of Computer Assisted Medical Interventions, German Cancer Research Center (DKFZ), Heidelberg, Germany; Department of Biostatistics, Harvard T.H. Chan School of Public Health, Boston, USA; German Center for Infection Research (DZIF), partner site Heidelberg

**Keywords:** Covid-19, outcome prediction, meta-analysis, risk factors, mortality

## Abstract

**Background:** COVID-19 has been reported in over 40million people globally with variable clinical outcomes. In this systematic review and meta-analysis, we assessed demographic, laboratory and clinical indicators as predictors for severe courses of COVID-19.

**Methods:** We systematically searched multiple databases (PubMed, Web of Science Core Collection, MedRvix and bioRvix) for publications from December 2019 to May 31^st^ 2020. Random-effects meta-analyses were used to calculate pooled odds ratios and differences of medians between (1) patients admitted to ICU versus non-ICU patients and (2) patients who died versus those who survived. We adapted an existing Cochrane risk-of-bias assessment tool for outcome studies.

**Results:** Of 6,702 unique citations, we included 88 articles with 69,762 patients. There was concern for bias across all articles included. Age was strongly associated with mortality with a difference of medians (DoM) of 13.15 years (95% confidence interval (CI) 11.37 to 14.94) between those who died and those who survived. We found a clinically relevant difference between non-survivors and survivors for C-reactive protein (CRP; DoM 69.10, CI 50.43 to 87.77), lactate dehydrogenase (LDH; DoM 189.49, CI 155.00 to 223.98), cardiac troponin I (cTnI; DoM 21.88, CI 9.78 to 33.99) and D-Dimer (DoM 1.29mg/L, CI 0.9 - 1.69). Furthermore, cerebrovascular disease was the co-morbidity most strongly associated with mortality (Odds Ratio 3.45, CI 2.42 to 4.91) and ICU admission (Odds Ratio 5.88, CI 2.35 to 14.73).

**Discussion:** This comprehensive meta-analysis found age, cerebrovascular disease, CRP, LDH and cTnI to be the most important risk-factors in predicting severe COVID-19 outcomes and will inform decision analytical tools to support clinical decision-making.

**Summary:** In this systematic review we meta-analyzed 88 articles for risk factors of ICU admission and mortality in COVID-19. We found age, cerebrovascular disease, CRP, LDH and cTnI are the most important risk-factors for ICU admission or mortality.

## Introduction

Coronavirus disease (COVID-19) was declared a pandemic by the World Health Organization (WHO) on March 11^th^, 2020 [1]. As of October 31^st^ approximately 46 million people were infected with this virus [2]. The outcomes of COVID-19 vary from completely asymptomatic to hospitalization, ICU admission and death [3, 4].

Several studies aimed to identify possible risk factors for a severe outcome. Studies investigated demographic risk factors and found advanced age to be the strongest predictor of a severe course [5-7]. However, age alone does not explain the variability in the severity of disease with sufficient granularity [8]. Symptoms on presentation associated with severe disease include dyspnea, fever, cough, and fatigue [6, 9, 10]. Several co-morbidities have been identified as risk factors, including cardiovascular disease, obesity, chronic respiratory disease, diabetes, cerebrovascular disease, chronic renal failure and cancer [7, 11-16]. The effect of other co-morbidities on disease outcome remain less clear: e.g. hypertension being associated with a decreased risk [7, 17] for death in some and an increased risk [18] in other publications. Similarly, data on past and current smoking are inconsistent in respect of the association with disease severity [19-23]. Biomarkers predicting severe disease include different markers of inflammation and acute phase reaction (e.g. CRP, procalcitonin (PCT), white blood cells (WBC), lymphopenia, interleukin 6 (IL-6)) [24, 25]. Increased D-Dimer levels, as a marker for coagulation and thrombosis, were found to be elevated in non-survivors, whereas other coagulation markers failed to show statistical and clinical difference [13, 26-28]. Markers indicating cardiac damage, such as cardiac troponin I or T and N terminal pro B type natriuretic peptide (NT-proBNP) were also associated with severe disease and mortality [29].

This systematic review aims, to our knowledge for the first time, to comprehensively evaluate demographic, clinical and laboratory indicators for their association with severe COVID-19 and death.

## Methods

This trial was registered at PROSPERO on April 4^th^, 2020 (Registration number: CRD42020177154). The PRISMA checklist is provided in the supplementary file S2.

### Eligibility criteria

Studies eligible for inclusion provided data on demographic, clinical and/or laboratory risk factors for the following outcomes: hospitalization, intubation, ICU admission, and/or death. Laboratory values and vital parameters taken at hospital admission were considered. Cross-sectional studies, cohort studies, randomized and non-randomized controlled trials were included. No specific restrictions were placed in terms of demographic and clinical characteristics of the population being studied. The search was conducted on July 29^th^, the search date was set from December 1^st^ 2019 to May 31^st^ 2020.

### Search strategy

Medline [PubMed] and Web of Science Core Collection as well as preprint databases (bioRxiv and medRxiv) were searched. The exact search terms were developed with an experienced medical librarian (GG) using combinations of subject headings (when applicable) and text-words for the concepts without language restrictions. The full search strategy used for PubMed is presented in the supplementary file S1. The results of the search term were imported into the bibliography manager Zotero (Version 5.0.92) for further processing.

### Study screening and data extraction

Study selection was done by three authors (SK, CG and JG) initially in parallel for five randomly selected papers and after alignment in the selection was guaranteed, it was done independently by each of the reviewers. Article title and abstracts were screened for eligibility, followed by a full-text review for those eligible.

A structured electronic data extraction form was developed (AS, LMH, SO, LAS and BP), piloted on five randomly selected papers and then used to extract information from included studies. Six reviewers (SK, CG, SS, SaK, MG and AM) performed data extraction in duplicate for the first five randomly selected papers to ensure alignment and then independently, with concerns being discussed jointly. For continuous indicators we extracted means and standard deviation as well as medians, first quartiles and third quartiles if available. The comprehensive list of data items that were collected is presented in the supplementary file S12. Throughout screening and extraction, disagreements were discussed until consensus was reached, and a senior author (CMD) was consulted when necessary.

Given the concern for reporting of the same patients in different publications [30] leading to a bias in the data, we excluded papers which included patients from the same hospital with an overlapping inclusion date. Furthermore, we excluded data from 23 articles (peer-reviewed and preprint), because the reported laboratory values with the reported units were obviously incorrect (Supplementary file S3), unless we were able to clarify the issue with the authors of the respective paper directly.

### Assessment of study quality

To analyze risk of bias in individual studies, we evaluated the studies using an approach adapted from an existing Cochrane tool by Higgins et al. [31] for systematic reviews that assessed indicators of outcomes. Specifically, we analyzed three areas: 1) case definition and severity definition; 2) patient data availability and exclusions and; 3) selection bias and applicability. We rated the risk of bias in low, intermediate and high risks of bias.

### Statistical analysis

We grouped indicators into binary and continuous indicators across five categories: (1) demographics, (2) symptoms, (3) co-morbidities, (4) laboratory and (5) clinical course/treatment. We analyzed all available indicators between (1) hospitalized and non-hospitalized patients, (2) ICU-admitted patients and non-ICU admitted patients, (3) intubated and non-intubated patients, and (4) patients who died and patients who survived. Most data were available for ICU admission and death. Thus, we focus on these comparison groups in the main paper and present data on hospitalization and intubation in the supplement.

Meta-analyses were only performed when there were at least 4 primary studies reporting adequate summary data. As the continuous indicators were often skewed and were summarized by medians in most primary studies, we meta-analyzed the difference of medians across groups for continuous indicators. Specifically, we pooled the difference of medians in a random effects meta-analysis using the Quantile Estimation (QE) approach proposed by McGrath et al. [32]. In secondary analyses, median value of indicators in each comparison group were pooled using the same approach.

The QE approach estimates the variance of the difference of medians in studies that report the sample median and first and third quartiles of the outcome. When studies report sample means and standard deviations of the outcome, this approach estimates the difference of medians and its variance. Then, the standard inverse-variance approach is applied to obtain a pooled estimate of the population (difference of) medians.

For binary indicators, the pooled odds ratios (OR) and associated 95% confidence intervals (CI) were estimated in a random effects meta-analysis. For both binary and continuous indicators, the restricted maximum likelihood (REML) approach was used to estimate between-study heterogeneity. When REML failed to converge, we used the DerSimmonian and Laird (DL) estimator for continuous indicators. We considered findings as statistically significant if p<0.05.

For all analyses, between-study heterogeneity was assessed by the I^2^ statistic. The presence of publication bias was visually assessed in funnel plots. Analyses were performed in R (version 4.0.2) with package ‘metamedian’ [33] and in Stata (Version 16.1). The code is publicly available on GitHuB (https://github.com/stmcg/covid-ma).

## Results

The search resulted in 6,702 articles, of which 3,733 were excluded because they did not present primary data (e.g. guidelines, recommendations, letter to the editors or correspondences, study protocols, modeling), 792 were case reports, 465 focused on patients younger than 18 years and 381 were systematic reviews. In total, 88 articles were included (Figure 1). The majority of studies (52) were conducted in China, 21 in Europe, 12 in the USA, two in Iran, one in South Korea. Most studies were retrospective cohorts (n=84) and four had a prospective study design. All studies were in English. Data on mortality were reported in 64 studies, data on ICU admission were available in 26 studies (two studies reported both and patients were counted twice). In total, data from 69,762 patients were meta-analyzed, of whom 5,311 died and 57,321 survived and 2,112 provided data on ICU-admission while 5,018 did not require ICU admission.

**Figure 1.**
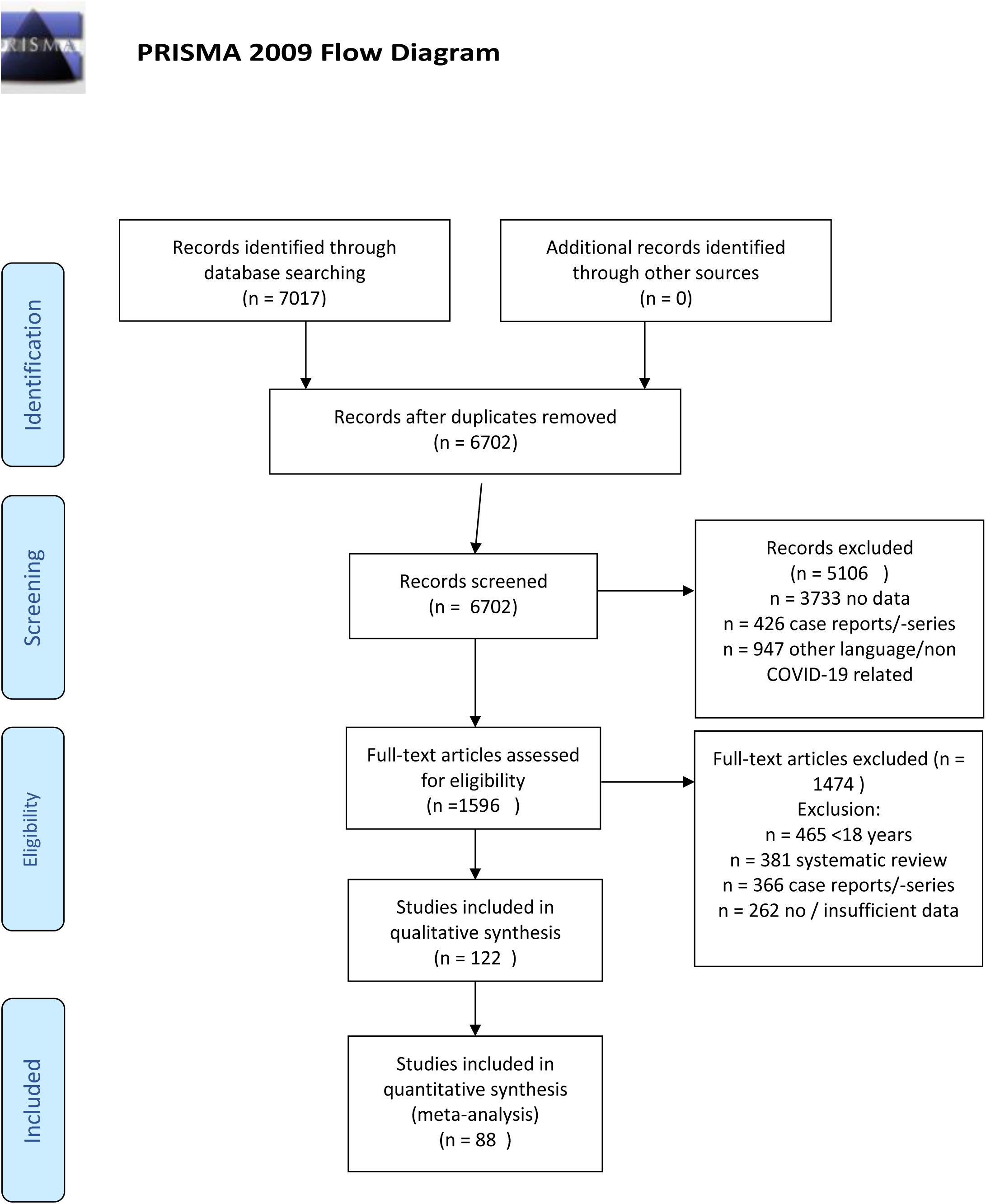
PRISMA Flow Diagram

### Study quality

The findings on study quality can be found in Figure 2. When considering the case and severity definition of COVID-19 almost 50% of studies were considered low risk of bias, while only 9.1% had a high risk. In contrast, many studies were identified to have high concerns for bias in respect to patient selection and generalizability of findings (36.4% high risk, 9.1% low risk). In more than a third of studies, we had high concern that the full data on patients were not available and inappropriate exclusion might have occurred (35.2%). The full explanation of the risk of bias assessment and the assessment of each paper individually is available in the supplement S4. Overall, high- or intermediate risk of bias for at least one category was found in almost three fourth (73.8%) of studies. No study scored low risk in all three categories.

**Figure 2.**
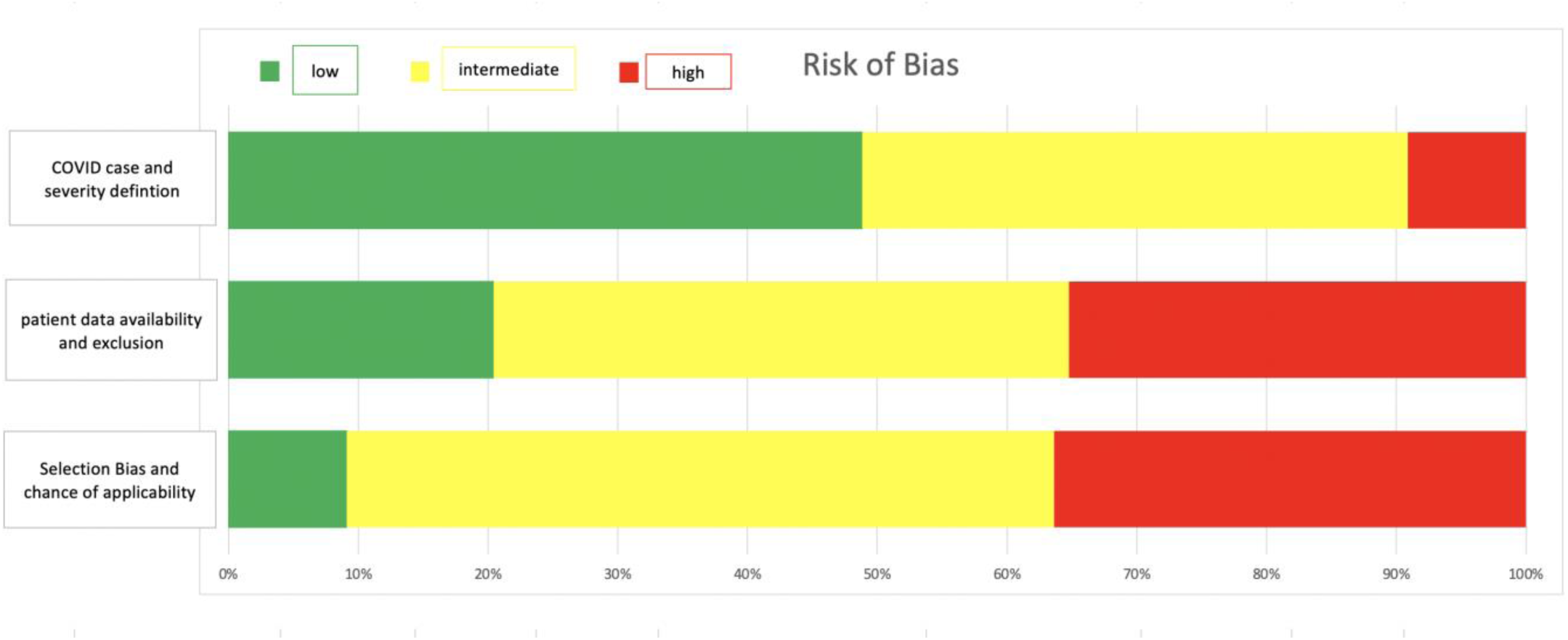
Risk of bias assessment

### ICU admission

Figure 3A and Table 1 show the pooled odds ratios (OR) and differences of medians, respectively, for ICU admission for the different indicators in the five categories: demographic, symptoms, comorbidities, laboratory and clinical values [12, 13, 27, 34-55].

**Table 1.**
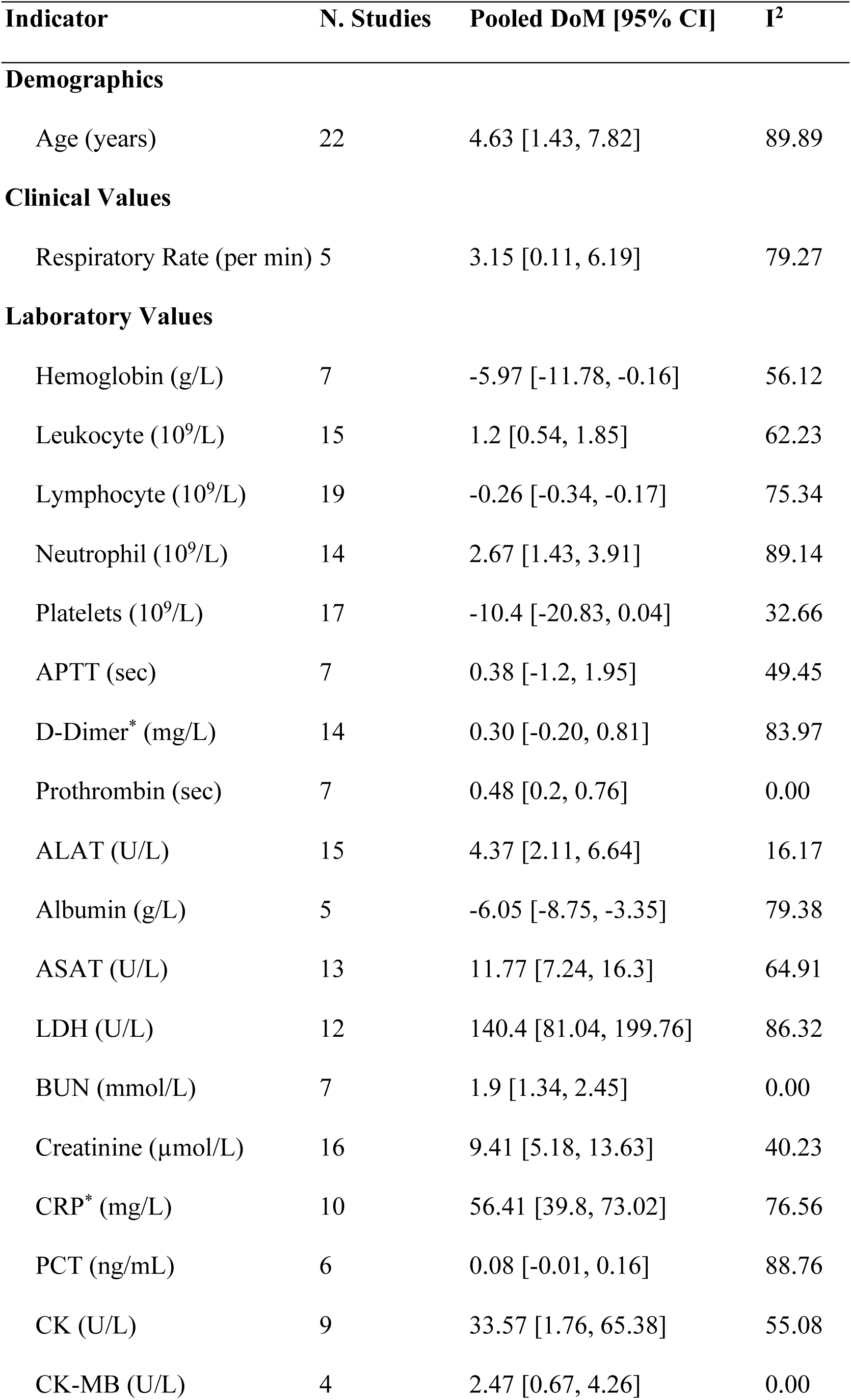
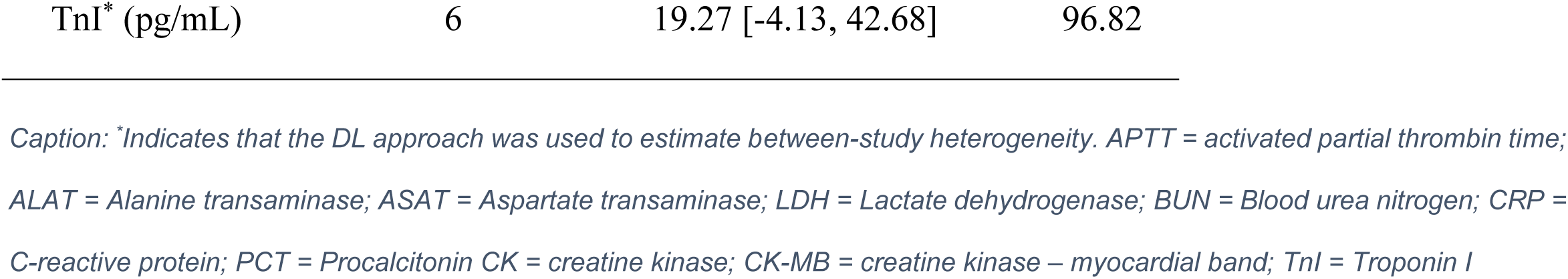
Summary of the meta-analysis results for continuous indicators comparing those who were admitted to the ICU and those who were not

**Figure 3.**
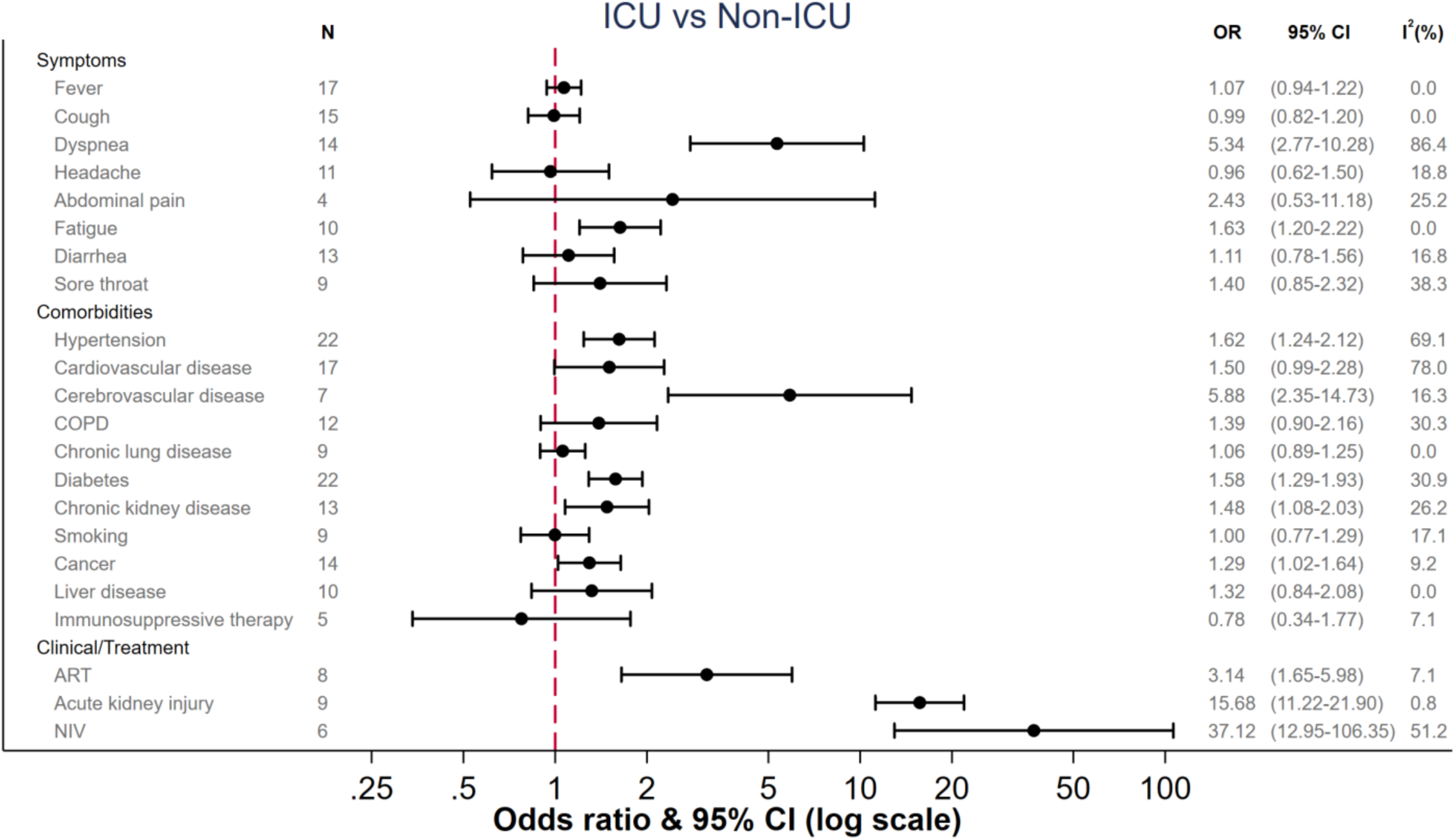
Pooled odds ratios among ICU vs. non ICU groups Caption: COPD = chronic obstructive pulmonary disease, ART = anti-retroviral therapy, NIV = non-invasive ventilation, OR = odds ratio, CI = Confidence Interval

**Figure 3b.**
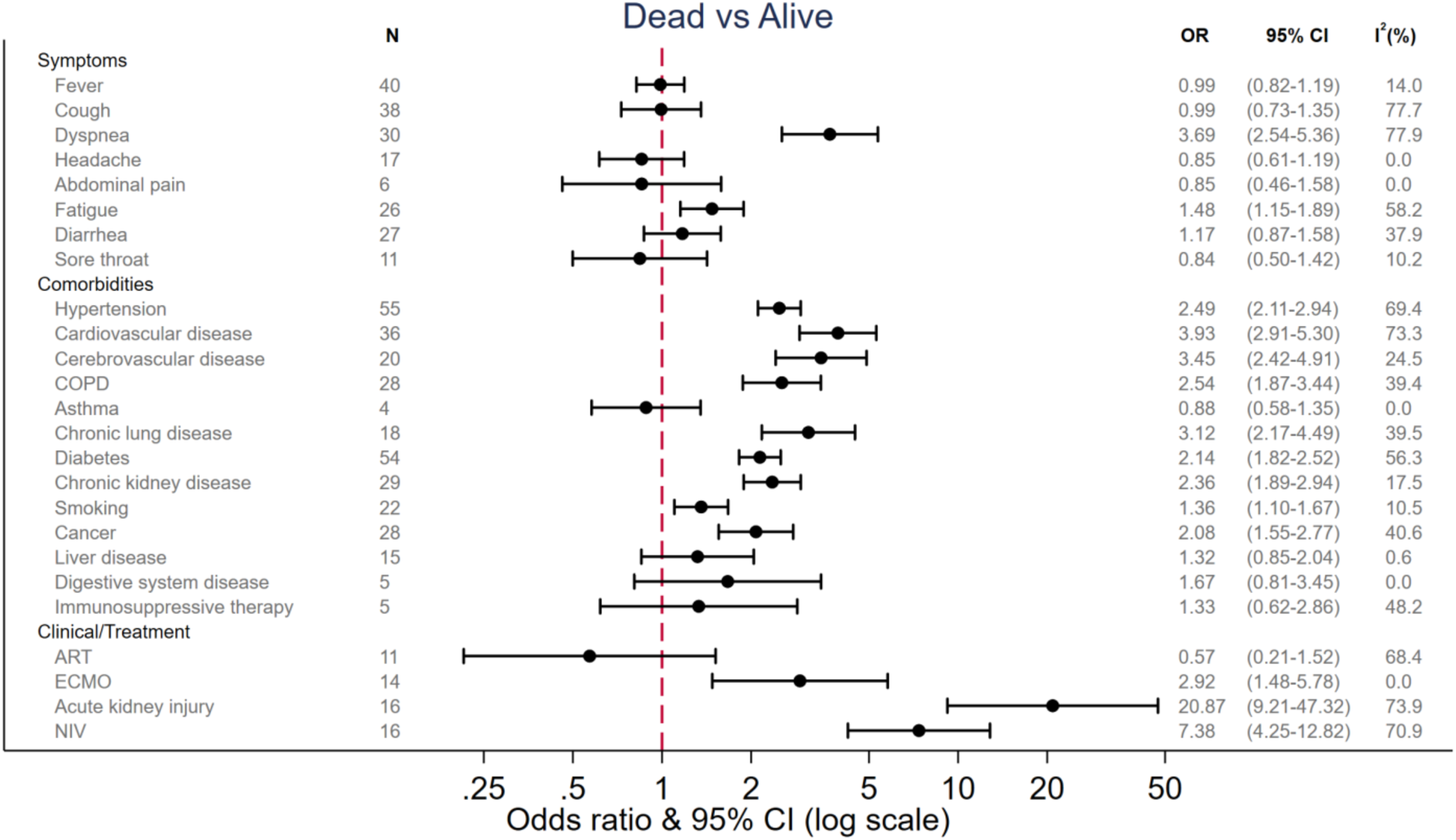
Pooled odds ratios among mortality vs. survival groups Caption: COPD = chronic obstructive pulmonary disease, ART = anti-retroviral therapy, NIV = non-invasive ventilation, ECMO = extra corporal membrane oxygenation, OR = odds ratio, CI = Confidence Interval

Patients requiring ICU admission had a median age of 65 years (CI 62.27 to 66.16). Those not requiring ICU admission were significantly younger with a median age of 59 years (CI 55.93 to 61.86) with a DoM of 4.63 years (CI 1.43 to 7.82) (Table 1). We were not able to perform a subgroup analysis of different age groups as data provided by primary studies was insufficient.

Of the many possible symptoms of COVID-19, we found dyspnea (OR 5.34, CI 2.77 to 10.28) and fatigue (OR 1.63, CI 1.20 to 2.22) to be significantly associated with ICU admission. In terms of co-morbidities, patients admitted to the ICU were more likely to suffer from cerebrovascular disease (OR 5.88, CI 2.35 to 14.73), hypertension (OR 1.62 CI 1.24 to 2.12), diabetes (OR 1.58, CI 1.29 to 1.93) and chronic kidney disease (OR 1.48, CI 1.08 to 2.03). In contrast, cardiovascular diseases (OR 1.50, CI 0.99 to 2.28), chronic obstructive pulmonary disease (COPD) (OR 1.39, CI 0.90 to 2.16), chronic lung disease (OR 1.06, CI 0.89 to 1.25) and smoking (OR 1.00, CI 0.77 to 1.29) were not associated with ICU admission.

Few laboratory values showed differences between patients that required ICU admission and those who did not (Table 1). D-Dimer failed to show a statistically significant difference (DoM 0.3 mg/L, CI −0.2 to 0.81). We found a clinically relevant elevation of CRP and cardiac Troponin I (cTnI) in patients requiring ICU admission, although cTnI failed to be statistically significant (DoM for CRP 56.41 mg/L, CI 39.8 to 73.02 and DoM for cTnI 19.27 pg/mL, CI −4.13 to 42.68). A clinically significant reduction in lymphocytes was also observed (DoM −0.34, CI −0.39 to −0.29). Leukocytes, neutrophiles and LDH were also significantly higher in patients admitted to an ICU, but the absolute elevation over those in non-ICU patients were small and of questionable clinical relevance (Table 1).

Patients developing acute kidney failure, as a complication at any stage, had the highest risk for ICU admission (OR 15.69, CI 11.22 to 21.90).

### Mortality

Figure 3B and Table 2 show the pooled odds ratios and differences of medians, respectively, for mortality for symptoms, comorbidities, laboratory and clinical values [11, 16, 26, 56-108].

**Table 2.** Summary of the meta-analysis results for continuous indicators comparing those who died and those who survived

| Indicator | N. Studies | Pooled DoM [95% CI] | I<sup>2</sup> |
| --- | --- | --- | --- |
| **Demographics** |  |  |  |
| Age (years) | 52 | 13.15 [11.37, 14.94] | 86.74 |
| **Clinical Values** |  |  |  |
| SpO2 - without O2 (%) | 15 | -6.33 [-8.14, -4.52] | 81.77 |
| Respiratory Rate (per min) | 15 | 3.41 [2.26, 4.55] | 62.32 |
| **Laboratory Values** |  |  |  |
| Hemoglobin (g/L) | 18 | -2.66 [-5.12, -0.2] | 43.36 |
| Leukocyte (10<sup>9</sup>/L) | 37 | 2.79 [2.23, 3.35] | 70.35 |
| Lymphocyte (10<sup>9</sup>/L) | 38 | -0.34 [-0.39, -0.29] | 70.03 |
| Neutrophil (10<sup>9</sup>/L) | 25 | 3.26 [2.56, 3.95] | 82.2 |
| Platelets (10<sup>9</sup>/L) | 30 | -31.94 [-41.11, -22.77] | 58.13 |
| APTT (sec) | 16 | 0.59 [-0.51, 1.69] | 61.88 |
| D-Dimer (mg/L)* | 30 | 1.29 [0.90, 1.69] | 81.53 |
| Fibrinogen (g/L) | 7 | 0.01 [-0.12, 0.15] | 0.00 |
| INR | 7 | 0.06 [0.01, 0.12] | 63.31 |
| Prothrombin (sec) | 25 | 0.91 [0.67, 1.14] | 54.65 |
| ALAT (U/L) | 34 | 4.43 [2.41, 6.46] | 26.64 |
| Albumin (g/L) | 21 | -4.64 [-5.83, -3.45] | 85.16 |
| ASAT (U/L) | 27 | 13.35 [10.54, 16.15] | 42.83 |
| LDH (U/L) | 23 | 189.49 [155, 223.98] | 75.03 |
| BUN (mmol/L) | 17 | 2.77 [2.07, 3.46] | 66.77 |
| Creatinine (μmol/L) | 29 | 15.3 [10.3, 20.29] | 61.63 |
| CRP (mg/L)* | 34 | 69.1 [50.43, 87.77] | 95.99 |
| IL-6 (pg/mL) | 11 | 31.19 [11.96, 50.41] | 99.75 |
| PCT (ng/mL) | 18 | 0.16 [0.1, 0.22] | 68.09 |
| BNP (pg/mL) | 7 | 405.26 [116.51, 694.02] | 95.81 |
| CK (U/L) | 18 | 64.09 [29.04, 99.13] | 81.47 |
| CK-MB (U/L) | 9 | 3.66 [1.19, 6.14] | 67.12 |
| TnI (pg/mL)* | 13 | 21.88 [9.78, 33.99] | 75.17 |

*Caption: \*Indicates that the DL approach was used to estimate between-study heterogeneity. SpO2 = Oxygen saturation; APTT = activated partial thrombin time; INR = Internationalized normalized ratio; ALAT = Alanine transaminase; ASAT = Aspartate transaminase; LDH = Lactate dehydrogenase; BUN = Blood urea nitrogen; CRP = C-reactive protein; IL-6 = Interleukin-6; BNP = brain natriuretic peptide; PCT = Procalcitonin CK = creatine kinase; CK-MB = creatine kinase – myocardial band; TnI = Troponin I*

Patients who died had a median age of 71 years (CI 69.3 to 71.61) compared to survivors with a median age of 58 years (CI 55.03 to 59.4) for a DoM of 13.15 years (CI 11.37 to 14.94] (Table 2). Again, dyspnea was the symptom that differentiated markedly between survivors and non-survivors (OR 3.69, CI 2.54 to 5.36). Also, fatigue was more frequently observed in those who died (OR 1.48, CI 1.15 to 1.89).

Patients who died were more likely to suffer from cardiovascular disease (OR 3.93, CI 2.91 to 5.30), cerebrovascular disease (OR 3.45, CI 2.42 to 4.91), chronic lung disease (OR 3.12, CI 2.17 to 4.49), COPD (OR 2.54, CI 1.87 to 3.44; Table 3B) and hypertension (OR 2.49, CI 2.11 to 2.94). Current and former smokers had an increased risk of mortality (OR 1.36, CI 1.10 to 1.67). Patients with chronic kidney disease (CKD) (OR 2.36, CI 1.89 to 2.94), diabetes (OR 2.14, CI 1.82 to 2.52) and cancer (OR 2.08, CI 1.55 to 2.77) also had an increased risk. Co-morbidities not associated with an increased risk of mortality were asthma, liver disease, digestive system disease and immunosuppressive therapy (Table 3B). Clinically relevant elevations outside the normal laboratory range in patients who died compared to those who survived were observed in two markers of inflammation: CRP was elevated by 69.1mg/L (CI 50.43 to 87.77) and IL-6 by 31.19 pg/mL (CI 11.96 to 50.41). Furthermore, clinically significant elevations were observed in cTnI by 21.88pg/mL (CI 9.78 to 33.99) and D-Dimer by 1.29mg/L (CI 0.9 to 1.69), while lymphocytes were significantly lower: −0.34×10^9^/L (CI −0.39 to −0.29). Other makers (hemoglobin, leukocytes, neutrophils, platelets, international normalized ratio (INR), Prothrombin, alanine transaminase (ALAT), aspartate transaminase (ASAT), Albumin, LDH, blood urea nitrogen (BUN), Creatinine, PCT, BNP, CK and creatine kinase myocardial band (CK-MB)) were also significantly elevated in those who died, however, the absolute difference compared to those who survived was small and thus likely not clinically relevant. For leukocytes, neutrophils, platelets, prothrombin, ALAT, ASAT, BUN, Creatinine, CK and CK-MB the point estimates even stayed within the normal laboratory range.

As a clinical complication, acute kidney injury showed the highest overall risk ratio for mortality (OR 20.87, CI 9.21 to 47.32), followed by requiring non-invasive ventilation (NIV) (OR 7.38, CI 4.25 to 12.82). Patients who died presented with a median peripheral oxygen saturation (SpO2) on room air of 89% (CI 87.32 to 90.91) to the hospital, while those who survived had 95% (CI 94.59 to 96.63) (DoM −6.33%, CI −8.14 to −4.52).

Figure 4 shows pooled median estimates along with their normal laboratory ranges for selected number of indicators among patients who died, patients who survived, ICU-admitted patients, and non-ICU admitted patients. Pooled difference of medians estimates for all indicators are available in the supplementary files (S5 for mortality, S6 for ICU admission, S7 for intubation and hospitalization in S8). After removing large outliers in a sensitivity analyses for CRP and D-Dimer, results did not change substantially (results available in the supplementary file S9). Funnel plots showed no substantial asymmetry suggesting publication bias except for data assessing acute kidney injury (supplementary file S10 for mortality and S11 for ICU admission).

**Figure 4.**
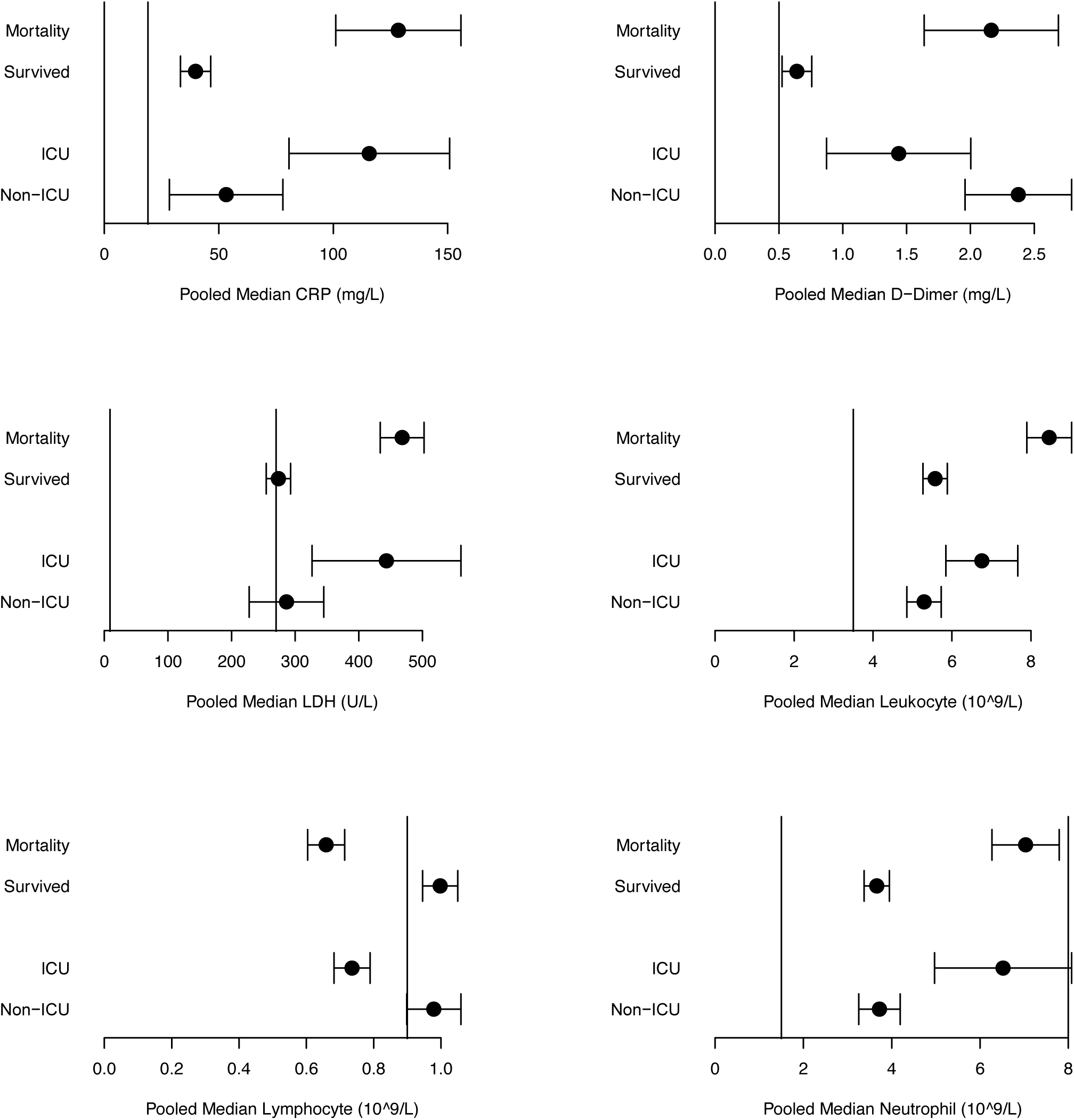
Pooled median estimates of selected indicators along with their normal laboratory ranges among patients who died, patients who survived, ICU-admitted patients, and non-ICU admitted patients Caption: Normal range (grey) was used as stated in included publications. If different normal ranges are reported in included publications, we used the lowest and highest values. Leukocytes and Lymphocytes upper range of the normal range is 11.0×10^9/L and 4.0×10^9/, respectively. CRP = C-reactive protein; LDH = Lactate dehydrogenase;

## Discussion

In this comprehensive systematic review and meta-analysis, we confirm known markers of severe disease for COVID-19 and shed light on further indicators, whose significance was indeterminate to date.

With respect to co-morbidities, we confirm cardiovascular disease (OR 3.93), chronic lung disease (OR 3.12) and COPD (OR 2.54) as strong risk factors of mortality among COVID-19 patients but not for ICU admission. Only cerebrovascular disease was strongly associated with an increased risk of both ICU admission and death (almost six- and three-fold higher ratio for ICU admission and death, respectively). Overall, the finding that cerebrovascular disease is associated with poor outcomes is in line with the more recent data highlighting the importance of delirium and an overall depressed mental state in severe COVID-19 [109-111]. Our findings confirm chronic kidney disease, diabetes and COPD/chronic lung disease as risk factors, however, associations are less strong than those for cardiovascular or cerebrovascular disease [109]. Evidence from previous studies regarding the risk associated with hypertension has been inconclusive. Our work identifies hypertension as a clear risk factor for ICU admission (OR 1.62, CI 1.24 to 2.12) and death (OR 2.49, CI 2.11 to 2.94) [7, 17, 18]. Similarly, while prior data were inconclusive with respect to the influence of smoking for severe COVID-19 [20-23], our meta-analysis shows the increased risk of mortality among smokers (OR 1.36, CI 1.10 to 1.67). However, our data did not allow for meta-regression to assess whether this effect was independent of the risk associated with chronic lung disease. In line with some recent studies on asthma, we could not find an increased risk for mortality [112] in our meta-analysis (OR 0.88, CI 0.58 to 1.35).

CRP was the only laboratory marker that was associated with a higher risk of ICU admission (DoM 56.41 mg/L) and death (DoM 69.1 mg/L), while D-Dimer elevation was only significantly associated with death (DoM 1.29 mg/L), but not with ICU admission. Although the median elevation of cTnI was clinically relevant both in those who were admitted to the ICU (DoM 19.27 pg/mL) and those who died (DoM 21.88 pg/mL), only in those who died was the difference statistically significant.

We were able to confirm clinically relevant lymphopenia as a marker. Lymphopenia was a marker that was used early on for triage purposes to predict disease severity [113] and our findings confirm the systematic review results on this topic published by Huang and Pranata [114].

### Strengths and limitations of this study

Our study provides a comprehensive review of the data from both pre-print and peer-reviewed sources with a broad geographic distribution and assesses the different categories of risk factors from symptoms, co-morbidities, laboratory values to clinical complications. Correlating the indicators to the two clinical outcomes death and ICU admission has both strength and limitations. While ICU admission is a clinical decision, it is, especially early on in a new disease, sometimes a measure of precaution. This might weaken the association of indicators with clinical outcomes. At the same time, when capacity of ICU beds is exhausted, triage decisions might have been made based on age and co-morbidities to not admit to the ICU, thus strengthening an association of an indicator beyond what would be expected under routine conditions. We also assessed the association with hospitalization and intubation (see Supplement), but here confounding factors seemed to be even more pronounced, and data are further limited. In addition, with improving care and novel therapies certain associations might be less pronounced. We did not observe improved survival of antiviral therapy in the studies included, suggesting that this effect might not yet have occurred in the timeframe of studies included here.

Additional limitations primarily relate to data quality of the included studies. Our quality assessment of studies clearly indicated that substantial bias was present across studies. Primarily the selection bias as suggested for example by the high case fatality rate (e.g. Zhou et al. [11], 28.3%, Chen et al. [115] 11.1%, and Huang et al. [12] 14.6%) is likely to have impacted our results and prospective data collection to confirm findings of these studies is important [116]. In addition, we found a large number of studies (n=21, list available in supplementary file S3) that included laboratory values that were obviously incorrect, which suggests that despite peer-review in some of them, the rush of publication in this pandemic impacted the quality of reporting [117].

## Conclusion

Our data on mortality and ICU admission confirms most of the proposed indicators of clinical outcomes, clarifies the strength of association and highlights additional indicators. In addition, this systematic review highlights the limitations of the studies published and calls for better quality in prospective collections.

## Data Availability

All available data is published either in the manuscript or the supplementary file. For more data please contact the author.

## Acknowledgements

We thank Genevieve Gore for her help with the search terms. We thank Stephani Schmitz, Sara Kraker, Marlene Ganslmeier and Amelie Muth for their help with the data extraction and Sinan Onogur, Laura Aguilera Saiz, and Bünyamin Pekdemir for their help setting up the data extraction tool.

## Funding

SK, AJZ, CG, JG, AS, LMH, GG, AB, JL, MAW, SM, SS, SaK, MG, AM, SO, LAS, BP and CMD declare no conflict of interest.

SM acknowledges support from the National Science Foundation Graduate Research Fellowship Program under Grant No. DGE1745303, National Library Of Medicine of the National Institutes of Health under Award Number T32LM012411, and Fonds de recherche du Québec-Nature et technologies B1X research scholarship.

CMD acknowledges the support of the Heidelberg University Hospital and the German Center of Infectious Disease, where Heidelberg is a core site as part of the tuberculosis focus group.

The work was supported by Heidelberg University Hospital internal funds

Any opinions, findings, and conclusions or recommendations expressed in this material are those of the author(s) and do not necessarily reflect the views of the funding agencies.

## Notes

### Competing Interest Statement

The authors have declared no competing interest.

### Author Declarations

This is not a clinical trial. It was registered at Prospero Registrationnumber:CRD42020177154

