## Supplementary File for "Can we predict the severe course of COVID-19 – a systematic review and meta-analysis of indicators of clinical outcome?"

### S1 Search strategy

**Search Terms for Pubmed**

**Search with concept of COVID-19 AND (predictive variables OR clinically relevant outcomes) conducted on 04.04.2020:**

("severe acute respiratory syndrome coronavirus 2"[Supplementary Concept] OR "severe acute respiratory syndrome coronavirus 2"[tw] OR "ncov"[tw] OR "n cov"[tw] OR "2019nCoV"[tw] OR "covid-19"[supplementary concept] OR "covid"[tw] OR "covid19"[tw] OR "sars cov 2"[tw] OR "sarscov2"[tw] OR "sars2"[tw] OR "sars 2"[tw] OR "new coronavirus"[tw] OR "new coronaviruses"[tw] OR ((("wuhan"[All Fields] OR "china"[mesh] OR "china"[all fields] OR "chinese"[all fields] OR novel[tw] OR 2019[tw]) AND ("coronavirus"[MeSH Terms] OR "coronavirus"[tw] OR "coronaviruses"[tw] OR "corona virus"[tw] OR "corona viruses"[tw] OR "pneumonia virus"[tw] OR "pneumonia viruses"[tw] OR "hcov"[tw] OR "h cov"[tw])) AND 2019/12/1 : 2030/12/31[Date - Publication])) AND (("risk"[mesh] OR risk factor*[tw] OR risk*[ti] OR predict*[tw] OR "epidemiology"[sh] OR "epidemiologic factors"[mesh] OR epidemiolog*[tw] OR age factor*[tw] OR age*[ti] OR sex*[ti] OR ((sex[tw] OR age[tw]) AND variable*[tw]) OR comorbid*[tw] OR multimorbid*[tw] OR multiple morbidit*[tw] OR pre-existing[tw] OR preexisting[tw] OR "clinical laboratory techniques"[mesh] OR "lymphopenia"[mesh] OR "fibrin fragment D"[Supplementary Concept] OR lymphopeni*[tw] OR lymphopaeni*[tw] OR "Leukopenia"[Mesh] OR leukopeni*[tw] OR leukopaeni*[tw] OR "Ferritins"[Mesh] OR ferritin*[tw] OR "C-Reactive Protein"[Mesh] OR "crp"[tw] OR c reactive protein*[tw] OR "Procalcitonin"[Mesh] OR procalcitonin*[tw] OR "Transferrin"[Mesh] OR transferrin*[tw] OR ldh[tw] OR tnt[tw] OR prebnp[tw] OR pro bnp[tw] OR d dimer*[tw] OR clinical*[tw] OR laborator*[tw] OR demograph*[tw] OR baseline[tw] OR sever*[tw] OR "Smoking"[Mesh] OR "Smokers"[Mesh] OR smoking[tw] OR smoker*[tw] OR o2 requirement*[tw] OR o2 consumption*[tw] OR oxygen requirement*[tw] OR oxygen consumption*[tw] OR "intubation"[mesh] OR intubat*[tw] OR "Immunosuppression"[Mesh] OR "Immunocompromised Host"[Mesh] OR immunosuppress*[tw] OR immunocompromis*[tw] OR immune suppress*[tw] OR immune compromis*[tw] OR "Heart Diseases"[Mesh] OR "Lung Diseases"[Mesh] OR heart disease*[tw] OR cardiac disease*[tw] OR cardiovascular disease*[tw] OR lung disease*[tw] OR pulmonary disease*[tw]) OR ("prognosis"[mesh] OR prognos*[tw] OR fatal*[tw] OR death*[tw] OR surviv*[tw] OR outcome*[tw] OR "mortality"[mesh] OR "mortality"[sh] OR mortalit*[tw] OR morbidit*[tw] OR "hospitalization"[mesh] OR hospitaliz*[tw] OR hospitalis*[tw] OR "ventilation"[mesh] OR ventilat*[tw] OR "intensive care units"[mesh] OR intensive care[tw] OR ICU[tw] OR ICUs[tw] OR PICU[tw] OR PICUs[tw] OR NICU[tw] OR NICUs[tw] OR intermediate care[tw] OR IMCU[tw] OR IMCUs[tw] OR "critical care"[mesh] OR critical care[tw] OR critically ill[tw] OR critical ill*[tw] OR "patient outcome assessment"[mesh] OR outcome*[tw] OR CFR[tw]))

**Search with concept of COVID AND predictive variables AND clinically relevant outcomes conducted on 31.05.2020:**

("severe acute respiratory syndrome coronavirus 2"[Supplementary Concept] OR "severe acute respiratory syndrome coronavirus 2"[tw] OR "ncov"[tw] OR "n cov"[tw] OR "2019nCoV"[tw] OR "covid-19"[supplementary concept] OR "covid"[tw] OR "covid19"[tw] OR "sars cov 2"[tw] OR "sarscov2"[tw] OR "sars2"[tw] OR "sars 2"[tw] OR "new coronavirus"[tw] OR "new coronaviruses"[tw] OR ((("wuhan"[All Fields] OR "china"[mesh] OR "china"[all fields] OR "chinese"[all fields] OR novel[tw] OR 2019[tw]) AND ("coronavirus"[MeSH Terms] OR "coronavirus"[tw] OR "coronaviruses"[tw] OR "corona virus"[tw] OR "corona viruses"[tw] OR "pneumonia virus"[tw] OR "pneumonia viruses"[tw] OR "hcov"[tw] OR "h cov"[tw])) AND 2019/12/1 : 2030/12/31[Date - Publication])) AND (("risk"[mesh] OR risk factor*[tw] OR risk*[ti] OR predict*[tw] OR "epidemiology"[sh] OR "epidemiologic factors"[mesh] OR epidemiolog*[tw] OR age factor*[tw] OR age*[ti] OR sex*[ti] OR ((sex[tw] OR age[tw]) AND variable*[tw]) OR comorbid*[tw] OR multimorbid*[tw] OR multiple morbidit*[tw] OR pre-existing[tw] OR preexisting[tw] OR "clinical laboratory techniques"[mesh] OR "lymphopenia"[mesh] OR "fibrin fragment D"[Supplementary Concept] OR lymphopeni*[tw] OR lymphopaeni*[tw] OR "Leukopenia"[Mesh] OR leukopeni*[tw] OR leukopaeni*[tw] OR "Ferritins"[Mesh] OR ferritin*[tw] OR "C-Reactive Protein"[Mesh] OR "crp"[tw] OR c reactive protein*[tw] OR "Procalcitonin"[Mesh] OR procalcitonin*[tw] OR "Transferrin"[Mesh] OR transferrin*[tw] OR ldh[tw] OR tnt[tw] OR prebnp[tw] OR pro bnp[tw] OR d dimer*[tw] OR clinical*[tw] OR laborator*[tw] OR demograph*[tw] OR baseline[tw] OR sever*[tw] OR "Smoking"[Mesh] OR "Smokers"[Mesh] OR smoking[tw] OR smoker*[tw] OR o2 requirement*[tw] OR o2 consumption*[tw] OR oxygen requirement*[tw] OR oxygen consumption*[tw] OR "intubation"[mesh] OR intubat*[tw] OR "Immunosuppression"[Mesh] OR "Immunocompromised Host"[Mesh] OR immunosuppress*[tw] OR immunocompromis*[tw] OR immune suppress*[tw] OR immune compromis*[tw] OR "Heart Diseases"[Mesh] OR "Lung Diseases"[Mesh] OR heart disease*[tw] OR cardiac disease*[tw] OR cardiovascular disease*[tw] OR lung disease*[tw] OR pulmonary disease*[tw]) AND ("prognosis"[mesh] OR prognos*[tw] OR fatal*[tw] OR death*[tw] OR surviv*[tw] OR outcome*[tw] OR "mortality"[mesh] OR "mortality"[sh] OR mortalit*[tw] OR morbidit*[tw] OR "hospitalization"[mesh] OR hospitaliz*[tw] OR hospitalis*[tw] OR "ventilation"[mesh] OR ventilat*[tw] OR "intensive care units"[mesh] OR intensive care[tw] OR ICU[tw] OR ICUs[tw] OR PICU[tw] OR PICUs[tw] OR NICU[tw] OR NICUs[tw] OR intermediate care[tw] OR IMCU[tw] OR IMCUs[tw] OR "critical care"[mesh] OR critical care[tw] OR critically ill[tw] OR critical ill*[tw] OR "patient outcome assessment"[mesh] OR outcome*[tw] OR CFR[tw])) AND (("2020/04/03"[PDat] : "2020/05/31"[PDat]))

**Search Terms of Web of Science (Science Citation Index Expanded (SCI-EXPANDED), Social Sciences Citation Index (SSCI), BIOSIS Previews, KCI-Korean Journal Database, Russian Science Citation Index, SciELO Citation Index)**

**Search with concept of COVID-19 AND (predictive variables OR clinically relevant outcomes) conducted on 04.04.2020:**

( TS=("severe acute respiratory syndrome coronavirus 2" OR "ncov" OR "n cov" OR "2019nCoV" OR "covid" OR "covid19" OR "sars cov 2" OR "sarscov2" OR “sars2” OR "sars 2" OR “new coronavirus*”) OR TS=(("wuhan" OR "china" OR "chinese" OR novel OR 2019) AND ("coronavirus" OR "coronaviruses" OR "corona virus" OR "corona viruses" OR "pneumonia virus" OR "pneumonia viruses" OR "hcov" OR "h cov"))) AND PY=(2019 OR 2020) AND ( ( ( TS=( “sex” OR “age”) AND TS=(variable*) ) OR TI=(“age*” OR “sex*”) OR TS=(“risk factor*”) OR TI=(“risk*”) OR TI=(“predict*” OR “epidemiolog* factor*” OR “age factor*” OR “comorbid*” OR “multimorbid*” OR “multiple morbidit*” OR “pre-existing” OR “preexisting” OR "lymphopeni*" OR “lymphopaeni*” OR “leukopeni*” OR “leukopaeni*” OR “ferritin*” OR "crp" OR “c reactive protein*” OR “procalcitonin*” OR “transferrin*” OR “ldh” OR “tnt” OR “prebnp” OR “pro bnp” OR “d dimer*” OR “clinical*” OR “laborator*” OR “demograph*” OR “baseline” OR “sever*” OR “smoking” OR “smoker*” OR “o2 requirement*” OR “o2 consumption*” OR “oxygen requirement*” OR “oxygen consumption*” OR “intubat*” OR "Immunocompromised Host" OR “immunosuppress*” OR “immunocompromis*” OR “immune suppress*” OR “immune compromis*” OR “heart disease*” OR “cardiac disease*” OR “cardiovascular disease*” OR “lung disease*” OR “pulmonary disease*”OR "epidemiology" OR "clinical laboratory techniques"OR "fibrin fragment D") ) OR TS = (“prognos*” OR “fatal*” OR “death*” OR “surviv*” OR “outcome*” OR “mortalit*” OR “morbidit*” OR “hospitaliz*” OR “hospitalis*” OR “ventilat*” OR "intensive care units" OR “intensive care” OR “ICU” OR “ICUs” OR “PICU” OR “PICUs” OR “NICU” OR “NICUs” OR “intermediate care” OR “IMCU” OR “IMCUs” OR “critical care” OR “critically ill” OR “critical ill*” OR “outcome*” OR “CFR” OR "patient outcome assessment") )

**Search for preprint Paper on medRxiv and bioRxiv**

All COVID-19 SARS-CoV-2 preprints from medRxiv and bioRxiv were included, that were listed on [https://connect.biorxiv.org/relate/content/181 on 04.04.2020](https://connect.biorxiv.org/relate/content/181%20on%2004.04.2020)

### S2 PRISMA Checklist and PROSPERO Protocol

| **Section/topic** | **#** | **Checklist item** | **Reported on page #** |
| --- | --- | --- | --- |
| **TITLE** | | |  |
| Title | 1 | Identify the report as a systematic review, meta-analysis, or both. | 1 |
| **ABSTRACT** | | |  |
| Structured summary | 2 | Provide a structured summary including, as applicable: background; objectives; data sources; study eligibility criteria, participants, and interventions; study appraisal and synthesis methods; results; limitations; conclusions and implications of key findings; systematic review registration number. | 2 |
| **INTRODUCTION** | | |  |
| Rationale | 3 | Describe the rationale for the review in the context of what is already known. | 3 |
| Objectives | 4 | Provide an explicit statement of questions being addressed with reference to participants, interventions, comparisons, outcomes, and study design (PICOS). | 4,5 |
| **METHODS** | | |  |
| Protocol and registration | 5 | Indicate if a review protocol exists, if and where it can be accessed (e.g., Web address), and, if available, provide registration information including registration number. | S2 |
| Eligibility criteria | 6 | Specify study characteristics (e.g., PICOS, length of follow-up) and report characteristics (e.g., years considered, language, publication status) used as criteria for eligibility, giving rationale. | 4 |
| Information sources | 7 | Describe all information sources (e.g., databases with dates of coverage, contact with study authors to identify additional studies) in the search and date last searched. | 4,5 |
| Search | 8 | Present full electronic search strategy for at least one database, including any limits used, such that it could be repeated. | S1 |
| Study selection | 9 | State the process for selecting studies (i.e., screening, eligibility, included in systematic review, and, if applicable, included in the meta-analysis). | 4,5 |
| Data collection process | 10 | Describe method of data extraction from reports (e.g., piloted forms, independently, in duplicate) and any processes for obtaining and confirming data from investigators. | 4,5 |
| Data items | 11 | List and define all variables for which data were sought (e.g., PICOS, funding sources) and any assumptions and simplifications made. | 4,5; S13 |
| Risk of bias in individual studies | 12 | Describe methods used for assessing risk of bias of individual studies (including specification of whether this was done at the study or outcome level), and how this information is to be used in any data synthesis. | 4,5; S5 |
| Summary measures | 13 | State the principal summary measures (e.g., risk ratio, difference in means). | 4,5 |
| Synthesis of results | 14 | Describe the methods of handling data and combining results of studies, if done, including measures of consistency (e.g., I^2^) for each meta-analysis. | 4,5 |

Page 1 of 2

| **Section/topic** | **#** | **Checklist item** | **Reported on page #** |
| --- | --- | --- | --- |
| Risk of bias across studies | 15 | Specify any assessment of risk of bias that may affect the cumulative evidence (e.g., publication bias, selective reporting within studies). | 6, 13, S5 |
| Additional analyses | 16 | Describe methods of additional analyses (e.g., sensitivity or subgroup analyses, meta-regression), if done, indicating which were pre-specified. | 7, S12 |
| **RESULTS** | | |  |
| Study selection | 17 | Give numbers of studies screened, assessed for eligibility, and included in the review, with reasons for exclusions at each stage, ideally with a flow diagram. | 6 |
| Study characteristics | 18 | For each study, present characteristics for which data were extracted (e.g., study size, PICOS, follow-up period) and provide the citations. | 6,7; S4, S13 |
| Risk of bias within studies | 19 | Present data on risk of bias of each study and, if available, any outcome level assessment (see item 12). | S5 |
| Results of individual studies | 20 | For all outcomes considered (benefits or harms), present, for each study: (a) simple summary data for each intervention group (b) effect estimates and confidence intervals, ideally with a forest plot. | 13-16  S6,S7,S8,S9 |
| Synthesis of results | 21 | Present results of each meta-analysis done, including confidence intervals and measures of consistency. | 13-16  S6,S7,S8,S9 |
| Risk of bias across studies | 22 | Present results of any assessment of risk of bias across studies (see Item 15). | 6, 13, S5 |
| Additional analysis | 23 | Give results of additional analyses, if done (e.g., sensitivity or subgroup analyses, meta-regression [see Item 16]). | 7, S12 |
| **DISCUSSION** | | |  |
| Summary of evidence | 24 | Summarize the main findings including the strength of evidence for each main outcome; consider their relevance to key groups (e.g., healthcare providers, users, and policy makers). | 8,9 |
| Limitations | 25 | Discuss limitations at study and outcome level (e.g., risk of bias), and at review-level (e.g., incomplete retrieval of identified research, reporting bias). | 8,9 |
| Conclusions | 26 | Provide a general interpretation of the results in the context of other evidence, and implications for future research. | 9 |
| **FUNDING** | | |  |
| Funding | 27 | Describe sources of funding for the systematic review and other support (e.g., supply of data); role of funders for the systematic review. | 9 |

*From:* Moher D, Liberati A, Tetzlaff J, Altman DG, The PRISMA Group (2009). Preferred Reporting Items for Systematic Reviews and Meta-Analyses: The PRISMA Statement. PLoS Med 6(7): e1000097. doi:10.1371/journal.pmed1000097

For more information, visit: **www.prisma-statement.org**.

### The PROSPERO Protocol is available under the number: CRD42020177154

### S3 List of laboratory parameters of eligible studies not included in the meta-analysis due to unphysiological values

| **Publication** | **Indicator** | **Outcome** |
| --- | --- | --- |
| Wang, K., et al. The experience of high-flow nasal cannula in hospitalized patients with 2019 novel coronavirus-infected pneumonia in two hospitals of Chongqing, China. Ann. Intensive Care 10, 37 (2020). https://doi.org/10.1186/s13613-020-00653-z | Hemoglobin | Hospitalized vs. non hospitalized |
| Guo, W., et al. Diabetes is a risk factor for the progression and prognosis of COVID‐19. Diabetes Metab Res Rev. 2020; 36:e3319. https://doi.org/10.1002/dmrr.3319 | Hemoglobin | Hospitalized vs. non hospitalized |
| Elisa Maria Stroppa, et al. Coronavirus disease-2019 in cancer patients. A report of the first 25 cancer patients in a western country (Italy) Future Oncology 2020 16:20, 1425-1432 | Lymphocytes | Mortality vs. survived |
| Goicoechea, Marian, et al. COVID-19: clinical course and outcomes of 36 hemodialysis patients in Spain Kidney International, Volume 98, Issue 1, 27 - 34 | Hemoglobin | Mortality vs. survived |
| Zhang, F., et al. Obesity predisposes to the risk of higher mortality in young COVID‐19 patients. J Med Virol. 2020; 92: 2536– 2542. https://doi.org/10.1002/jmv.26039 | Lymphocytes | Mortality vs. survived |
| Bicheng Zhang, et al. Clinical characteristics of 82 death cases with COVID-19 medRxiv 2020.02.26.20028191; doi: https://doi.org/10.1101/2020.02.26.20028191 | CD-4 | Mortality vs. survived |
| Desborough, Michael J.R., et al. Image-proven thromboembolism in patients with severe COVID-19 in a tertiary critical care unit in the United Kingdom Thrombosis Research, Volume 193, 1 - 4 | CRP | Mortality vs. survived |
| Crespo, M., et al. COVID‐19 in elderly kidney transplant recipients. Am J Transplant. 2020; 20: 2883– 2889. https://doi.org/10.1111/ajt.16096 | IL-6 | Mortality vs. survived |
| XU, Bo, et al. Suppressed T cell-mediated immunity in patients with COVID-19: A clinical retrospective study in Wuhan, China Journal of Infection, Volume 81, Issue 1, e51 - e60 | PCT | Mortality vs. survived |
| Zhang, L., et al. Clinical characteristics of COVID-19-infected cancer patients: a retrospective case study in three hospitals within Wuhan, China Annals of Oncology, Volume 31, Issue 7, 894 - 901 | Creatinine | Hospitalized vs. non hospitalized |
| Fan, Hua et al. Cardiac injuries in patients with coronavirus disease 2019: Not to be ignored  International Journal of Infectious Diseases, Volume 96, 294 - 297 | D-Dimer | Mortality vs. survived |
| Martín‐Moro, F. et al. (2020), Survival study of hospitalised patients with concurrent COVID‐19 and haematological malignancies. Br J Haematol, 190: e16-e20. doi:10.1111/bjh.16801 | D-Dimer | Mortality vs. survived |
| Qiao Shi, et al. Clinical Characteristics and Risk Factors for Mortality of COVID-19 Patients With Diabetes in Wuhan, China: A Two-Center, Retrospective Study  Diabetes Care Jul 2020, 43 (7) 1382-1391; DOI: 10.2337/dc20-0598 | D-Dimer | Mortality vs. survived |
| Jianlei Cao et al. Clinical Features and Short-term Outcomes of 102 Patients with Coronavirus Disease 2019 in Wuhan, China, Clinical Infectious Diseases, Volume 71, Issue 15, 1 August 2020, Pages 748–755, https://doi.org/10.1093/cid/ciaa243 | D-Dimer | Mortality vs. survived |
| Yang, Xiao et al. Extracorporeal Membrane Oxygenation for Coronavirus Disease 2019-Induced Acute Respiratory Distress Syndrome: A Multicenter Descriptive Study*, Critical Care Medicine: September 2020 - Volume 48 - Issue 9 - p 1289-1295 doi: 10.1097/CCM.0000000000004447 | D-Dimer | Mortality vs. survived |
| Pan F. et al Factors associated with death outcome in patients with severe coronavirus disease-19 (COVID-19): a case-control study. Int J Med Sci 2020; 17(9):1281-1292. doi:10.7150/ijms.46614. Available from http://www.medsci.org/v17p1281.htm | Troponin I | Mortality vs. survived |
| Rong-Hui Du et al., Predictors of mortality for patients with COVID-19 pneumonia caused by SARS-CoV-2: a prospective cohort study, European Respiratory Journal May 2020, 55 (5) 2000524; DOI: 10.1183/13993003.00524-2020 | Troponin I | Mortality vs. survived |
| Zhihua Wang et al. Elevated serum IgM levels indicate poor outcome in patients with coronavirus disease 2019 pneumonia: A retrospective case-control study  medRxiv 2020.03.22.20041285; doi: https://doi.org/10.1101/2020.03.22.20041285 | D-Dimer | Mortality vs. survived |
| Zhang, J. et al. The clinical data from 19 critically ill patients with coronavirus disease 2019: a single-centered, retrospective, observational study. J Public Health (Berl.) (2020). https://doi.org/10.1007/s10389-020-01291-2 | CRP | Mortality vs. survived |

### S4 Figures Risk of bias assessment

Q1… What is the risk of bias that the sample size was too small to be representative for the relevant population (Covid-19 patients)?

Q2… What is the chance of applicability that the sample within the screened patients was a true or close representation of the target population?

Q3… What was the patient selection?

Q4… What is the risk of bias of the sampling procedure?

Q5… What is the risk that there was a selection bias?

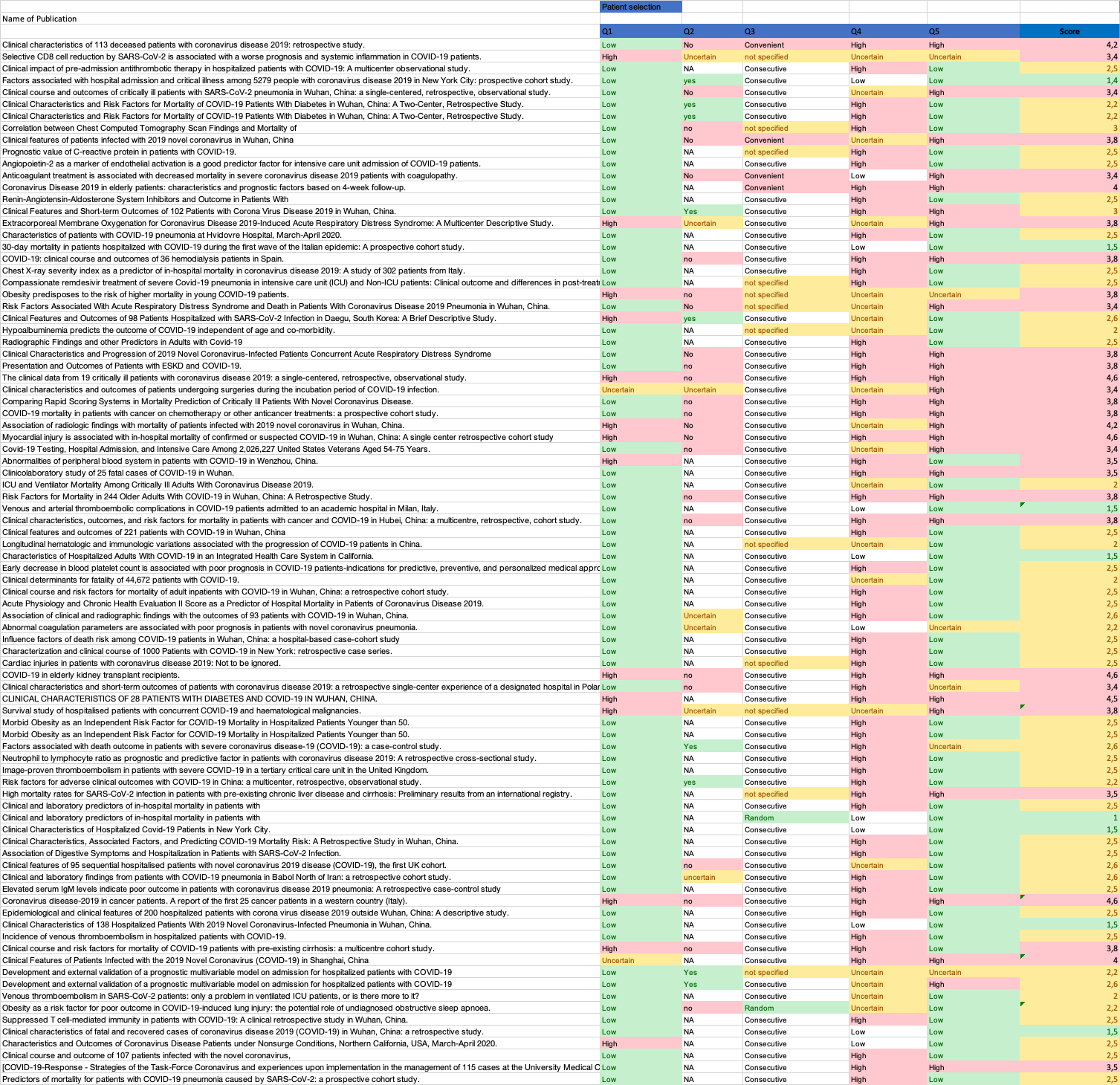

Figure 1 - Risk of bias assessment for 'Patient selection'

Q6… Were there inappropriate exclusions?

Q7… Does the study report number screen, number enrolled, and number assessed?

Q8… Are the patients reported that are excluded for a reported reason?

Q9… What is the risk of incomplete outcome data?

Q10… Were data collected directly form the patients (as opposed to a registry)?

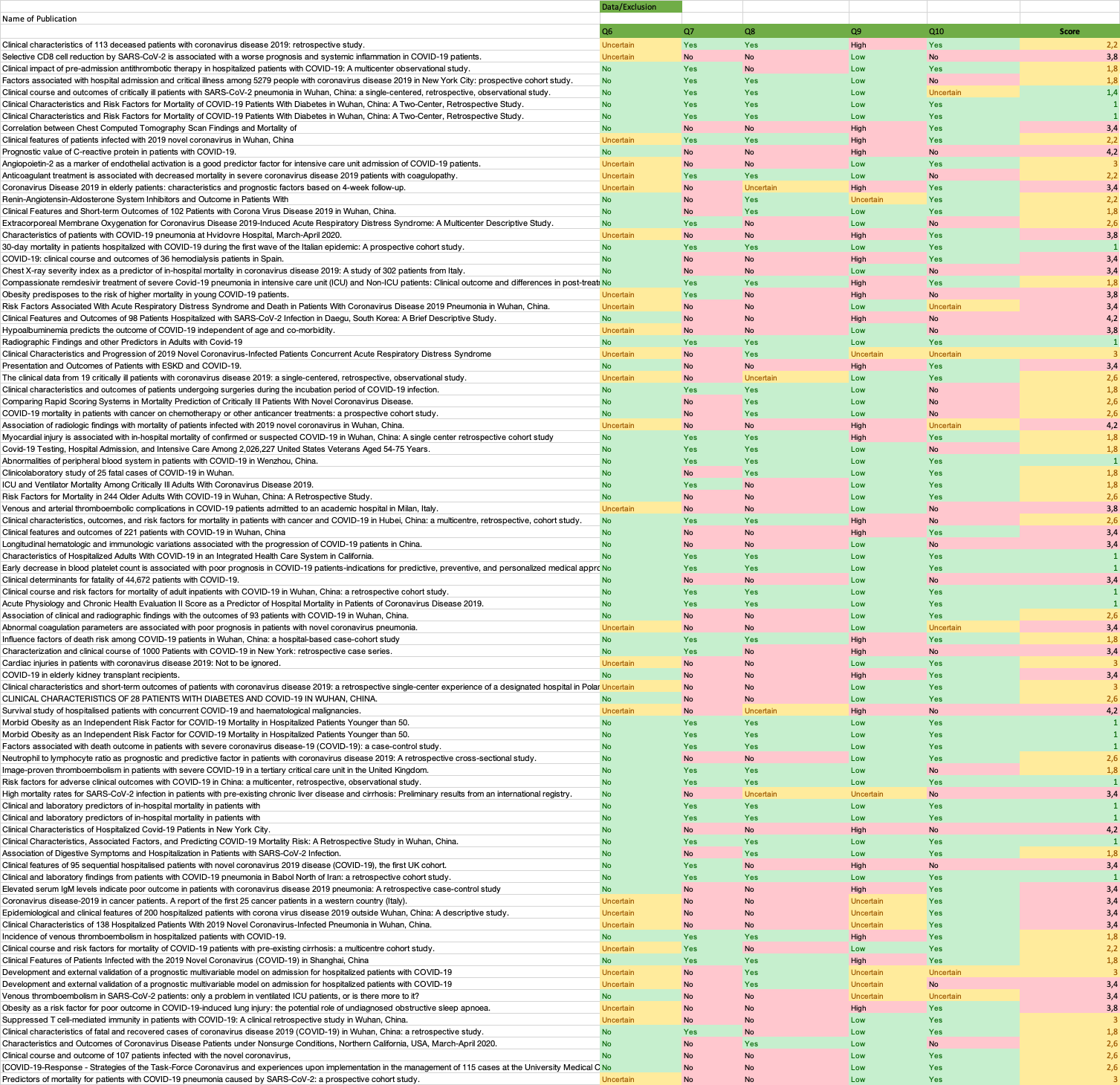

Figure 2 - Risk of bias assessment for 'Data availability and Exclusions'

Q11… What is the risk of applicability of the case definition used in this study?

Q12… Was the same case definition used on all patients?

Q13… Were definitions severe/critical as proposed by the WHO proposition?

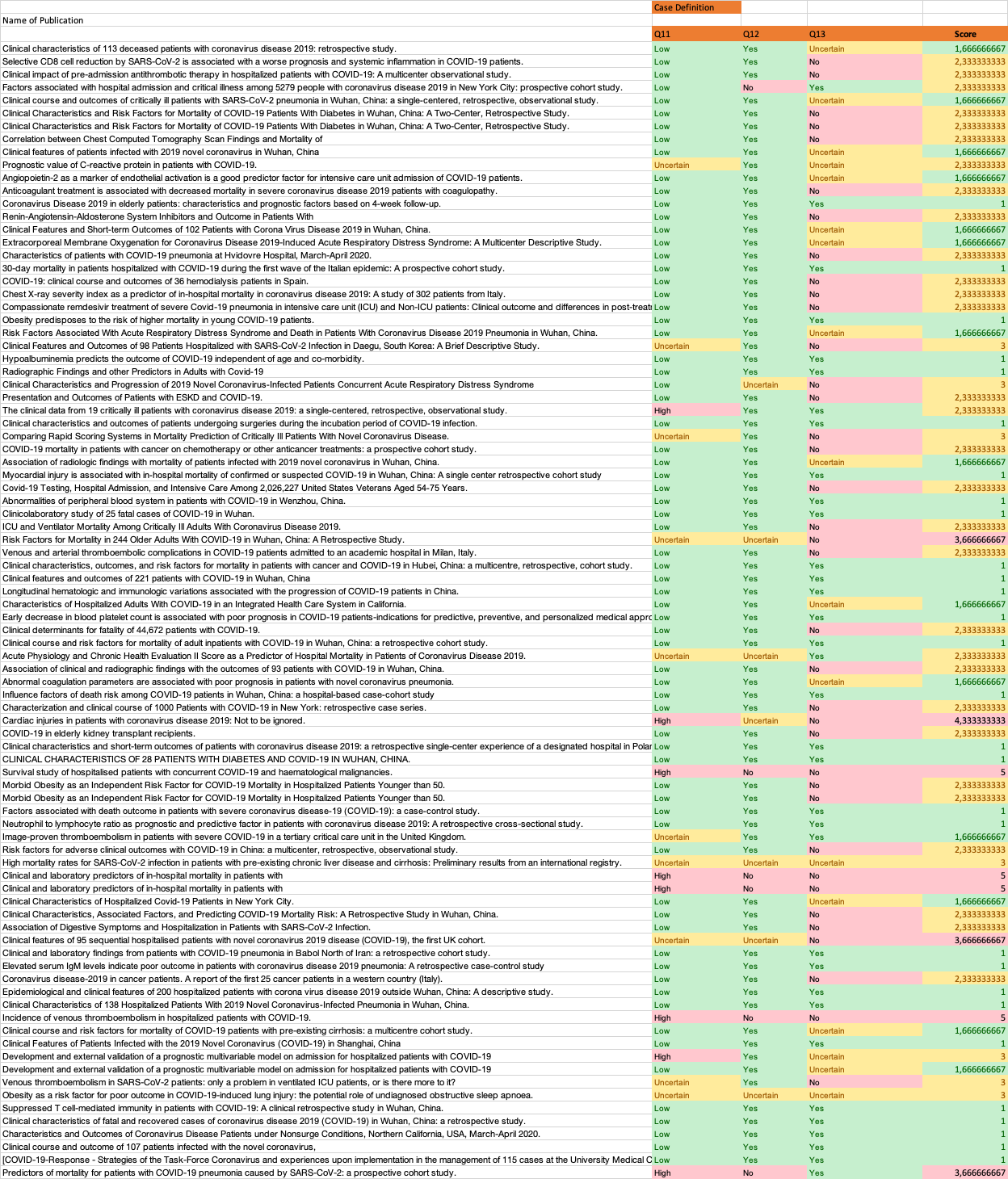

Figure 3 - Risk of bias assessment for 'Covid- and severity case definition'

Supplementary Table 1 - Risk of bias assessment for ‘Patient selection and chance of applicability'

|  | Low Risk | Intermediate Risk | High Risk |
| --- | --- | --- | --- |
| Q1 – Representative Sample Size | n>30 |  | n<30 |
| Q2 – Selection of screened patients representative | Multicenter study | Singlecenter study with n>30 | Singlecenter study n<30 |
| Q3 – Patient selection | randomized | consecutive | convenient |
| Q4 – Risk of bias of sampling procedure | randomized |  | Non randomized |
| Q5 – Risk of selection bias | No subgroup or special treatment | Specialized treatment, e.g. drug-trial | Specialized subgroup, e.g. cancer patients, transplant recipients, dialysis, etc. |

Supplementary Table 2 - Risk of bias assessment for ‘Patient data availability and exclusions’

|  | Low Risk | Intermediate Risk | High Risk |
| --- | --- | --- | --- |
| Q6 – Inappropriate Exclusions | All exclusion criterias listed |  | No exclusion criterias listed |
| Q7 – Numbers Reported | Screened, enrolled, analyzed listed | Only two listed | Non/one listed |
| Q8 – Exclusions reported | All exclusions reported with cause of exclusion | Only numbers reported | No exclusions reported |
| Q9 – Risk of incomplete outcome data | All patients have a definitive outcome |  | Some are still in hospital |
| Q10 – Data directly from patients | From patient or patient file |  | From a registry |

Supplementary Table 3 - Risk of bias assessment for 'Covid-19 and severity case definition'

|  | Low Risk | Intermediate Risk | High Risk |
| --- | --- | --- | --- |
| Q11 – COVID-19 Case definition | PCR Test | Combination from  - Symptom screen  - CT scan | Only  Symptom screen or CT scan |
| Q12 – Definiton used for all patients | yes |  | no |
| Q13 – Severity definitions as proposed by WHO | yes | Local guidelines used | None used |

### S5 Forest Plots for difference of medians for mortality vs. survived across different indicators

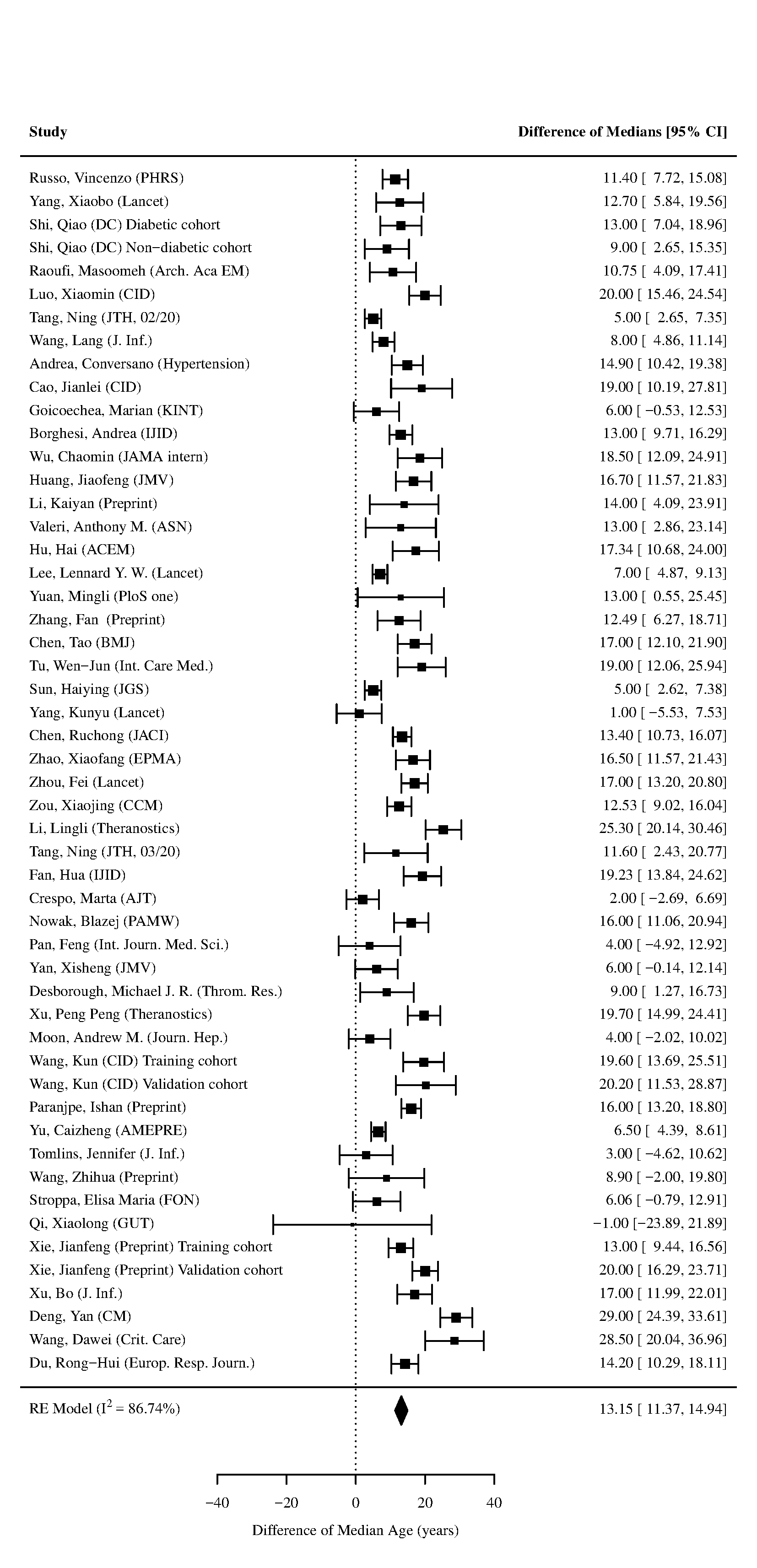

Figure 4 - Forest plot for difference of medians of **Age** in those who died vs. survived

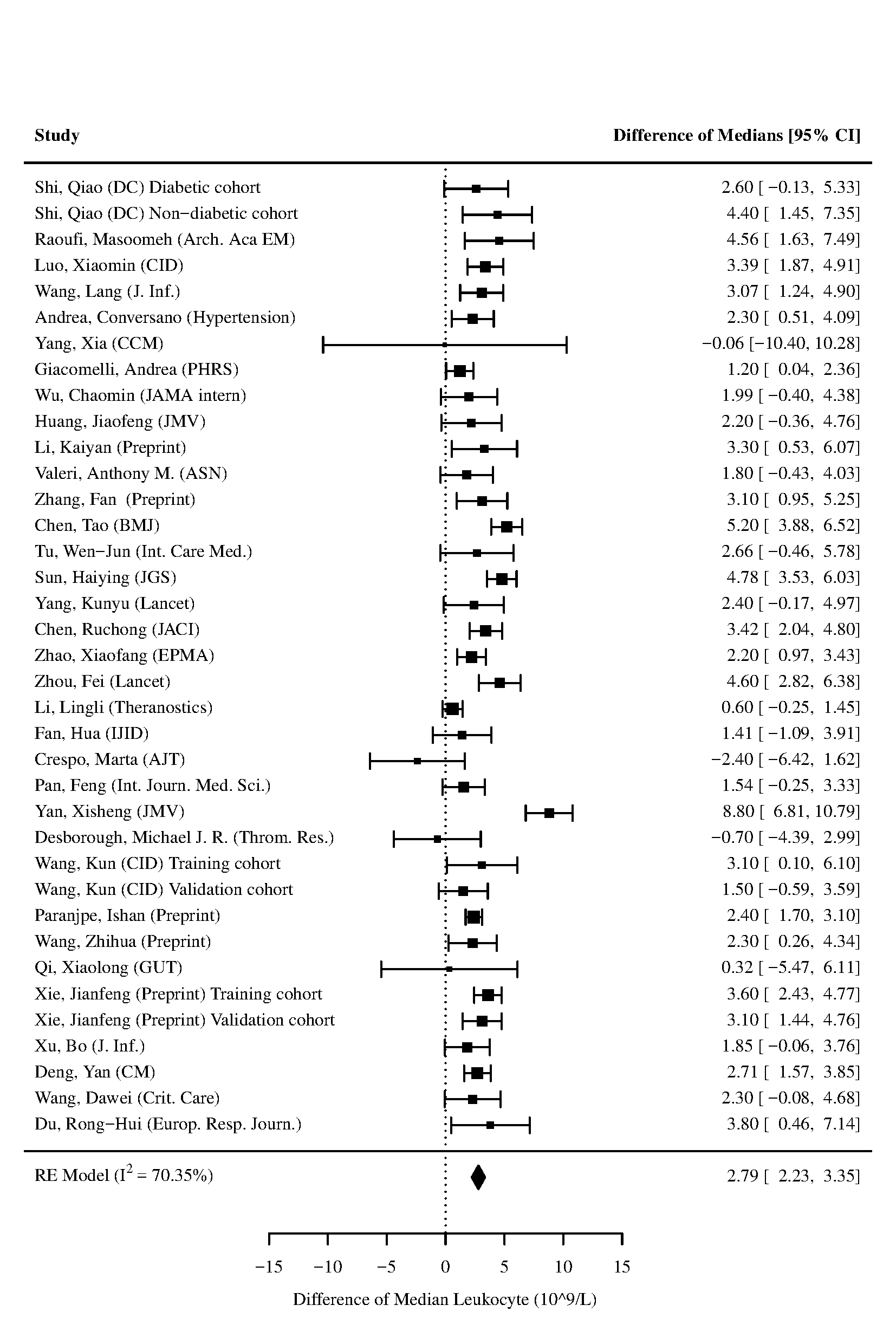

Figure 5 - Forest plot for difference of medians of **Leukocyte** in those who died vs. survived

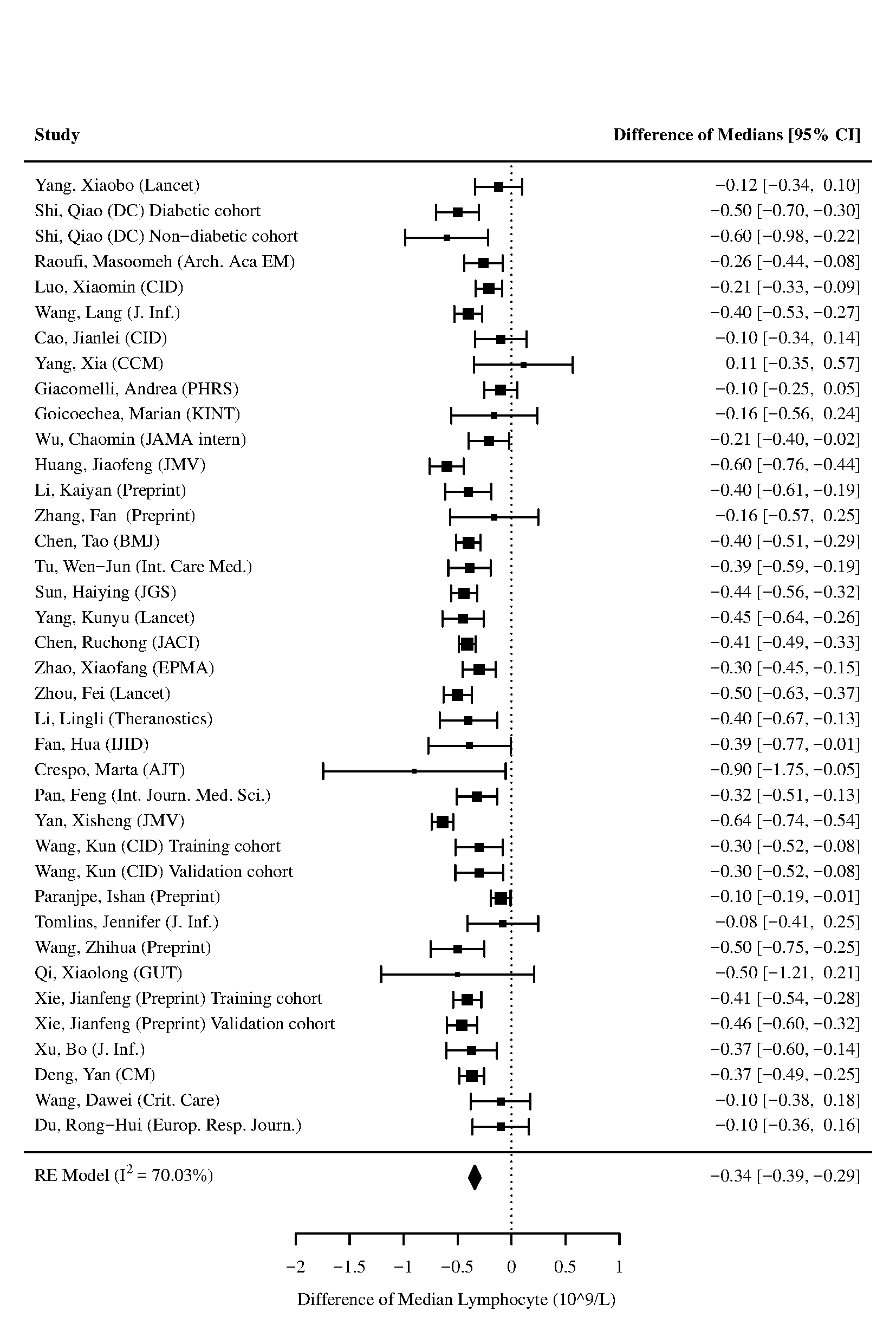

Figure 6 - Forest plot for difference of medians of **Lymphocyte** in those who died vs. survived

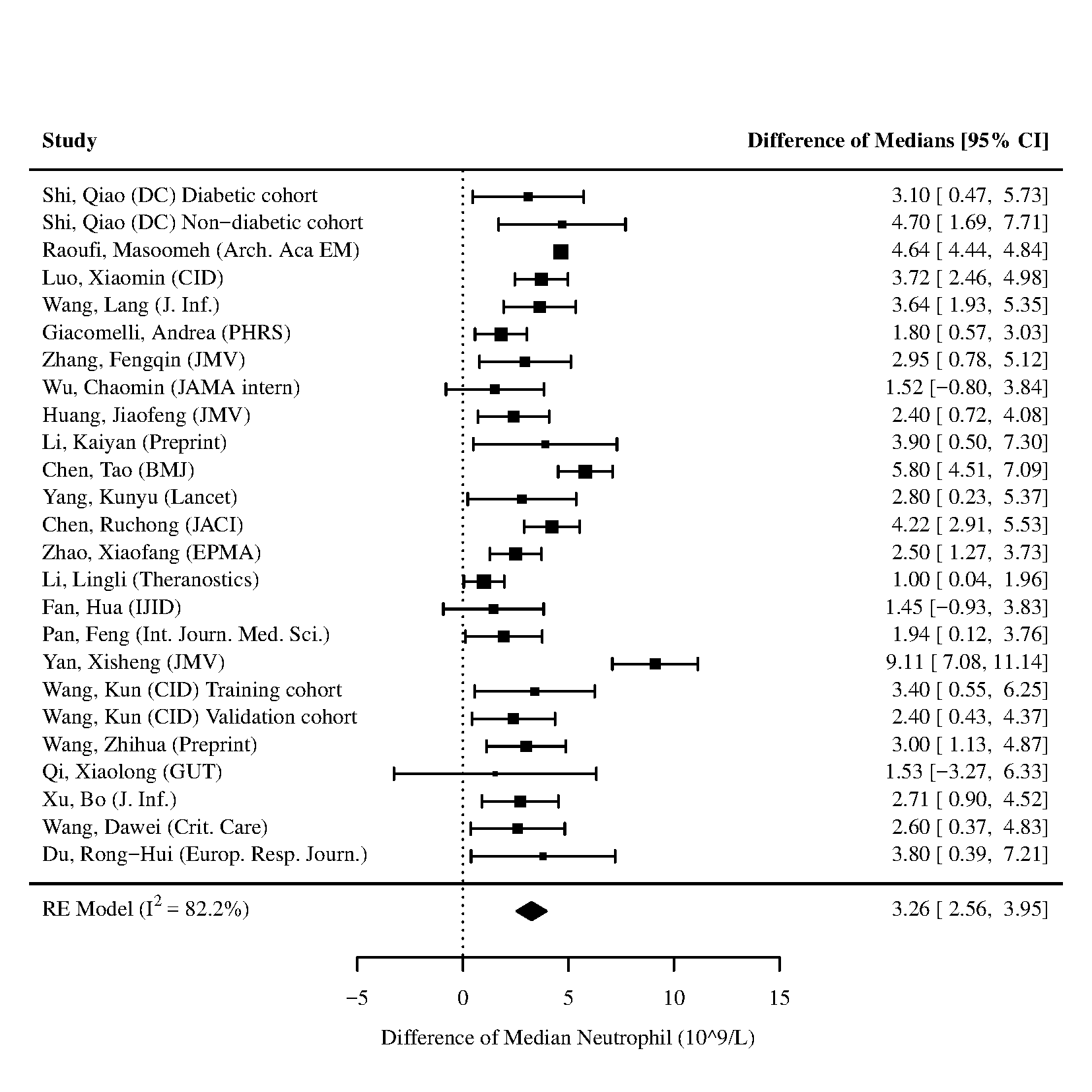

Figure 7 - Forest plot for difference of medians of **Neutrophils** in those who died vs. survived

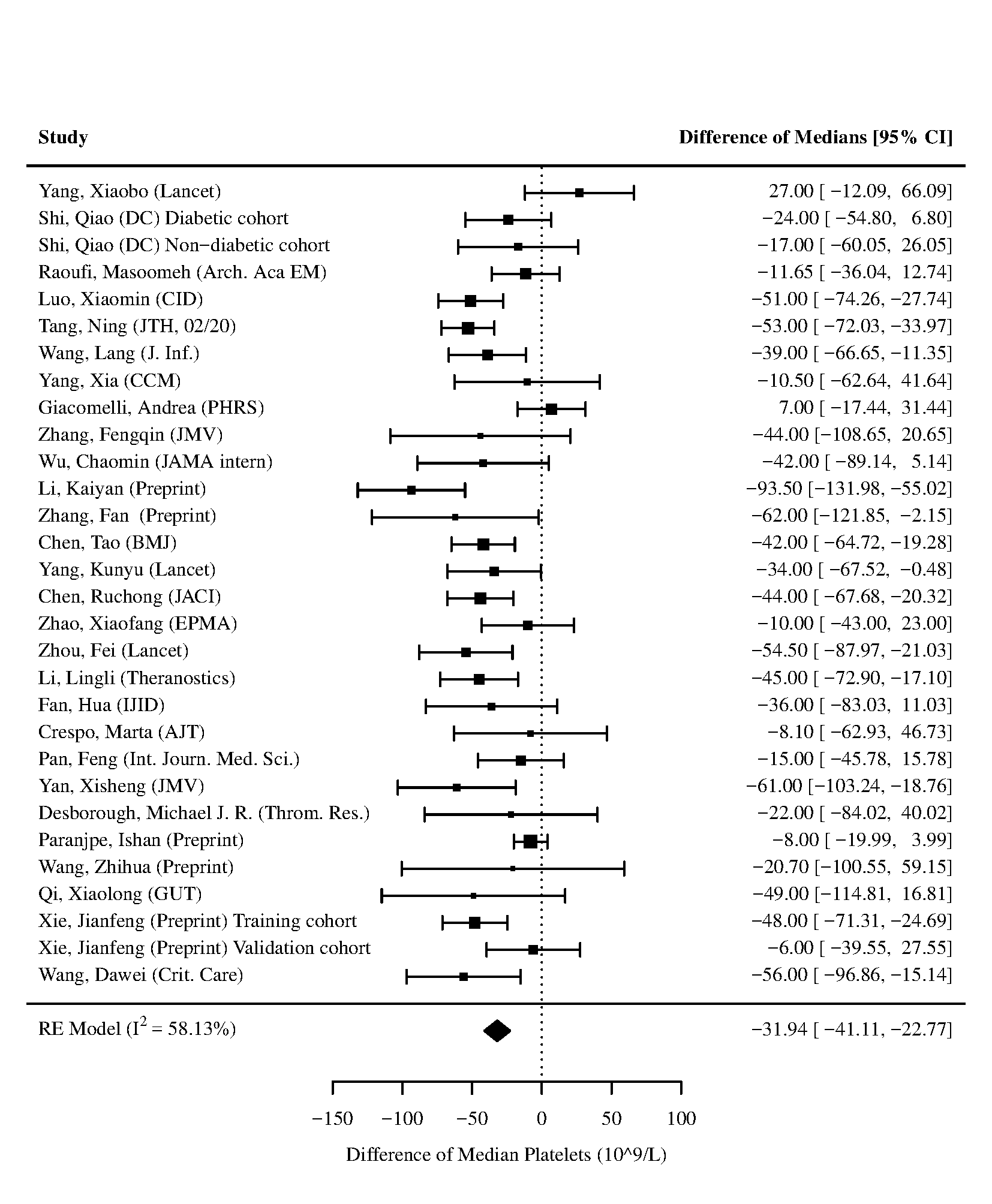

Figure 8 - Forest plot for difference of medians of **Platelets** in those who died vs. survived

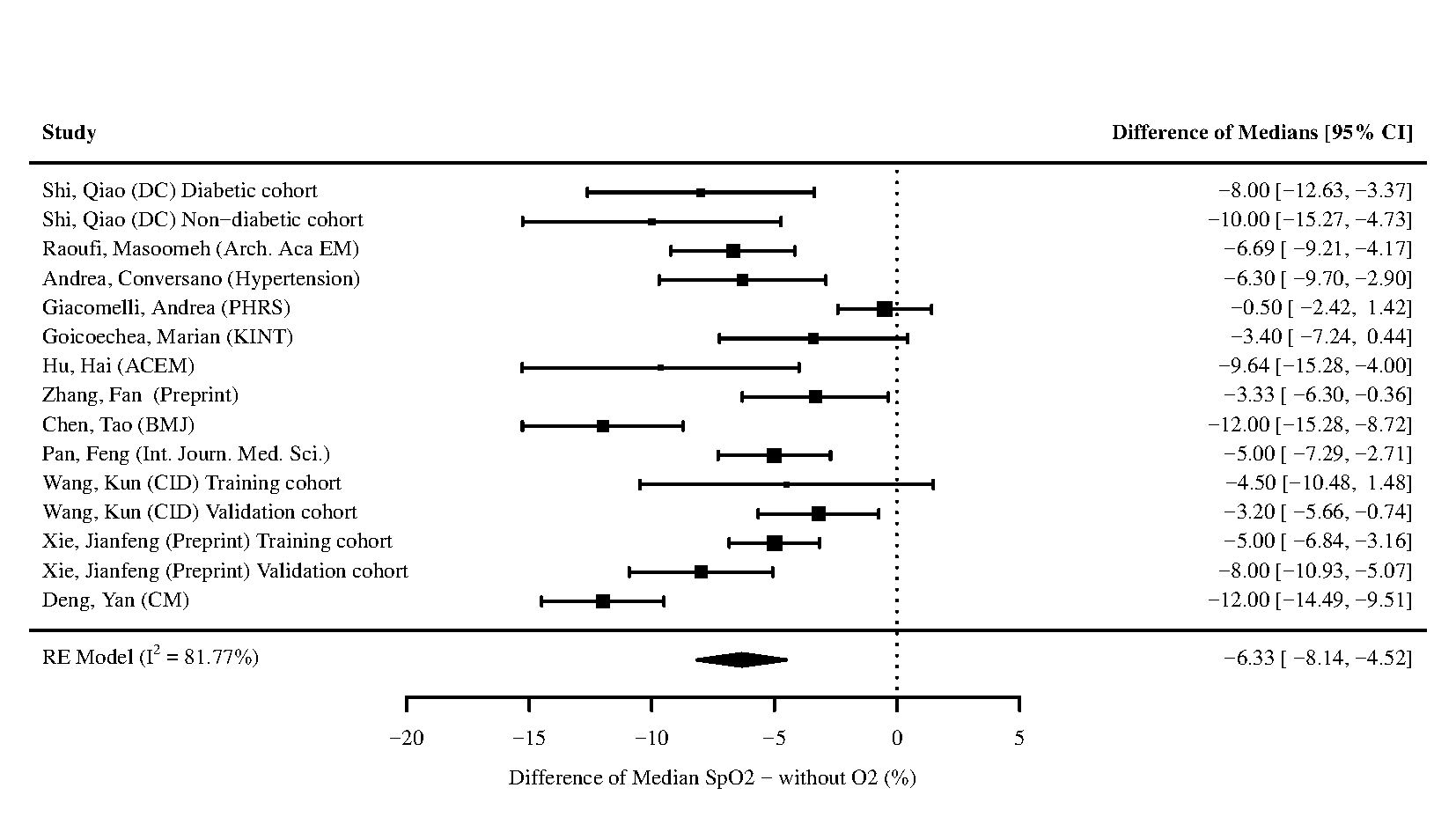

Figure 9 - Forest plot for difference of medians of **Oxygen Saturation (SpO2)** without oxygen (O2) in those who died vs. survived

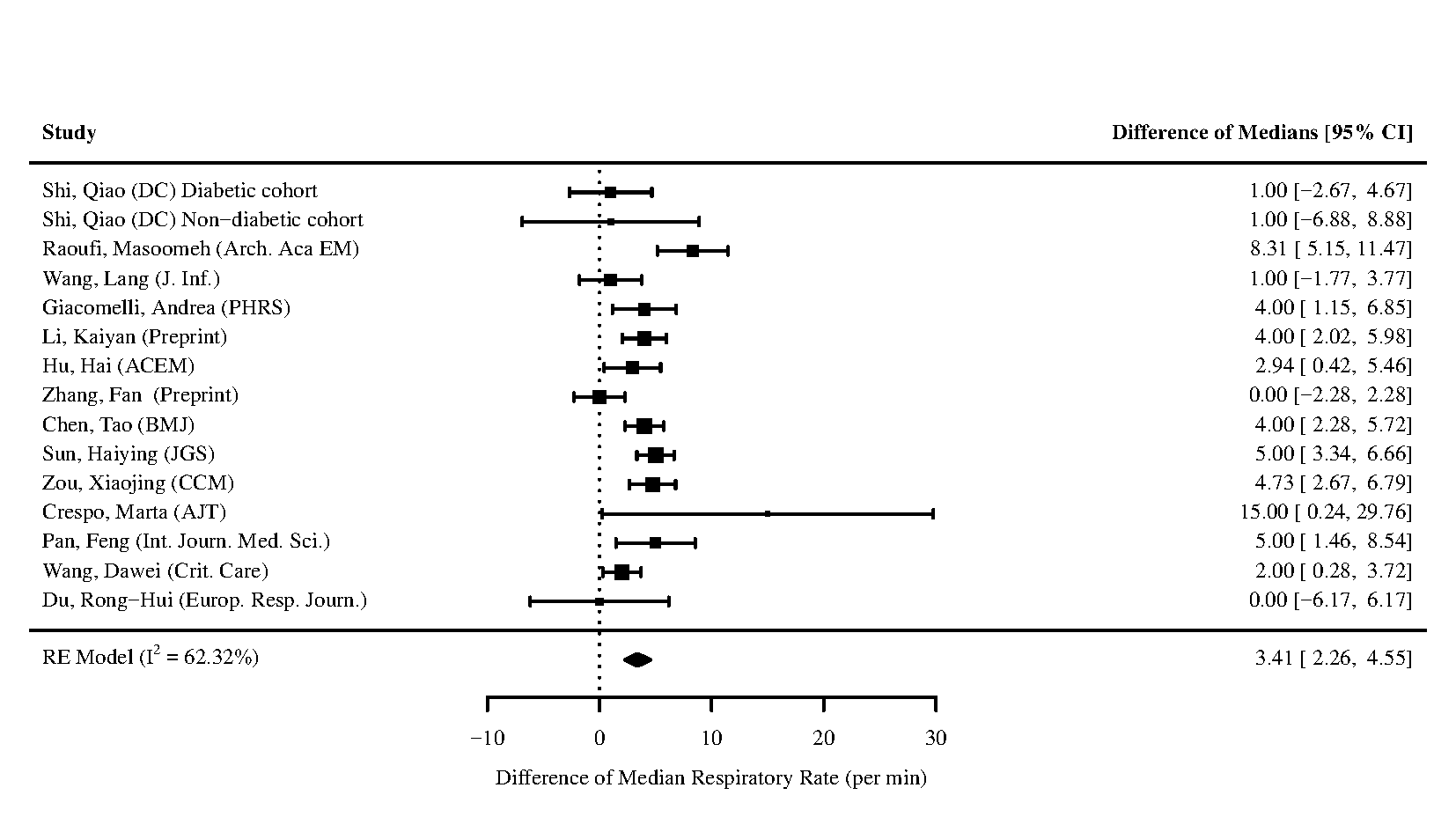

Figure 10 - Forest plot for difference of medians of **Respiratory Rate** in those who died vs. survived

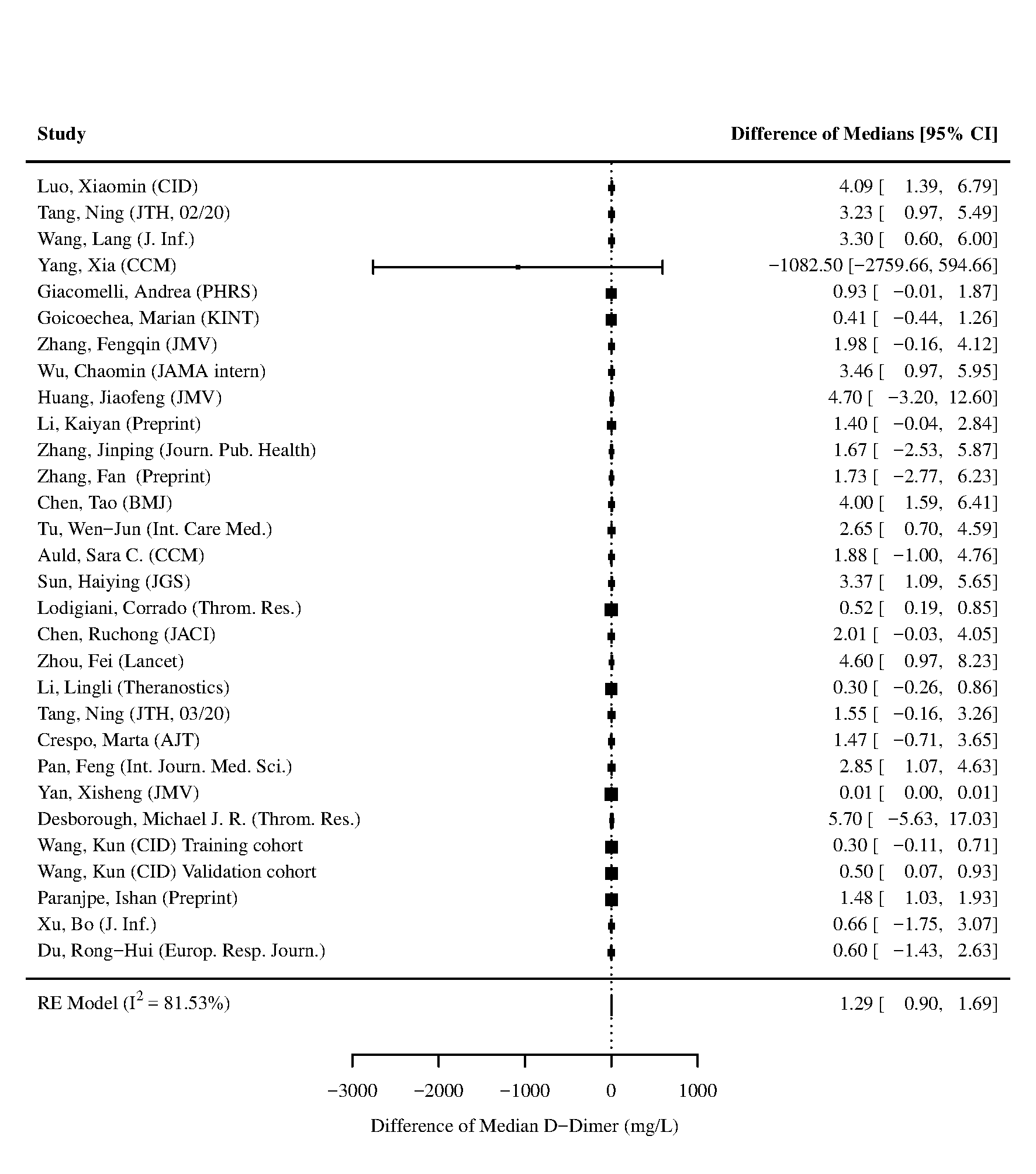

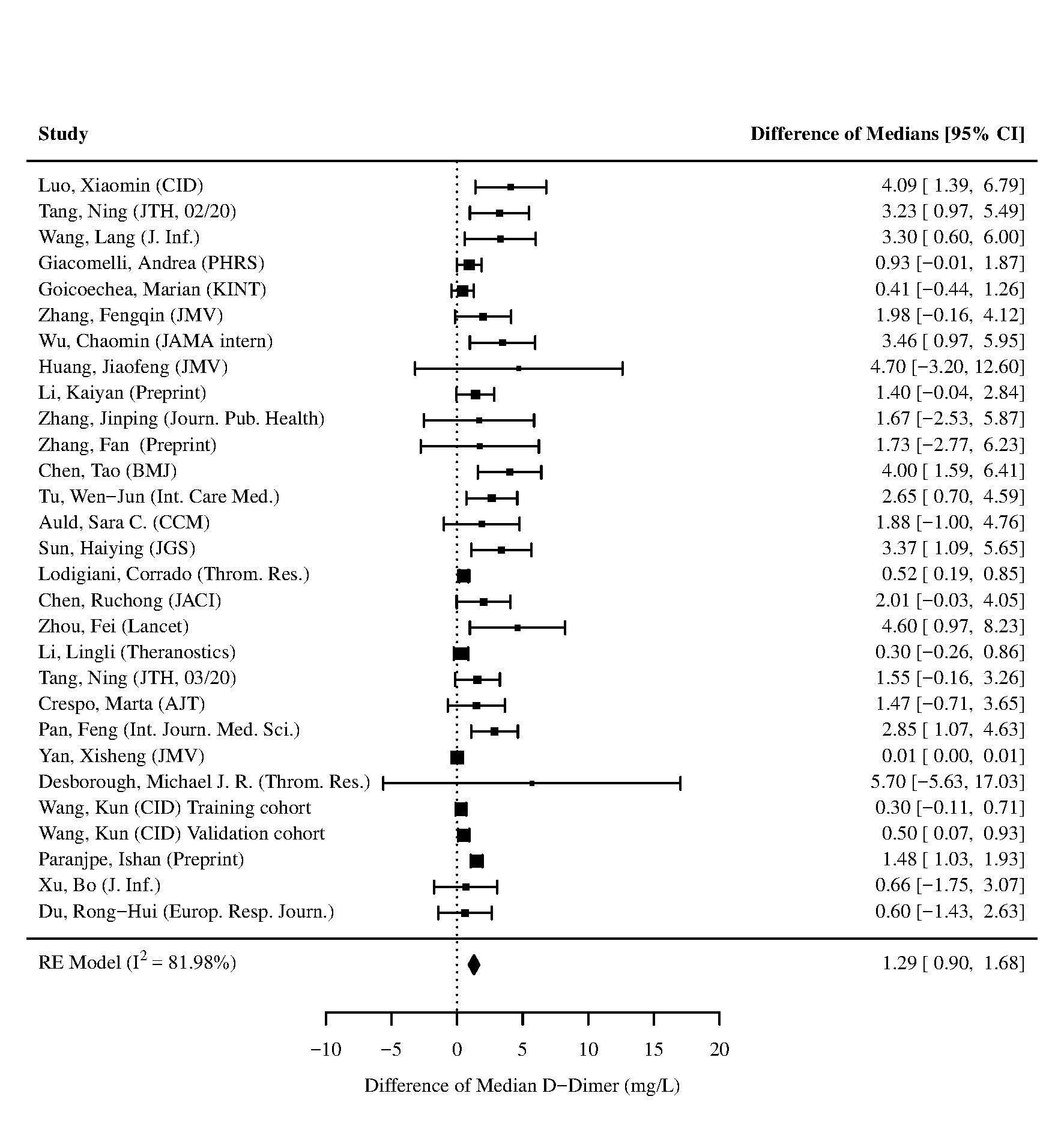

Figure 11 - Forest plot for difference of medians of **D-Dimer** in those who died vs. survived. The study by Yang et al. used a different laboratory assay. The left panel contains the forest plot of the analysis including the study by Yang et al., and the right panel contains the forest plot of the analysis excluding the study by Yang et al.

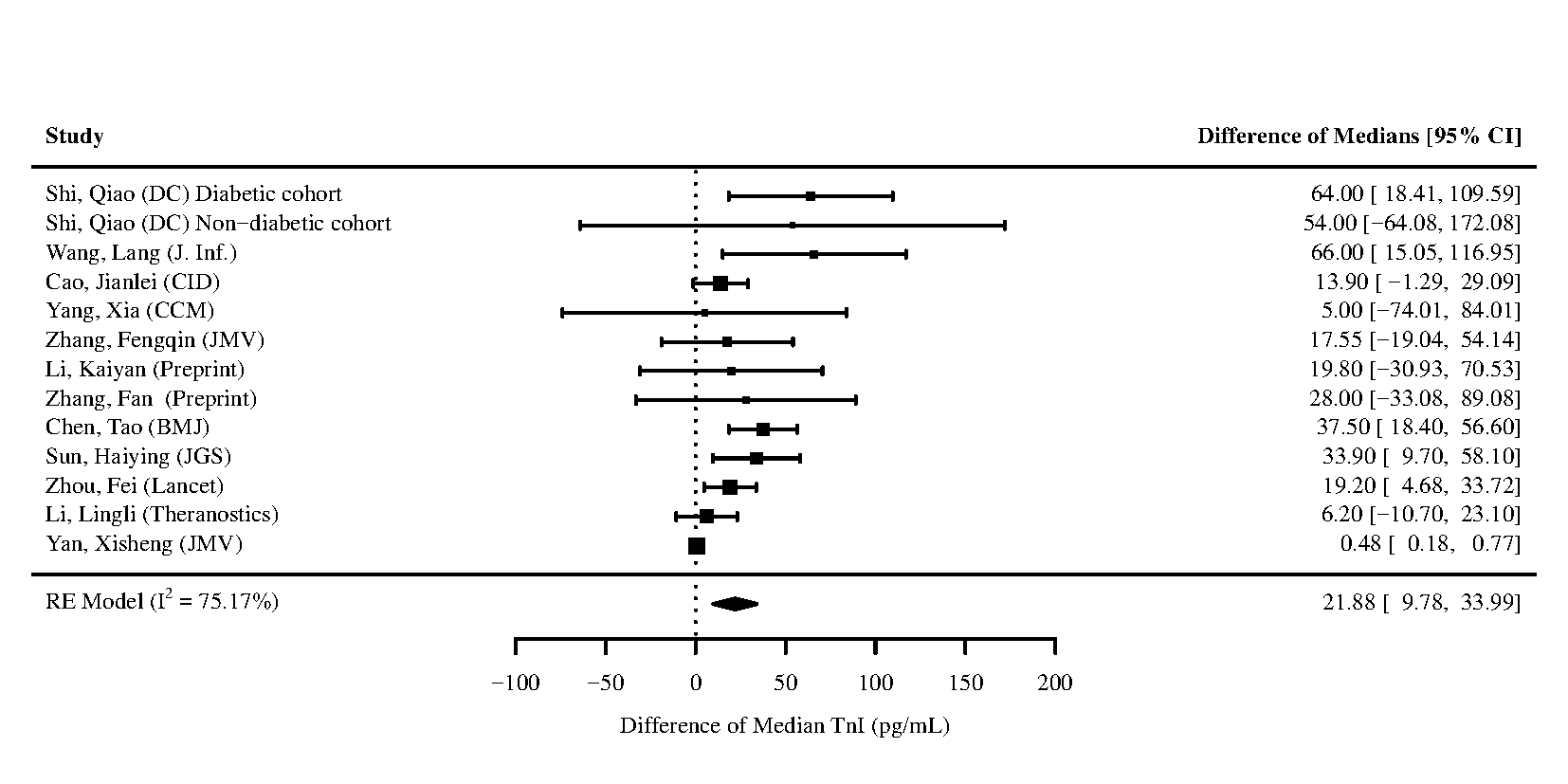

Figure 12 - Forest plot for difference of medians of **Troponin I (TnI)** in those who died vs. survived

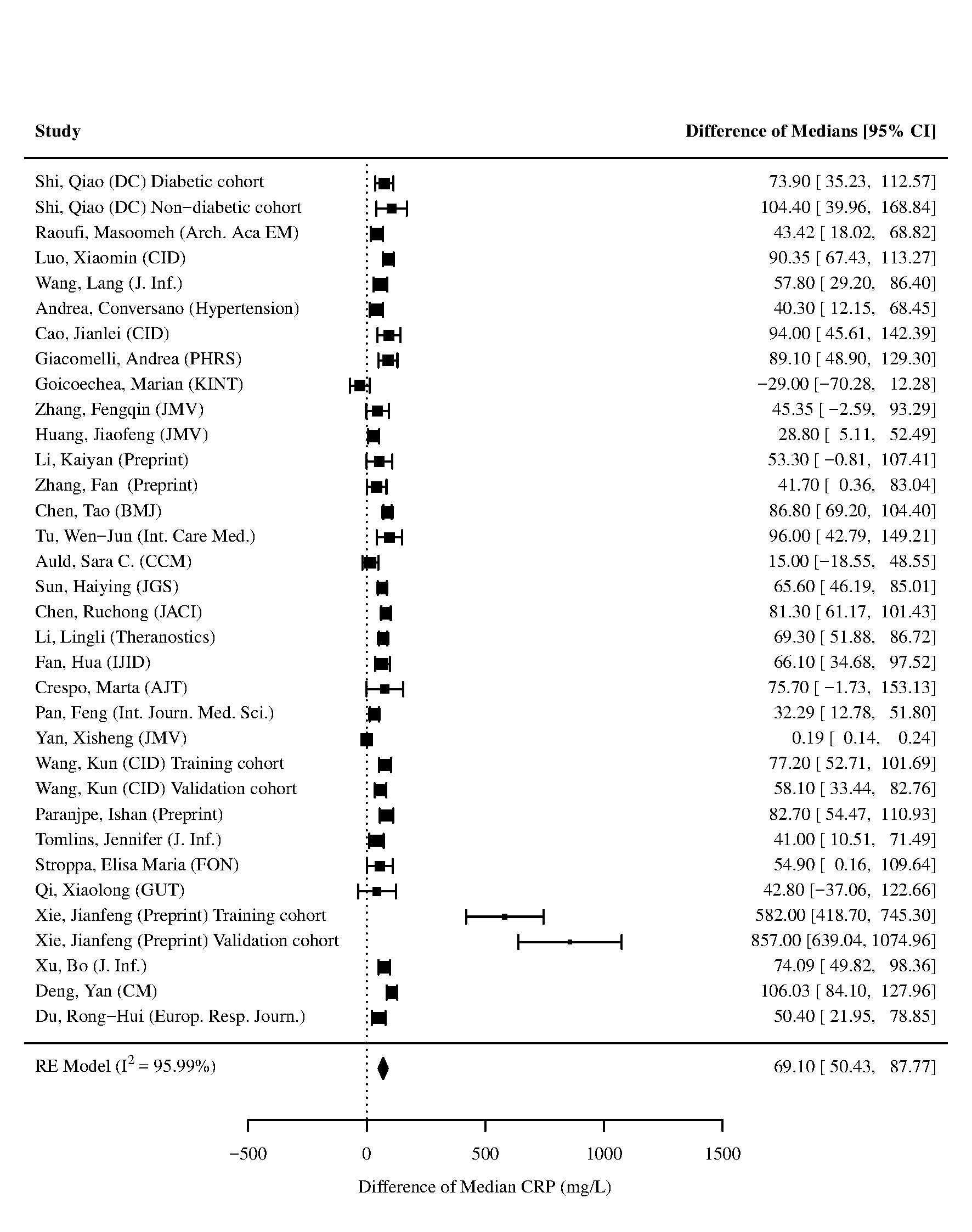

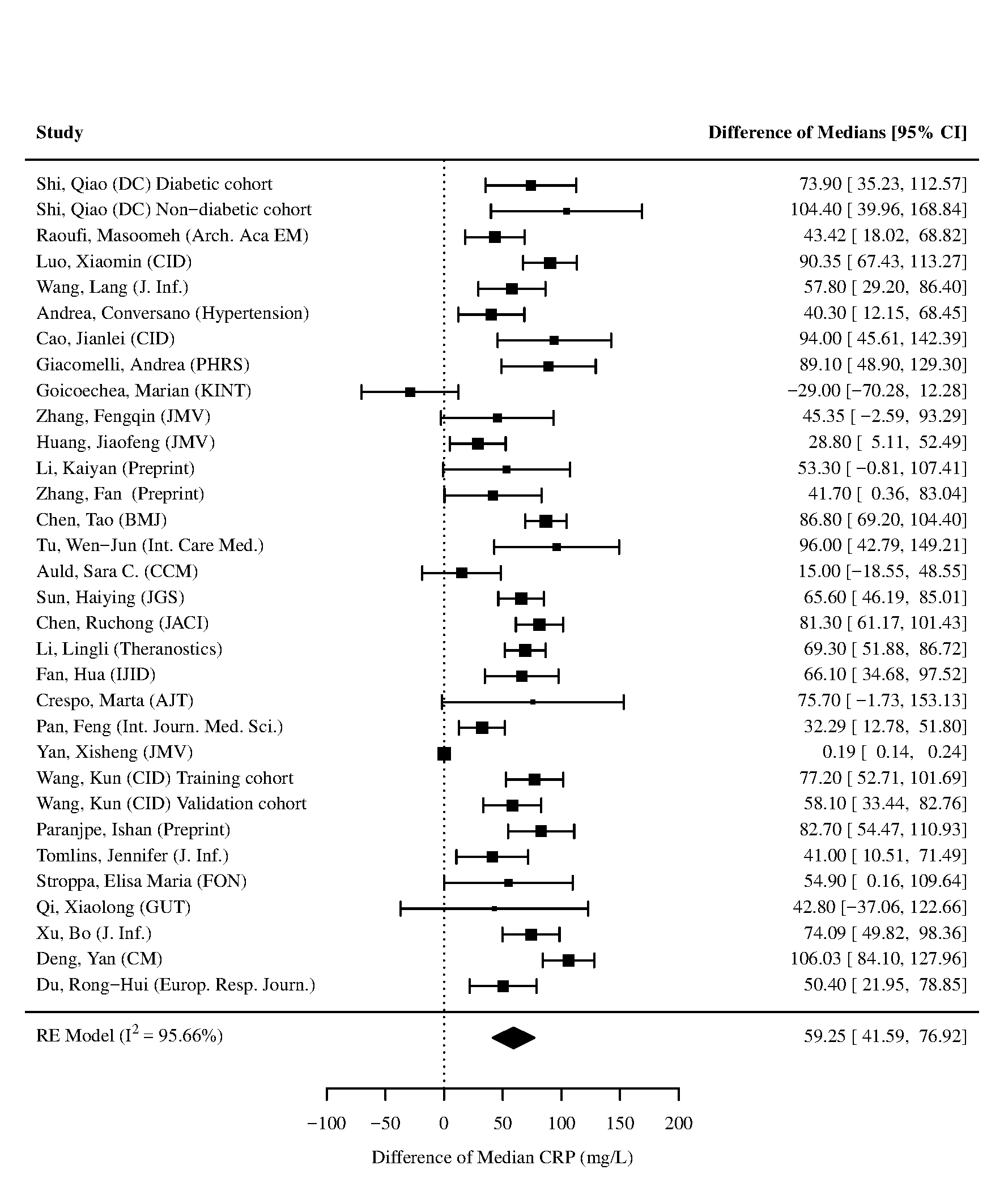

Figure 13 - Forest plot for difference of medians of **C-reactive protein (CRP)** in those who died vs. survived. Xie et al. developed a mortality prediction tool and had substantial higher values of CRP than other studies. The left panel contains the forest plot for the analysis including the study by Xie et al., and the right panel contains the forest plot for the analysis excluding the study by Xie et al.

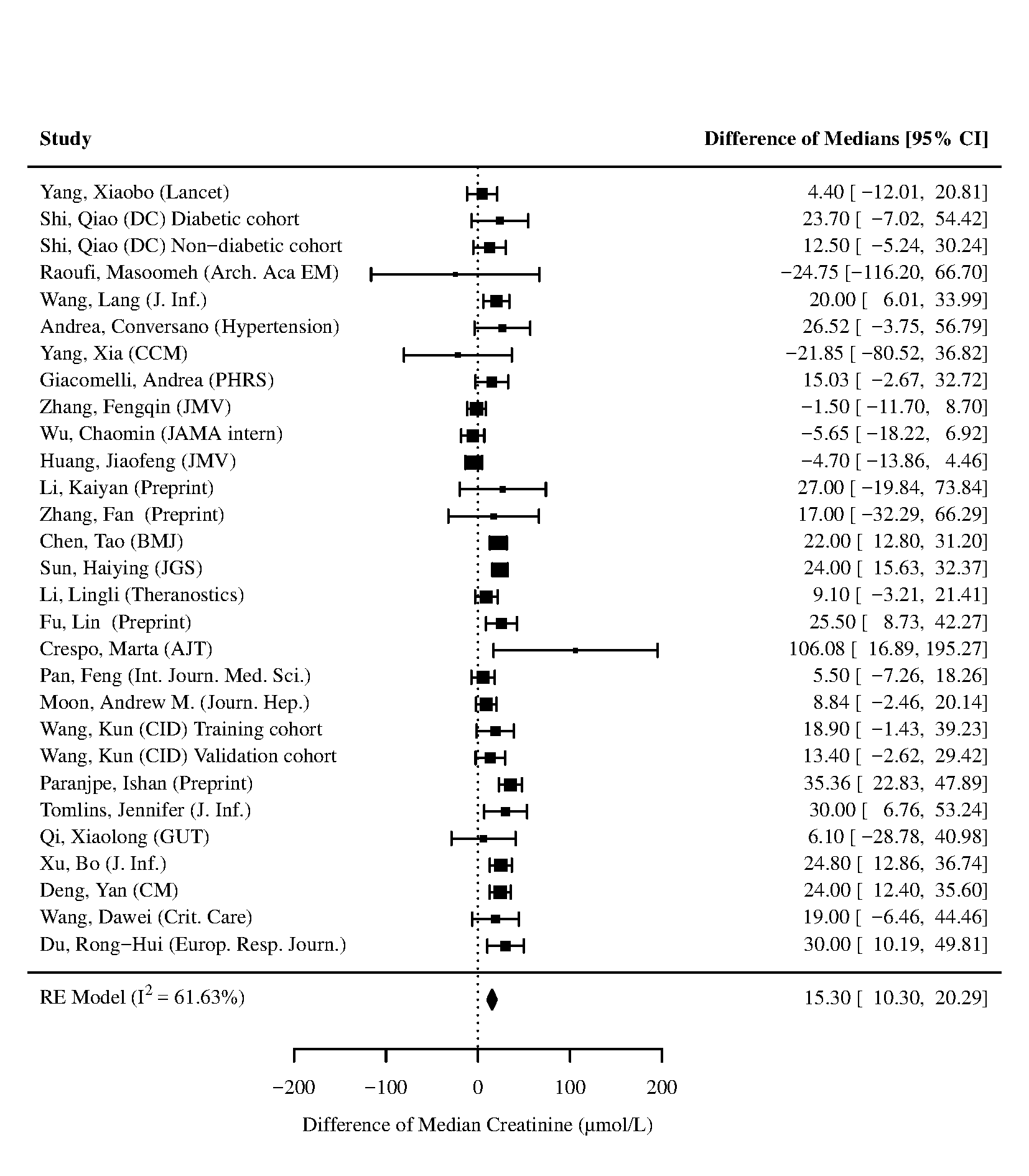

Figure 14 - Forest plot for difference of medians of **Creatinine** in those who died vs. survived

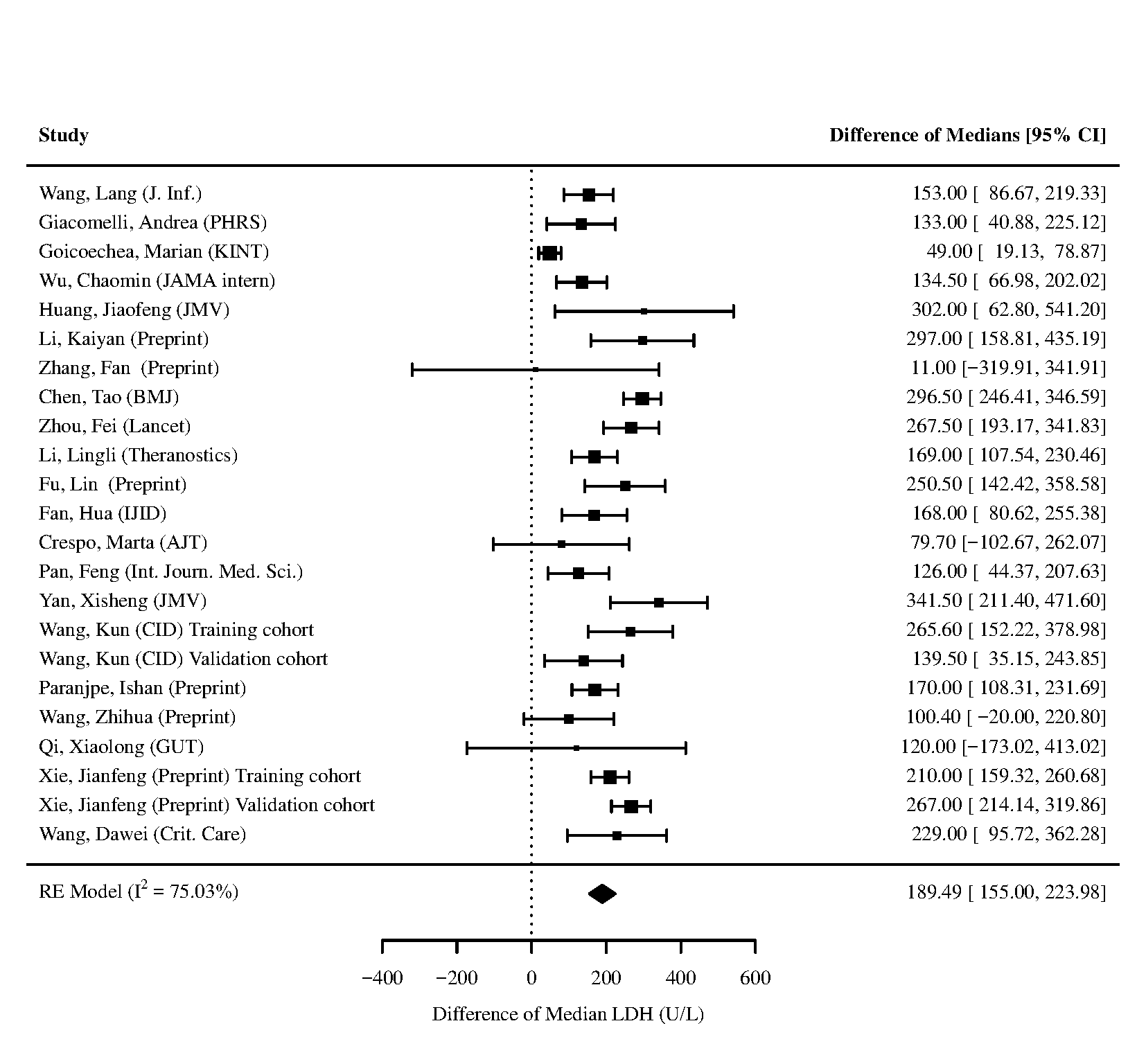

Figure 15 - Forest plot for difference of medians of **Lactate Dehydrogenase (LDH)** in those who died vs. survived

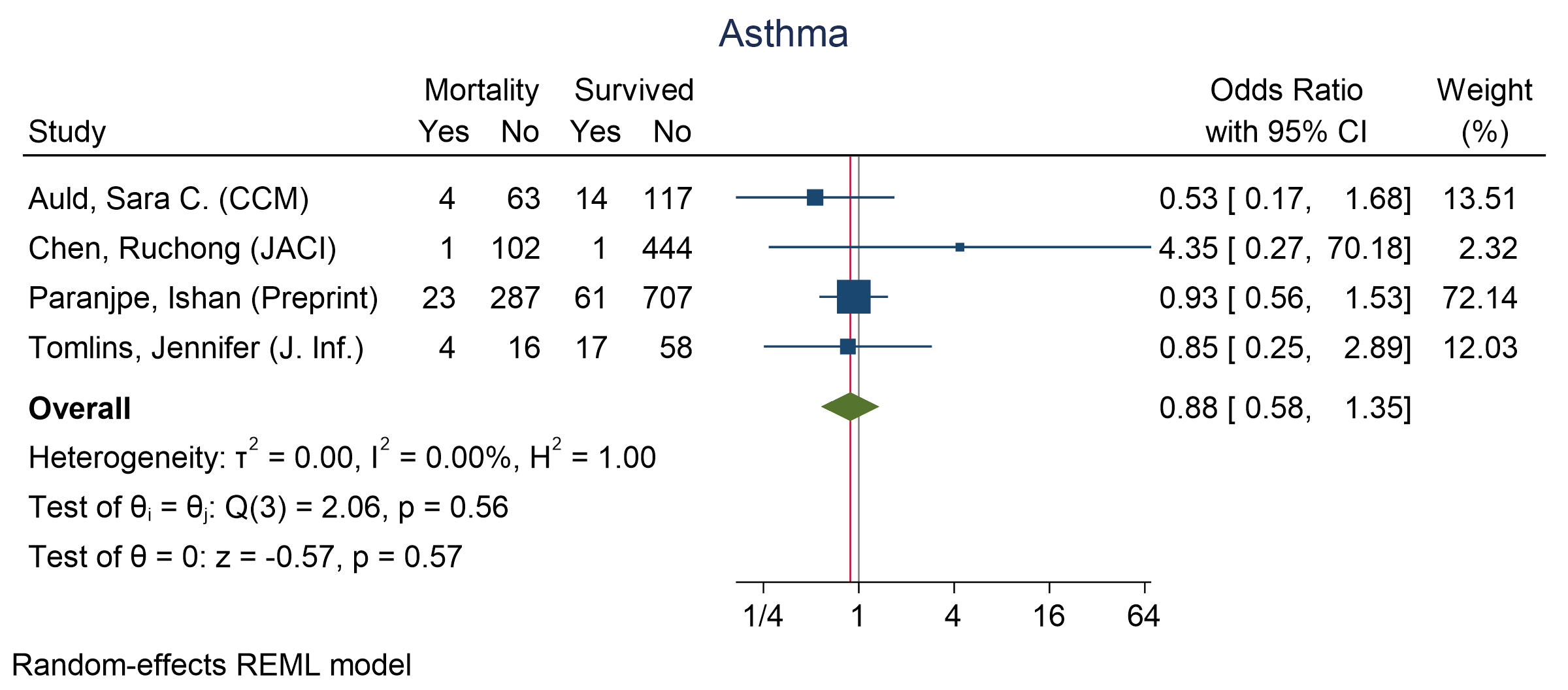

Figure 16 - Forest plot for Odds Ratio and 95% Confidence Interval (CI) of **Asthma** for those who died vs. survived

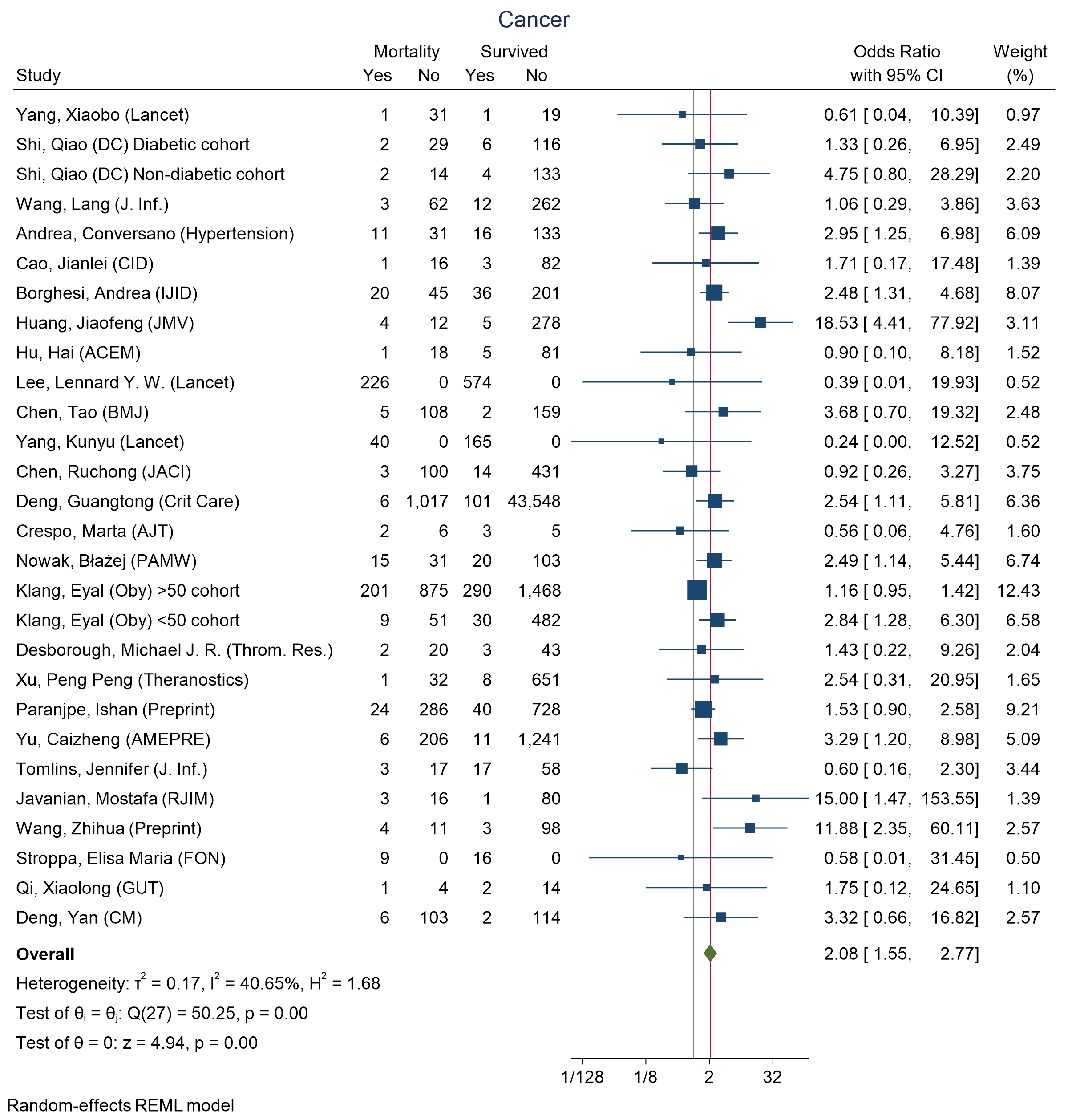

Figure 17 - Forest plot for Odds Ratio and 95% Confidence Interval (CI) of **Cancer** for those who died vs. survived

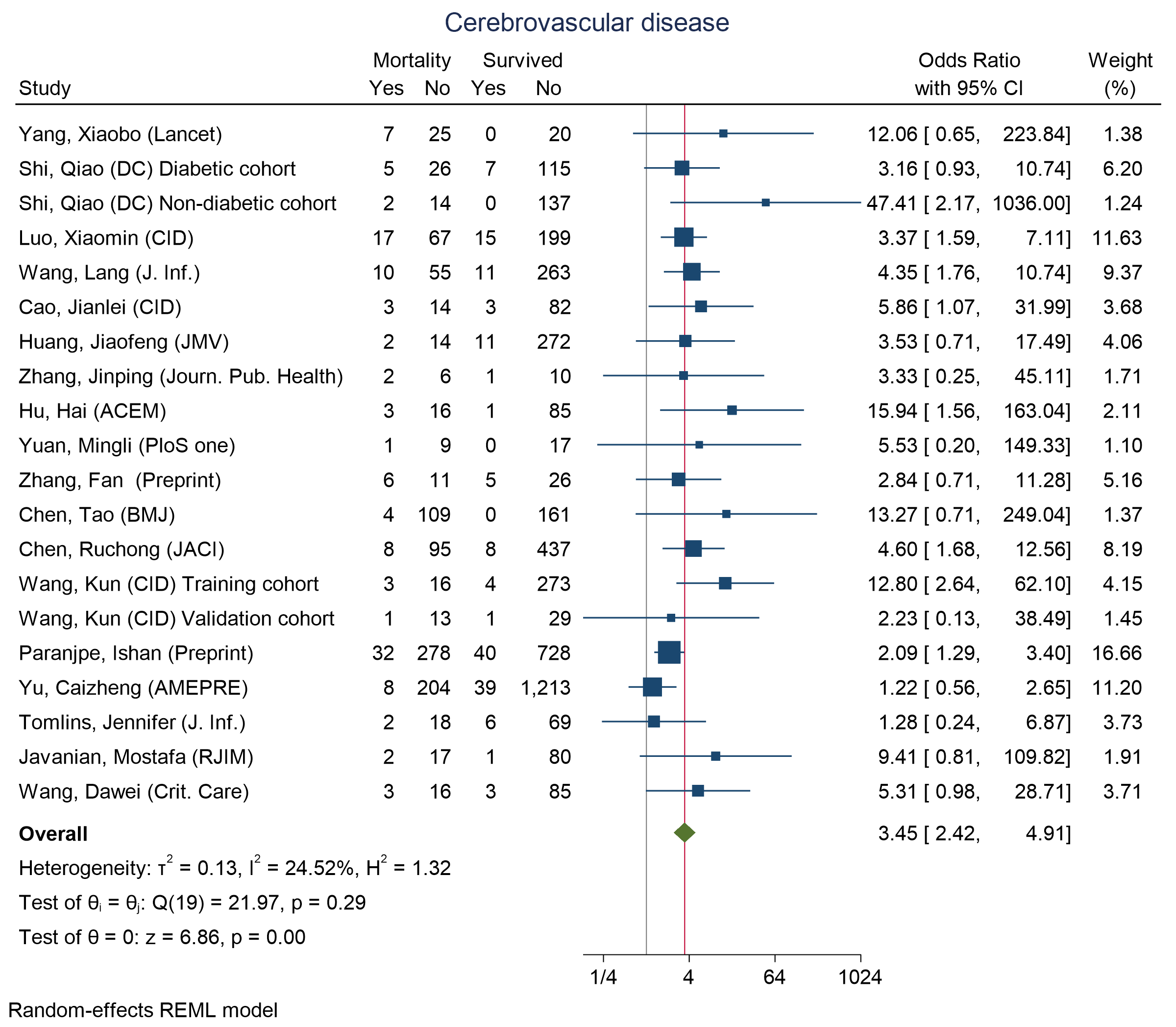

Figure 18 - Forest plot for Odds Ratio and 95% Confidence Interval (CI) of **Cerebrovascular disease** for those who died vs. survived

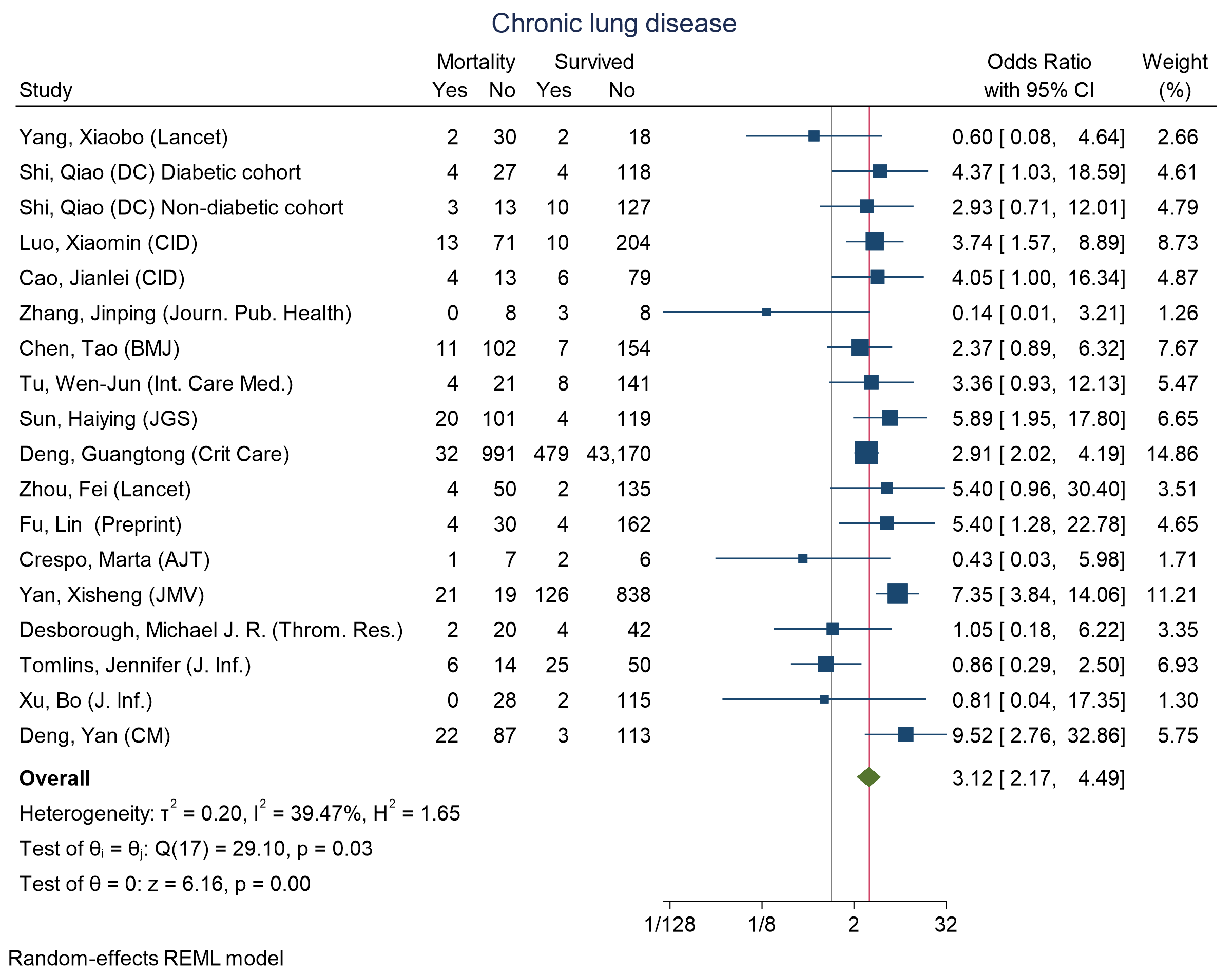

Figure 19 - Forest plot for Odds Ratio and 95% Confidence Interval (CI) of **Chronic lung disease** for those who died vs. survived

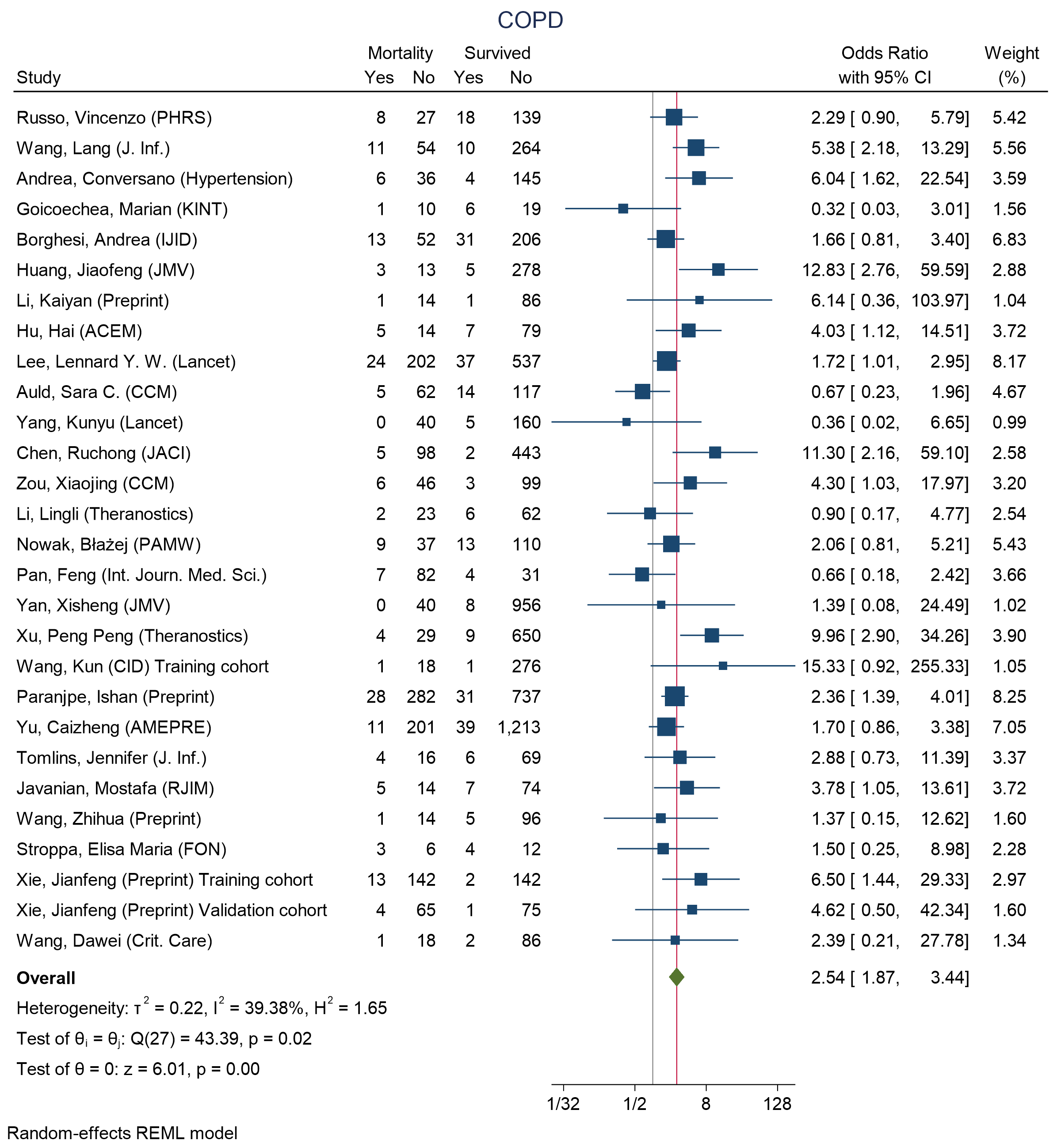

Figure 20 - Forest plot for Odds Ratio and 95% Confidence Interval (CI) of **chronic obstructive pulmonary disease (COPD)** for those who died vs. survived

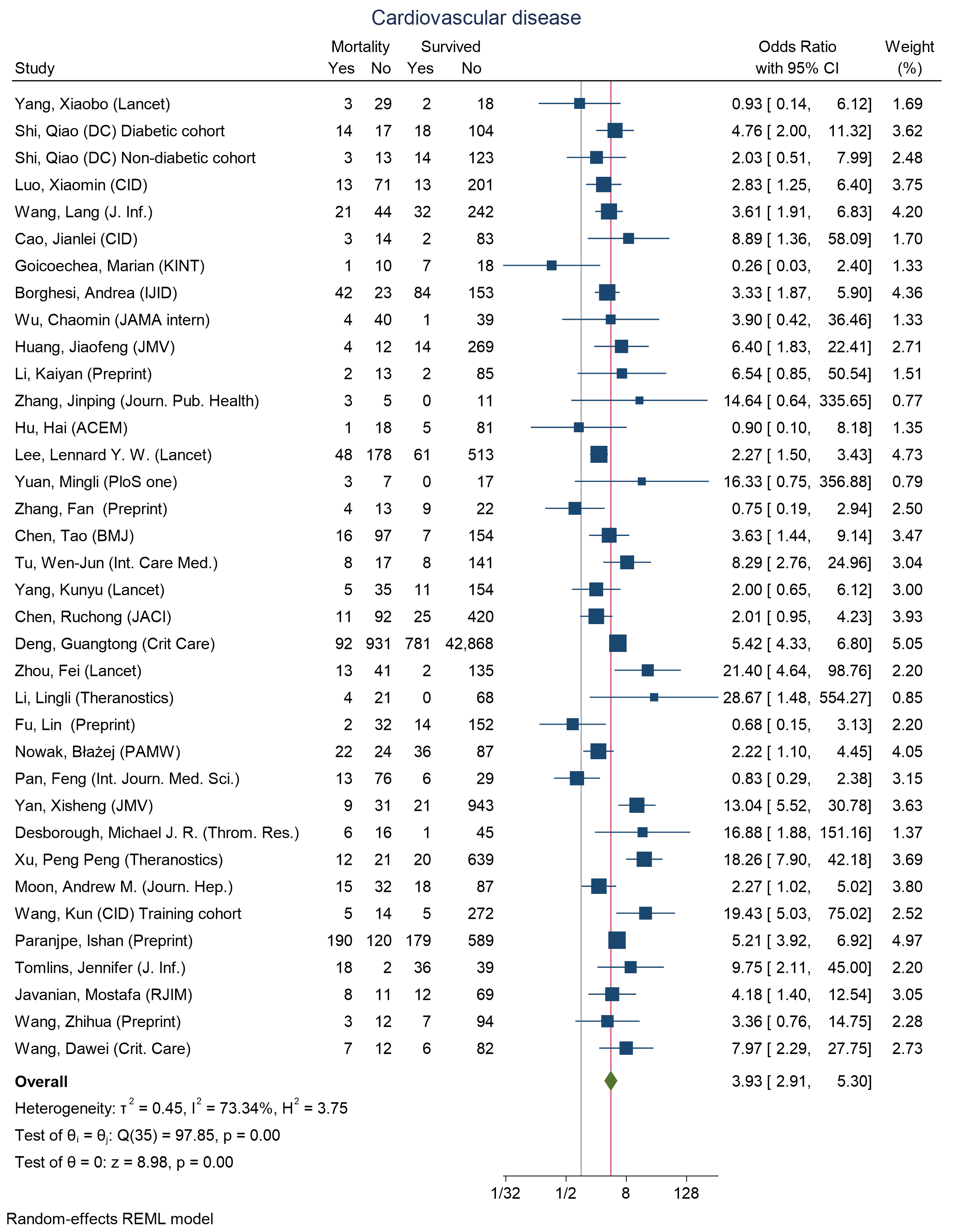

Figure 21 - Forest plot for Odds Ratio and 95% Confidence Interval (CI) of **Cardiovascular disease** for those who died vs. survived

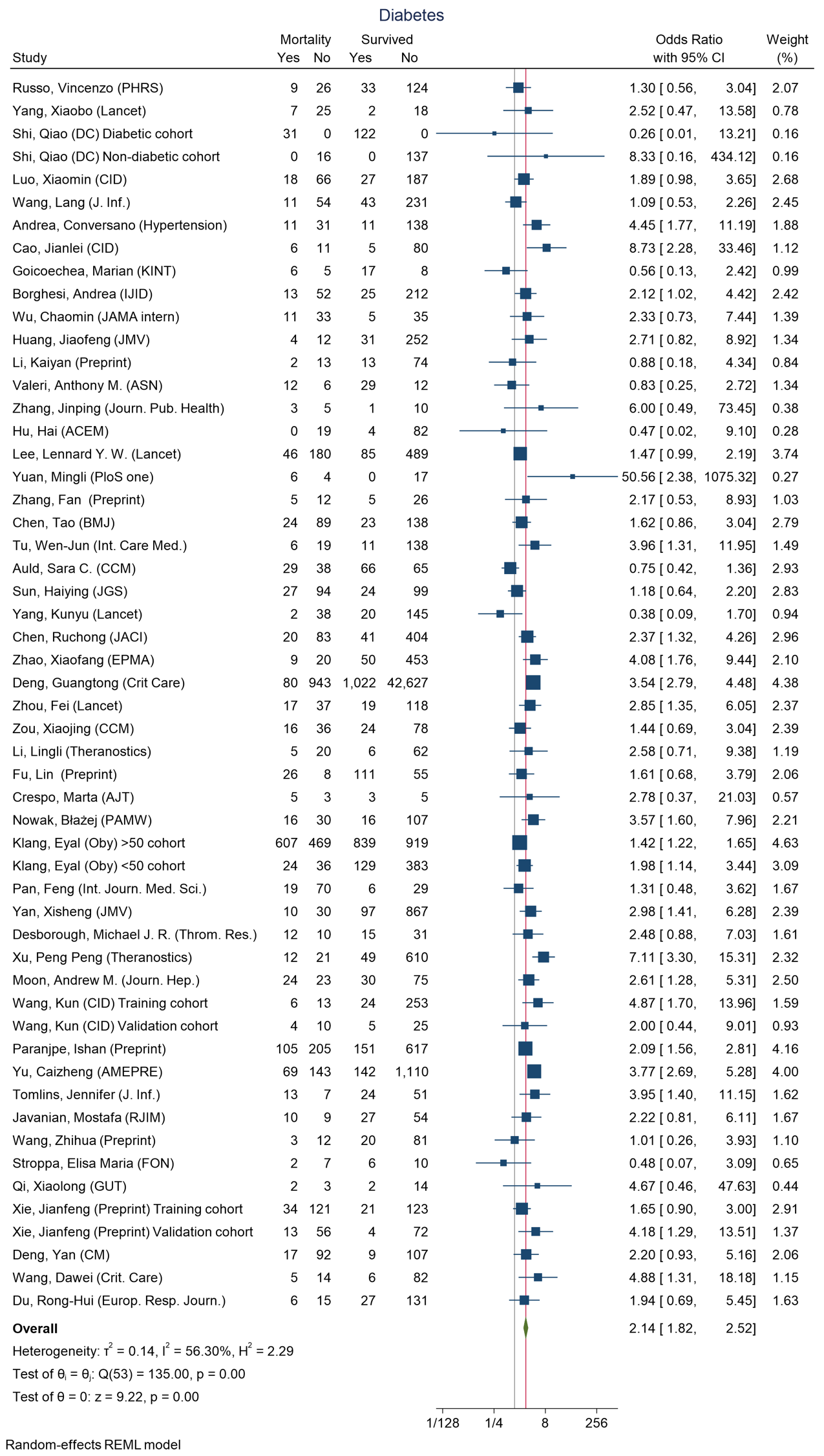

Figure 22 - Forest plot for Odds Ratio and 95% Confidence Interval (CI) of **Diabetes** for those who died vs. survived

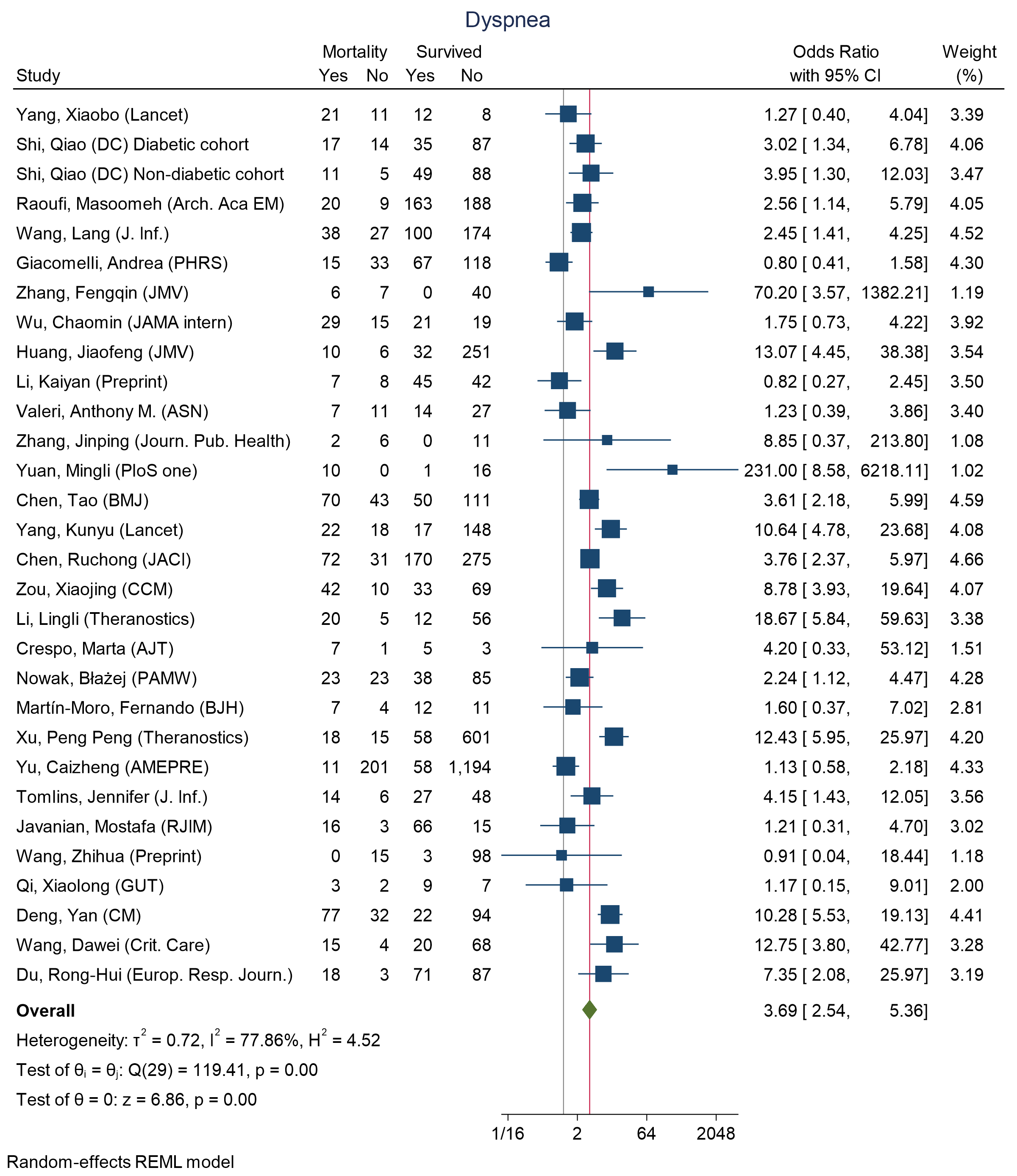

Figure 23 - Forest plot for Odds Ratio and 95% Confidence Interval (CI) of **Dyspnea** for those who died vs. survived

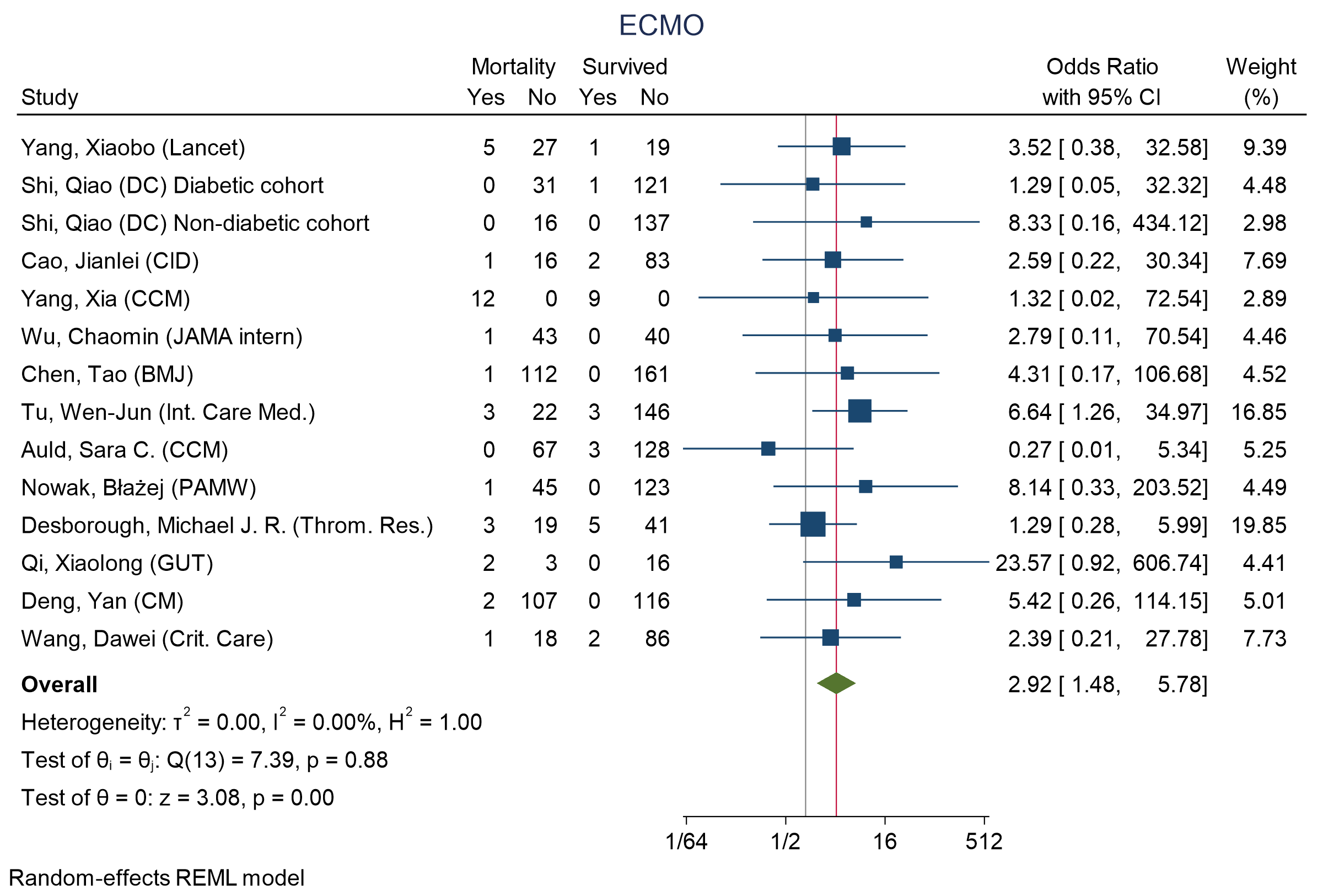

Figure 24 - Forest plot for Odds Ratio and 95% Confidence Interval (CI) of **extra corporal membrane oxygenation (ECMO)** for those who died vs. survived

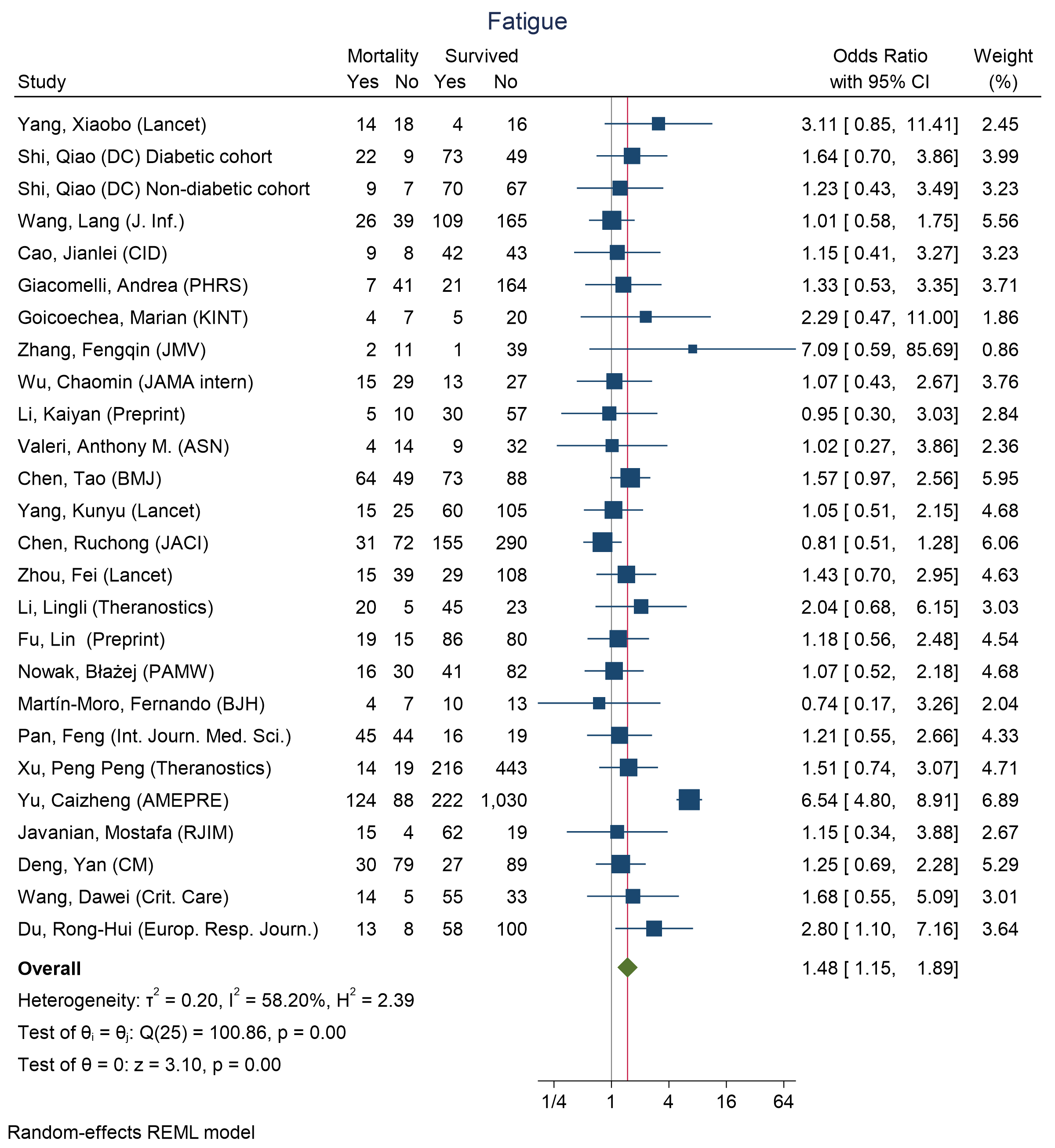

Figure 25 - Forest plot for Odds Ratio and 95% Confidence Interval (CI) of **Fatigue** for those who died vs. survived

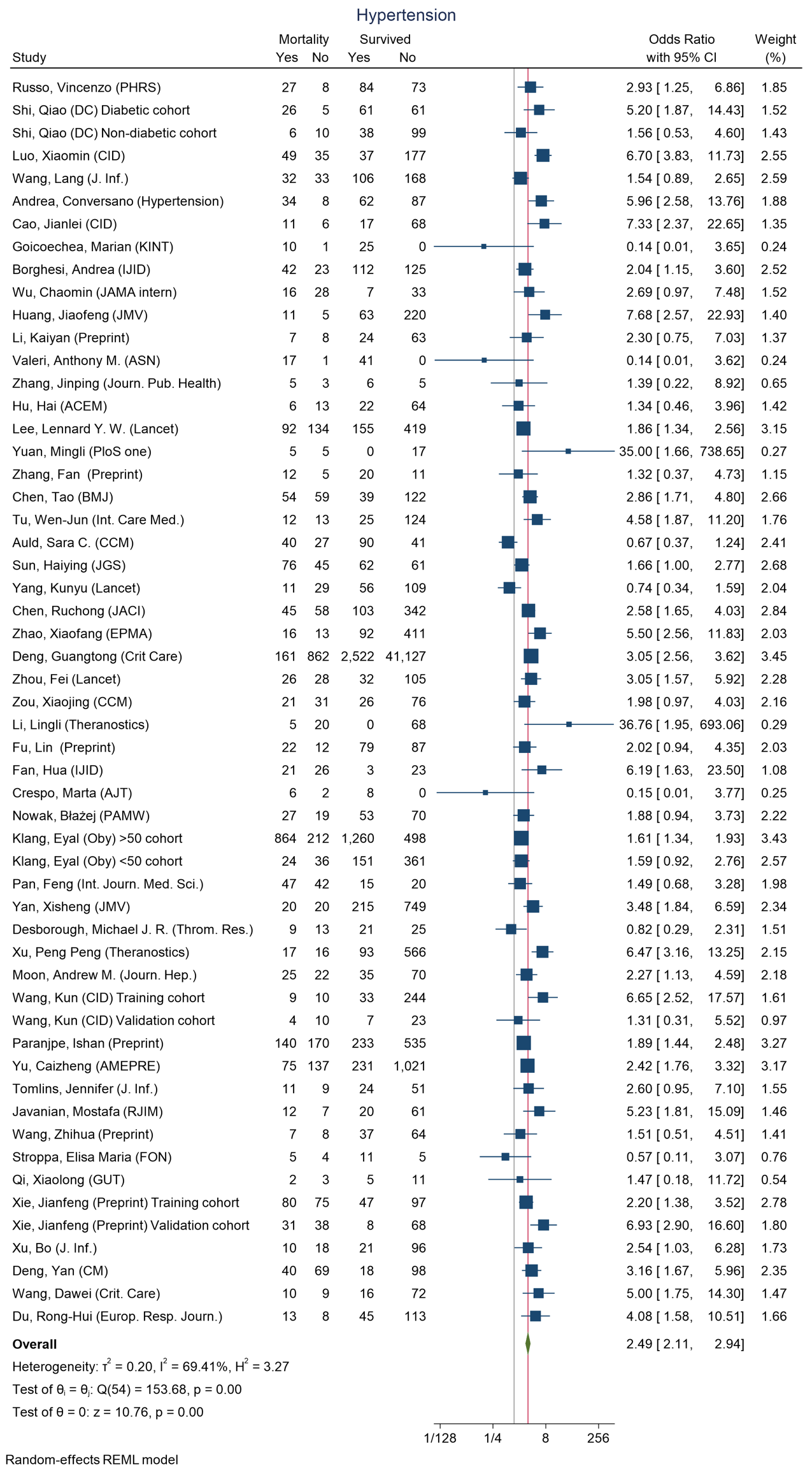

Figure 26 - Forest plot for Odds Ratio and 95% Confidence Interval (CI) of **Hypertension** for those who died vs. survived

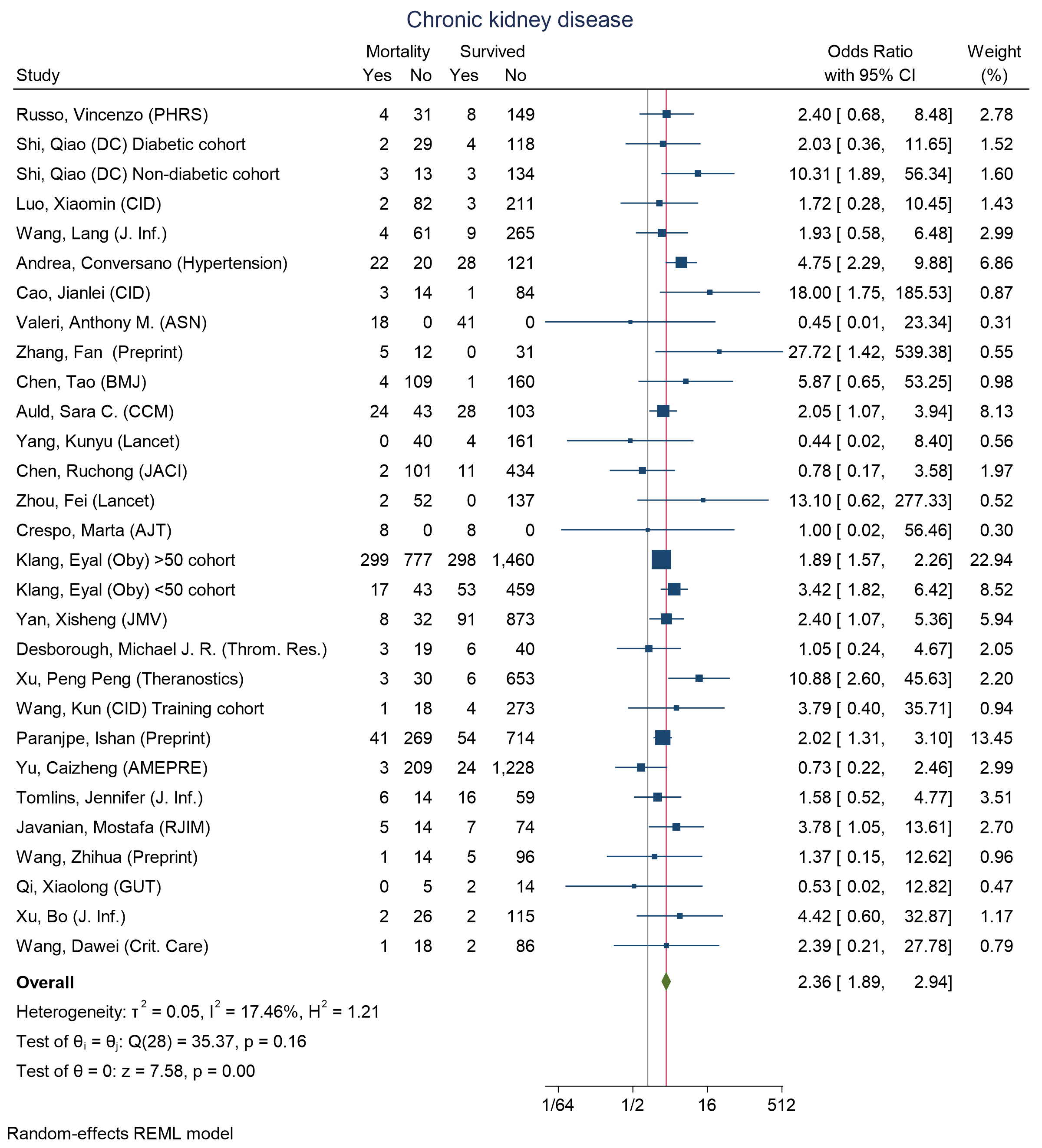

Figure 27 - Forest plot for Odds Ratio and 95% Confidence Interval (CI) of **Chronic kidney disease** for those who died vs. survived

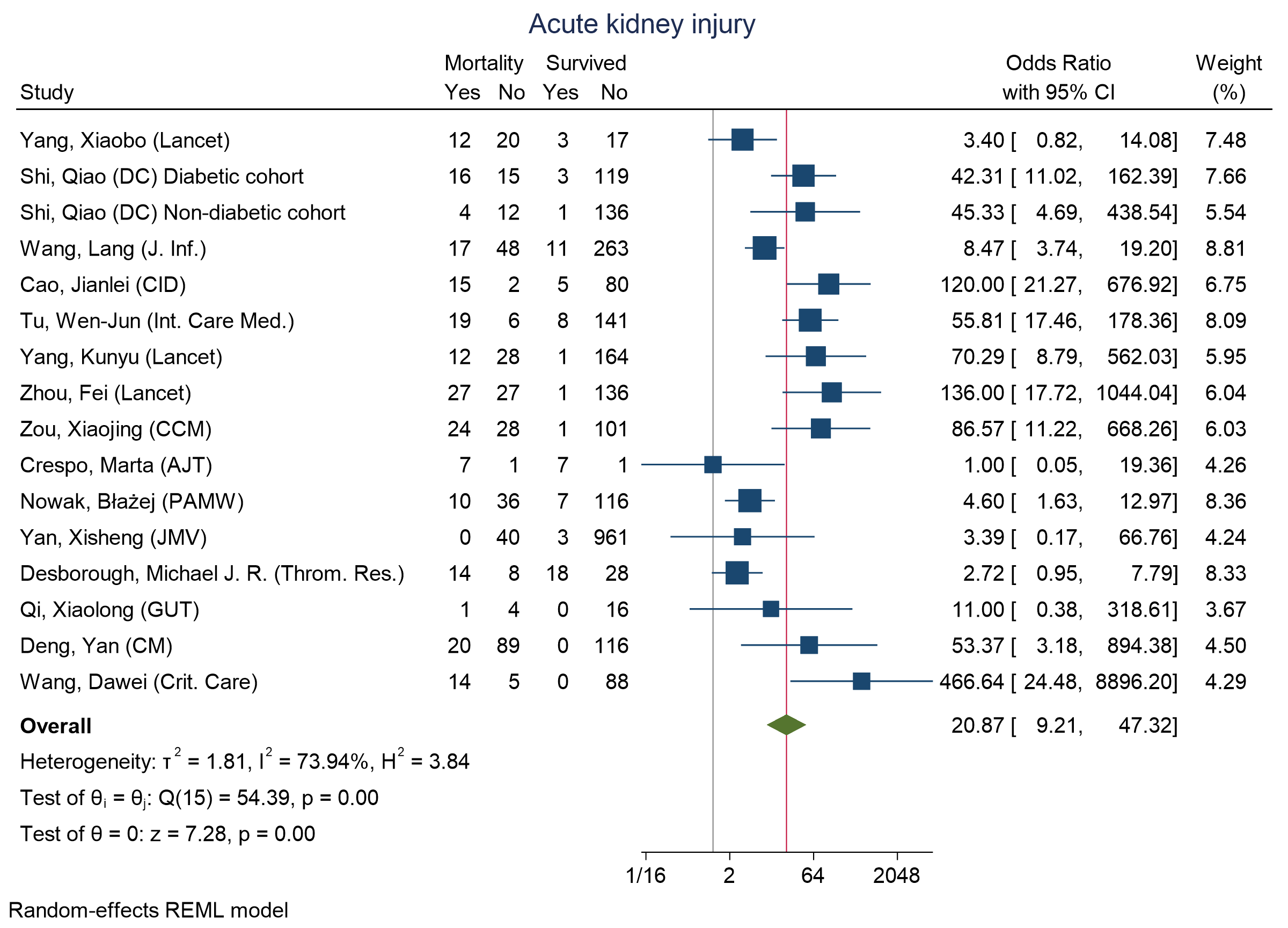

Figure 28 - Forest plot for Odds Ratio and 95% Confidence Interval (CI) of **Acute kidney injury** for those who died vs. survived

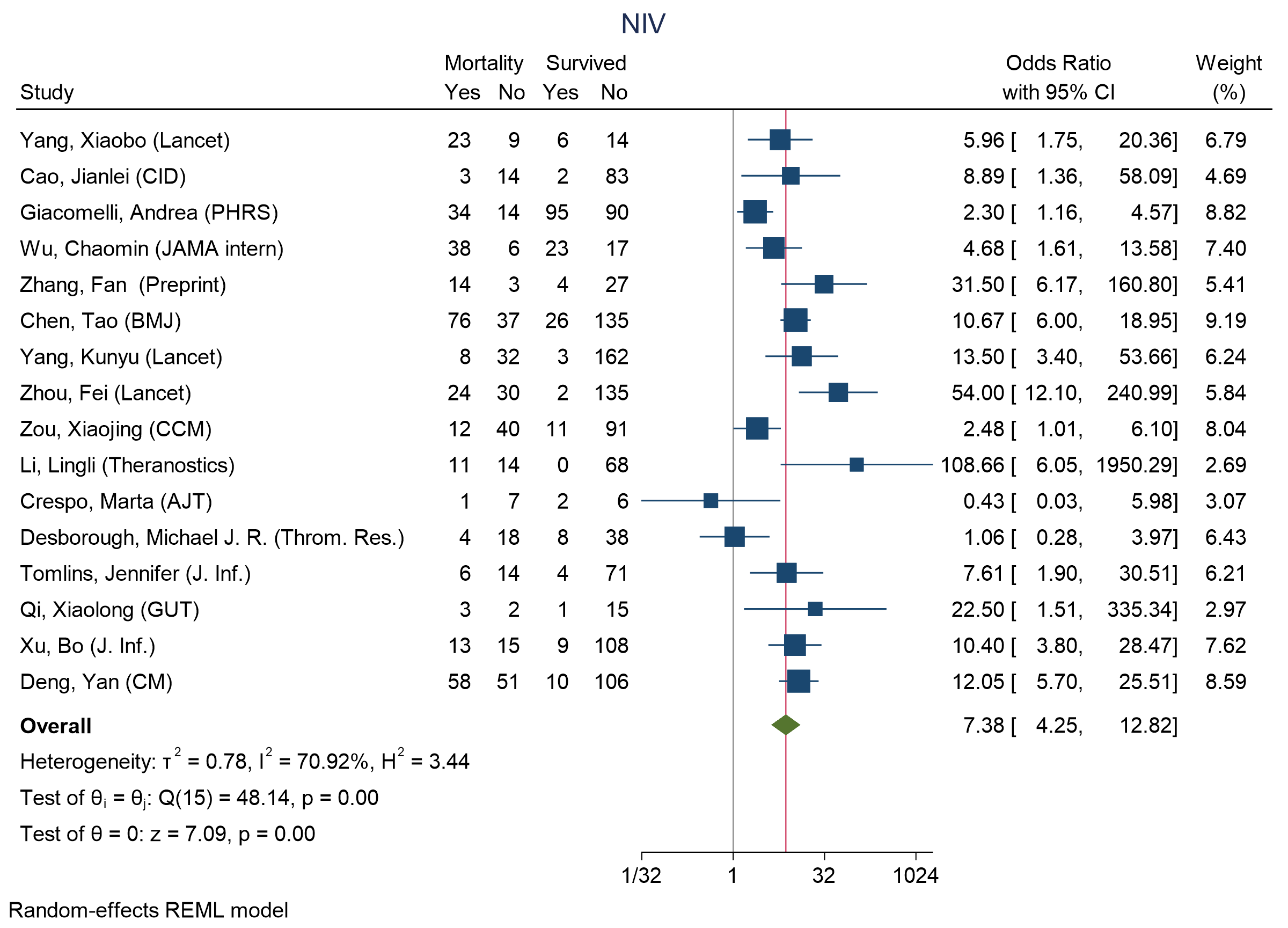

Figure 29 - Forest plot for Odds Ratio and 95% Confidence Interval (CI) of **non-invasive ventilation (NIV)** for those who died vs. survived

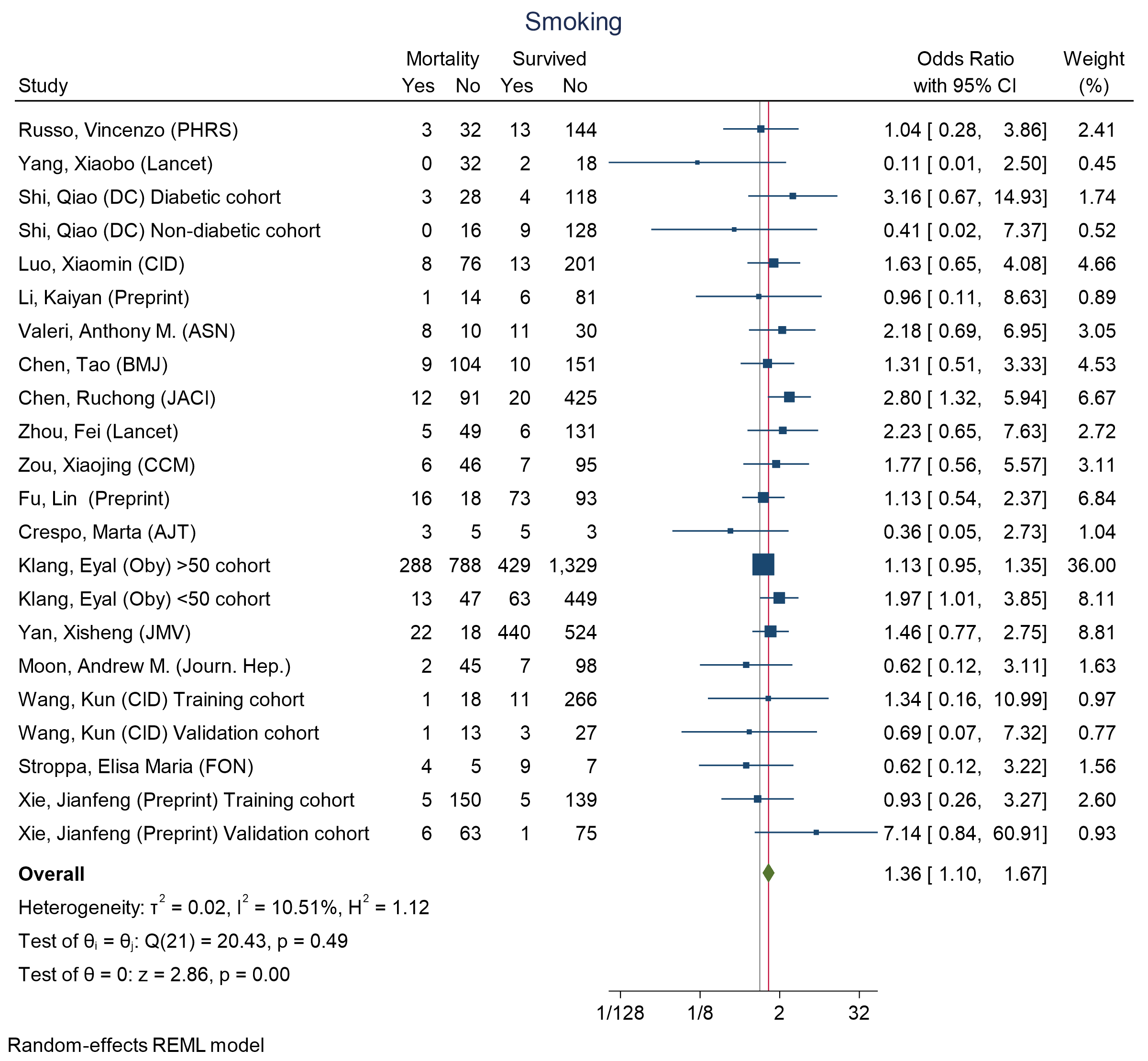

Figure 30 - Forest plot for Odds Ratio and 95% Confidence Interval (CI) of **Smoking** for those who died vs. survived

### S6 Forest plots for difference of medians in ICU admission vs. non- ICU admission across different indicators

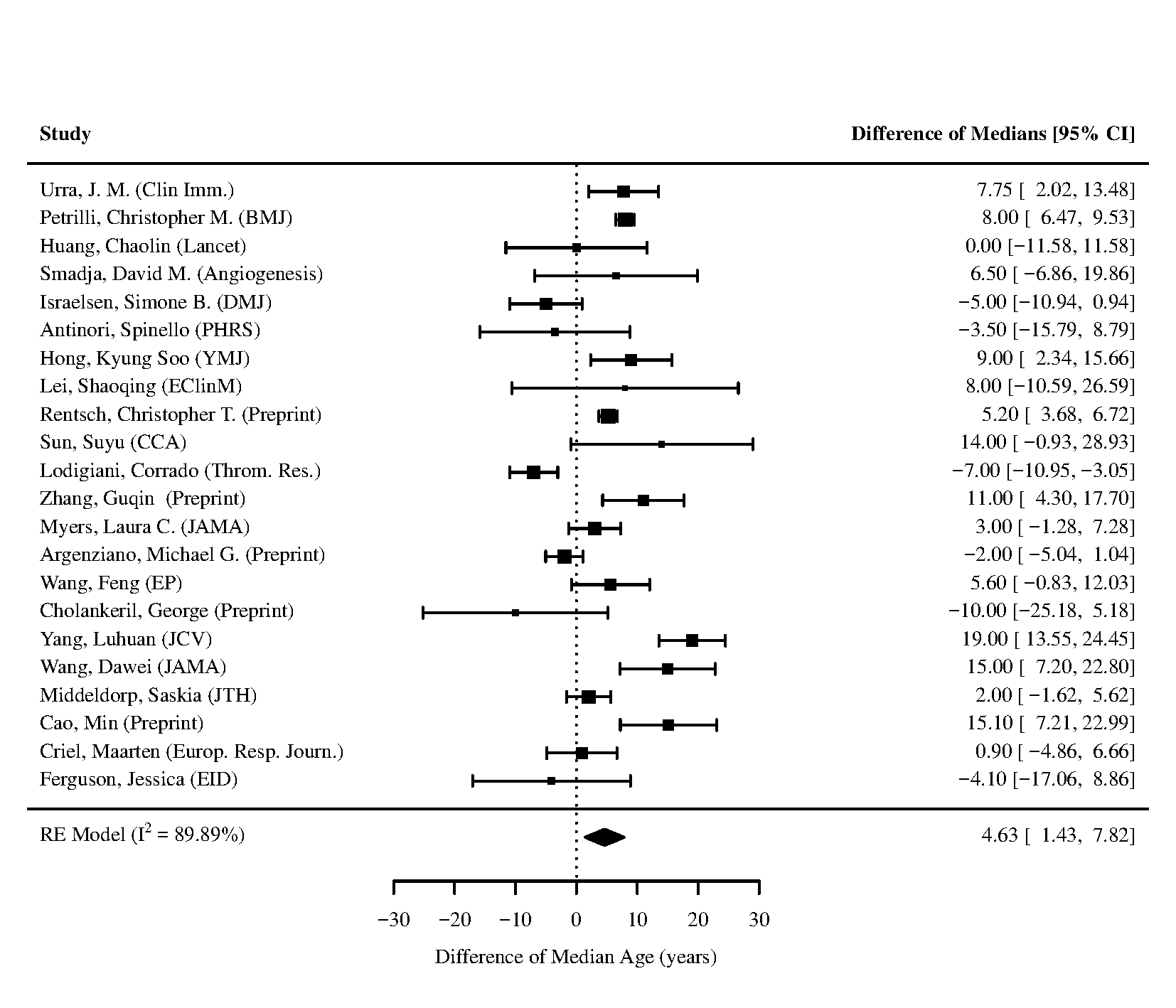

Figure 31 - Forest plot for difference of medians of **Age** for those who require ICU admission vs. non-ICU admission

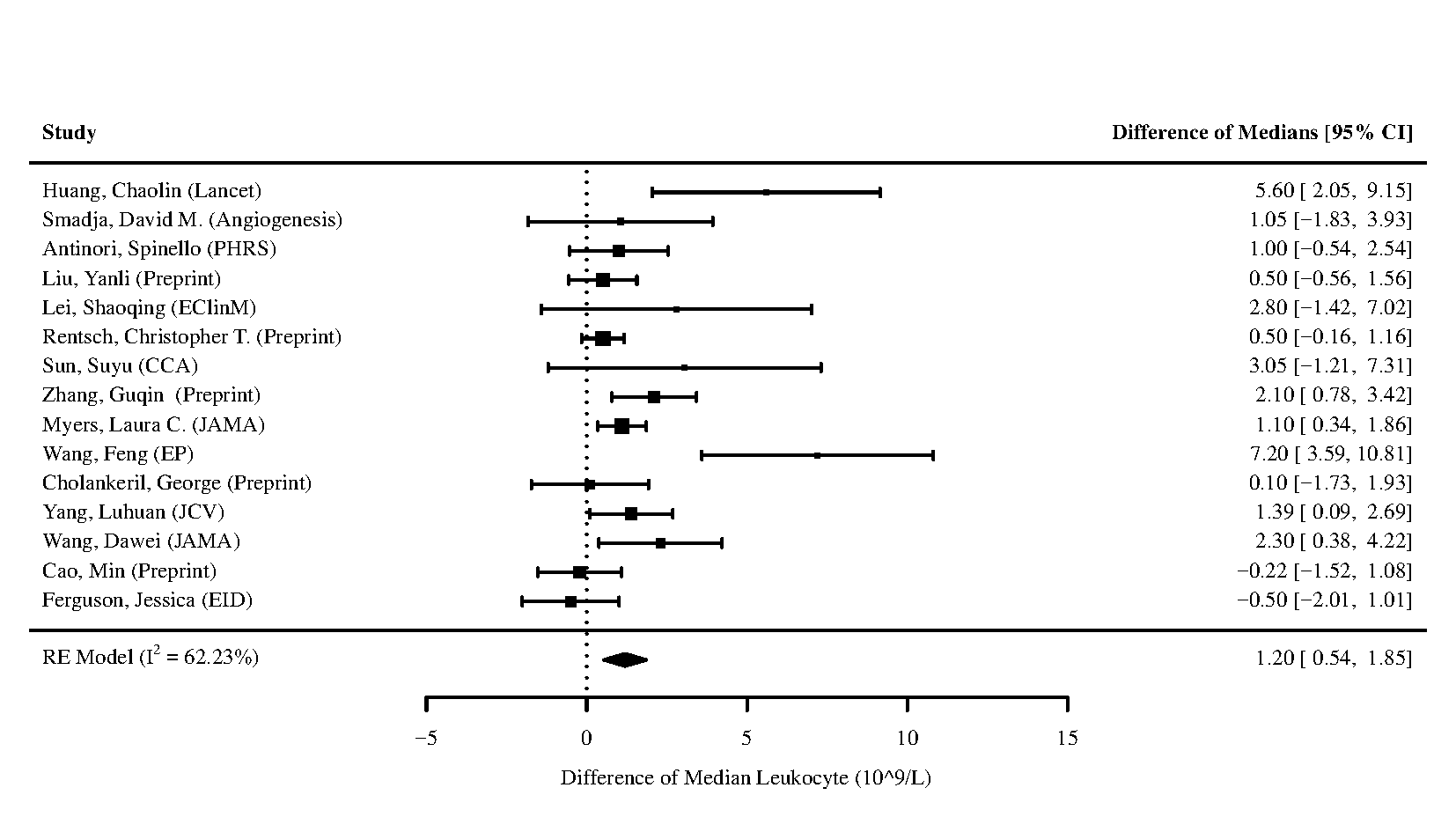

Figure 32 - Forest plot for difference of medians of **Leukocytes** for those who require ICU admission vs. non-ICU admission

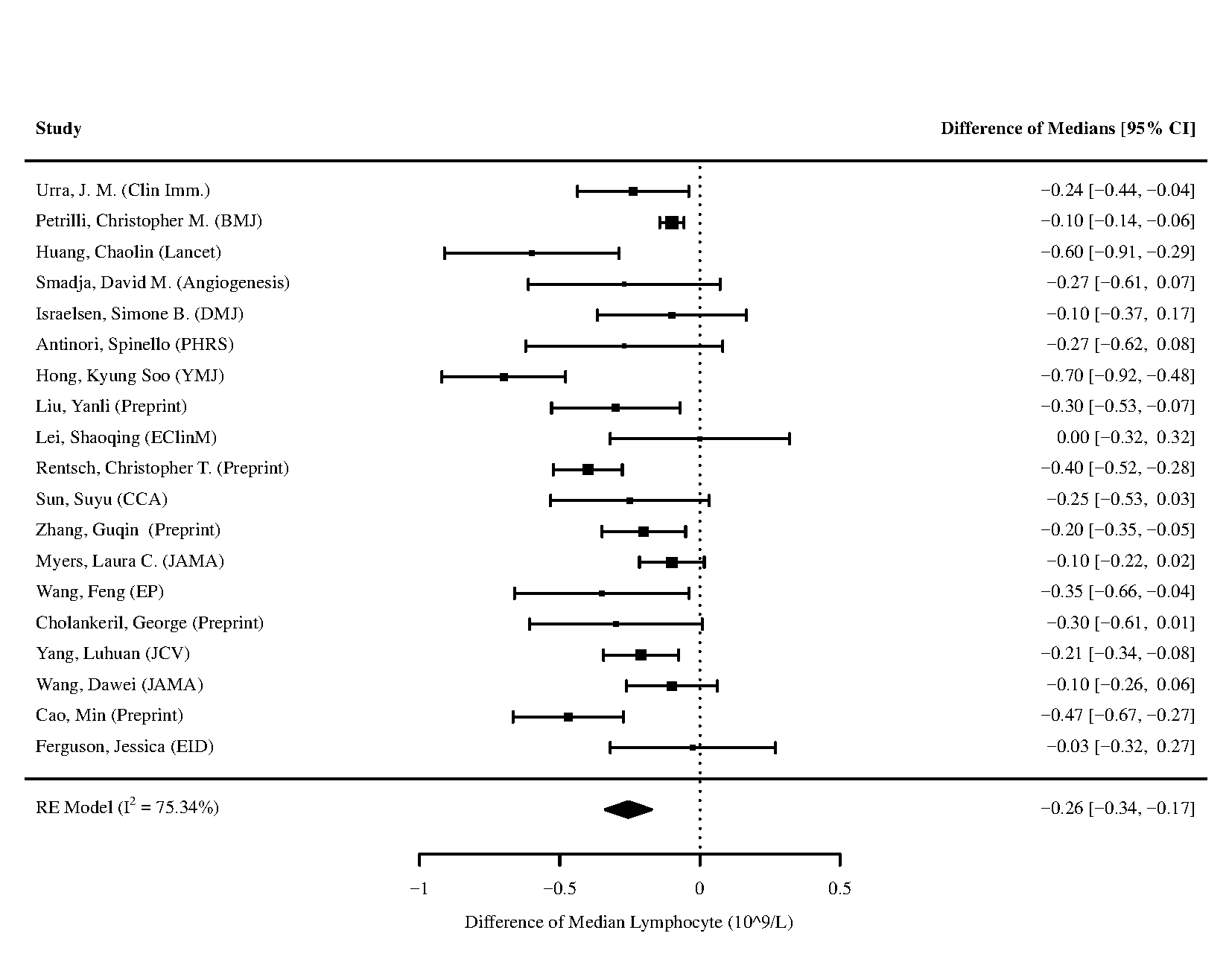

Figure 33 - Forest plot for difference of medians of **Lymphocyte** for those who require ICU admission vs. non-ICU admission

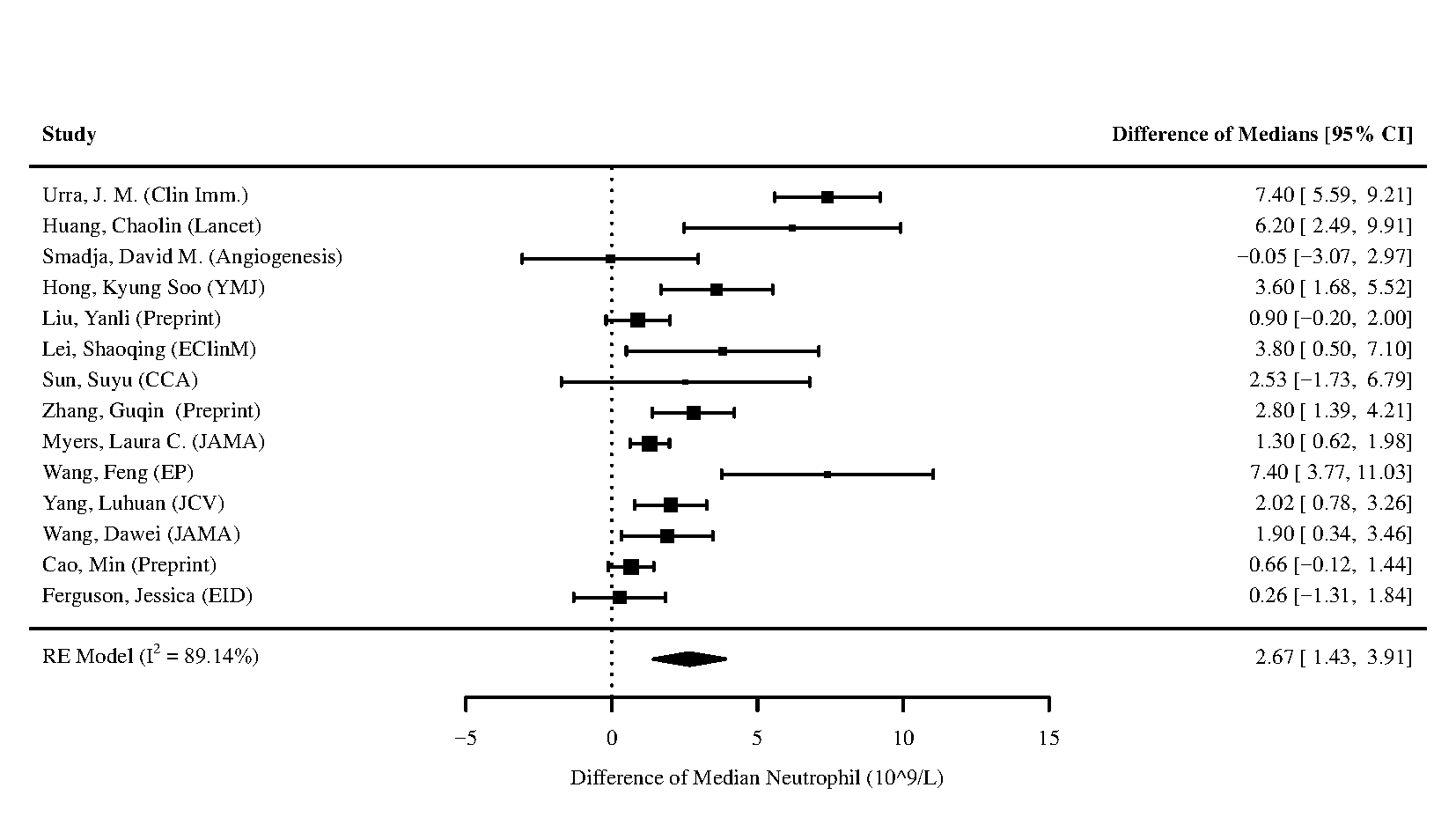

Figure 34 - Forest plot for difference of medians of **Neutrophil** for those who require ICU admission vs. non-ICU admission

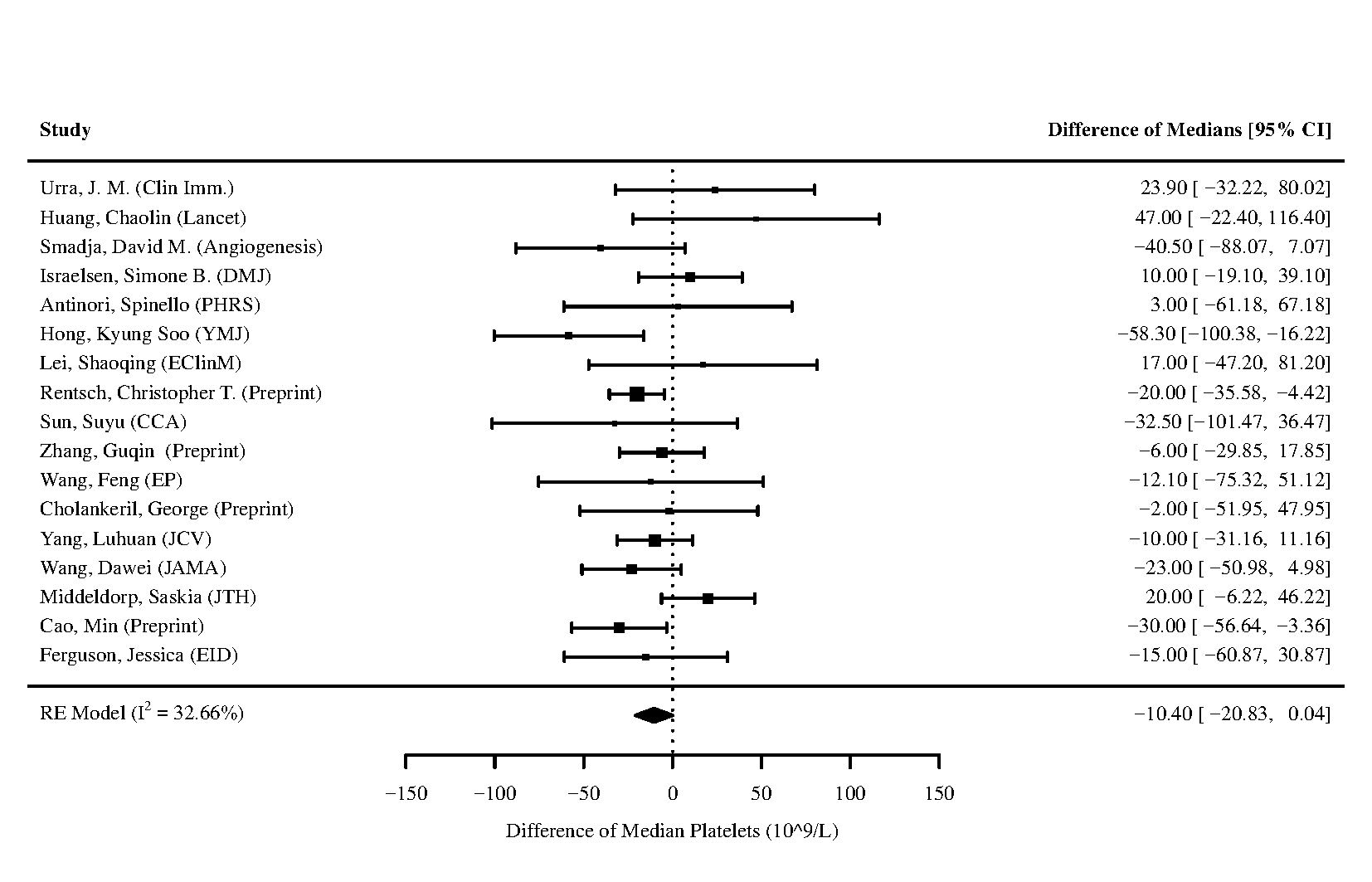

Figure 35 - Forest plot for difference of medians of **Platelets** for those who require ICU admission vs. non-ICU admission

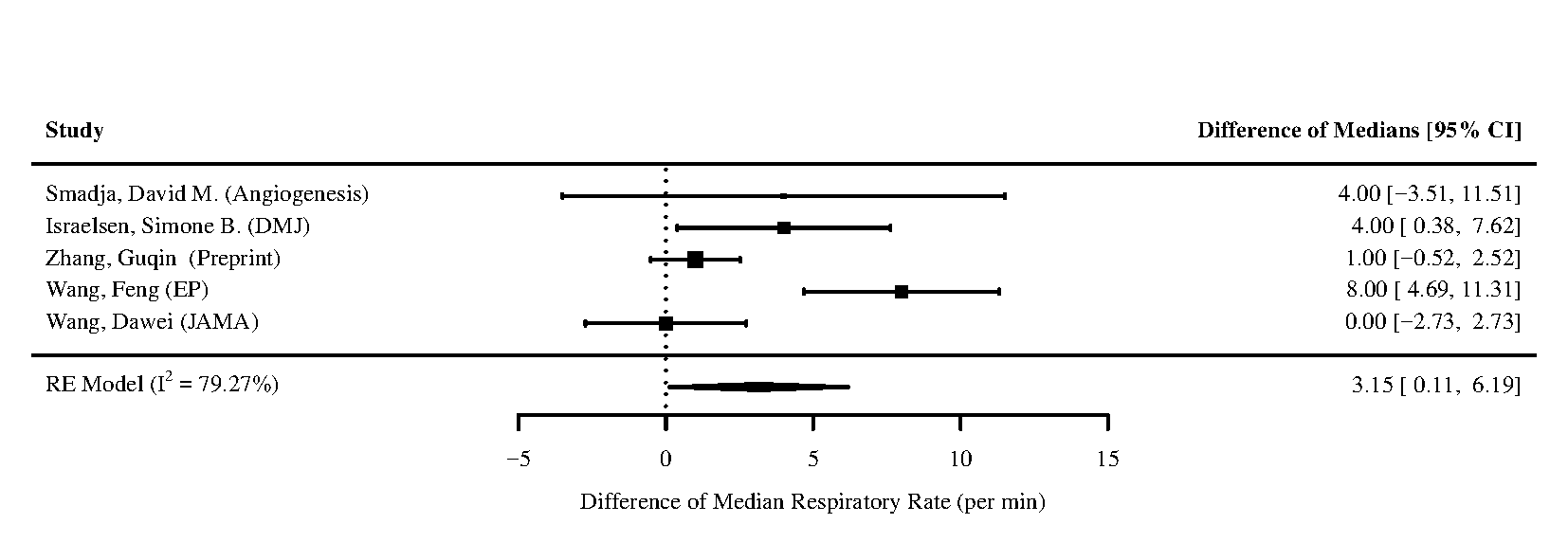

Figure 36 - Forest plot for difference of medians of **Respiratory Rate** for those who require ICU admission vs. non-ICU admission

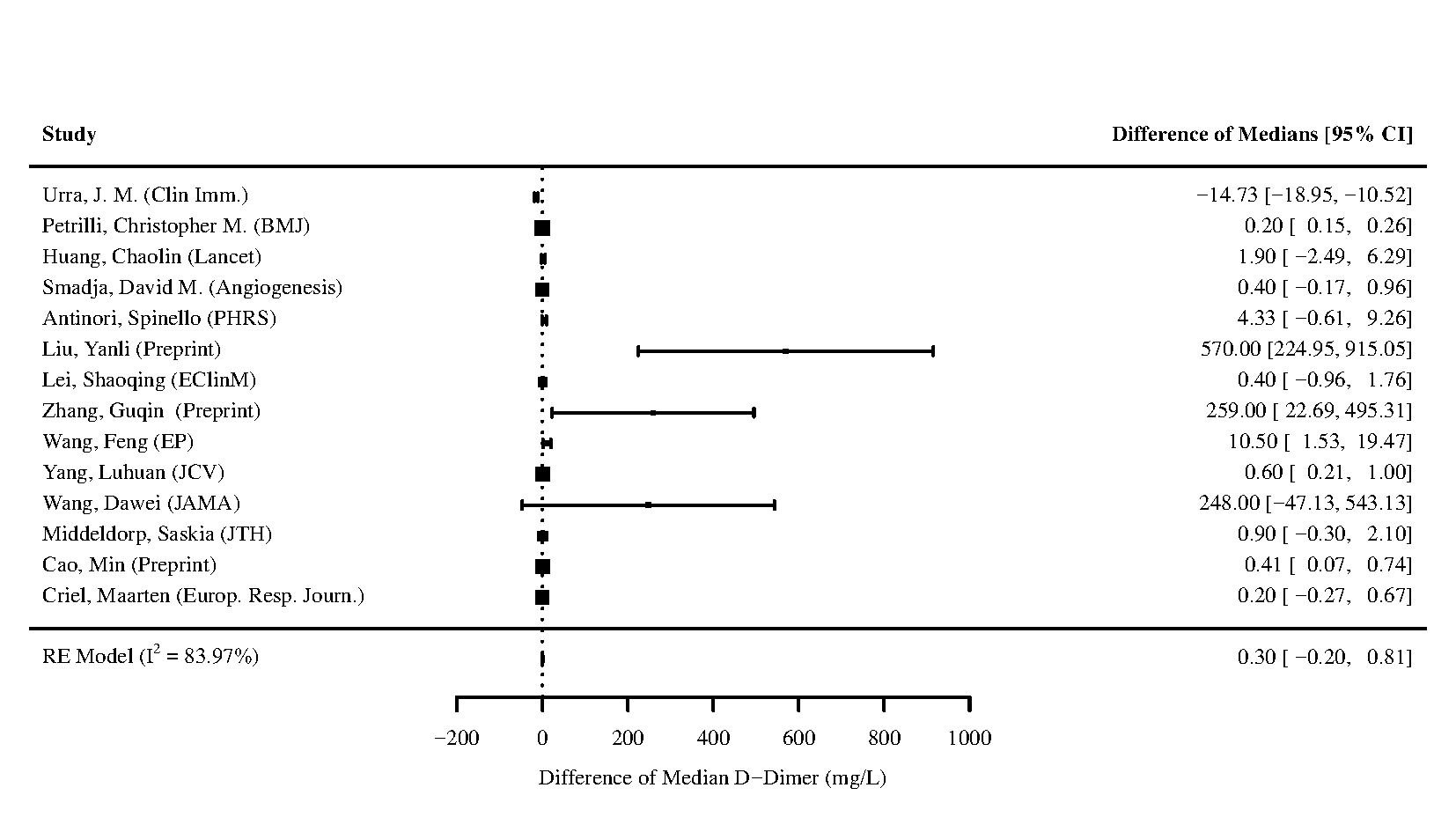

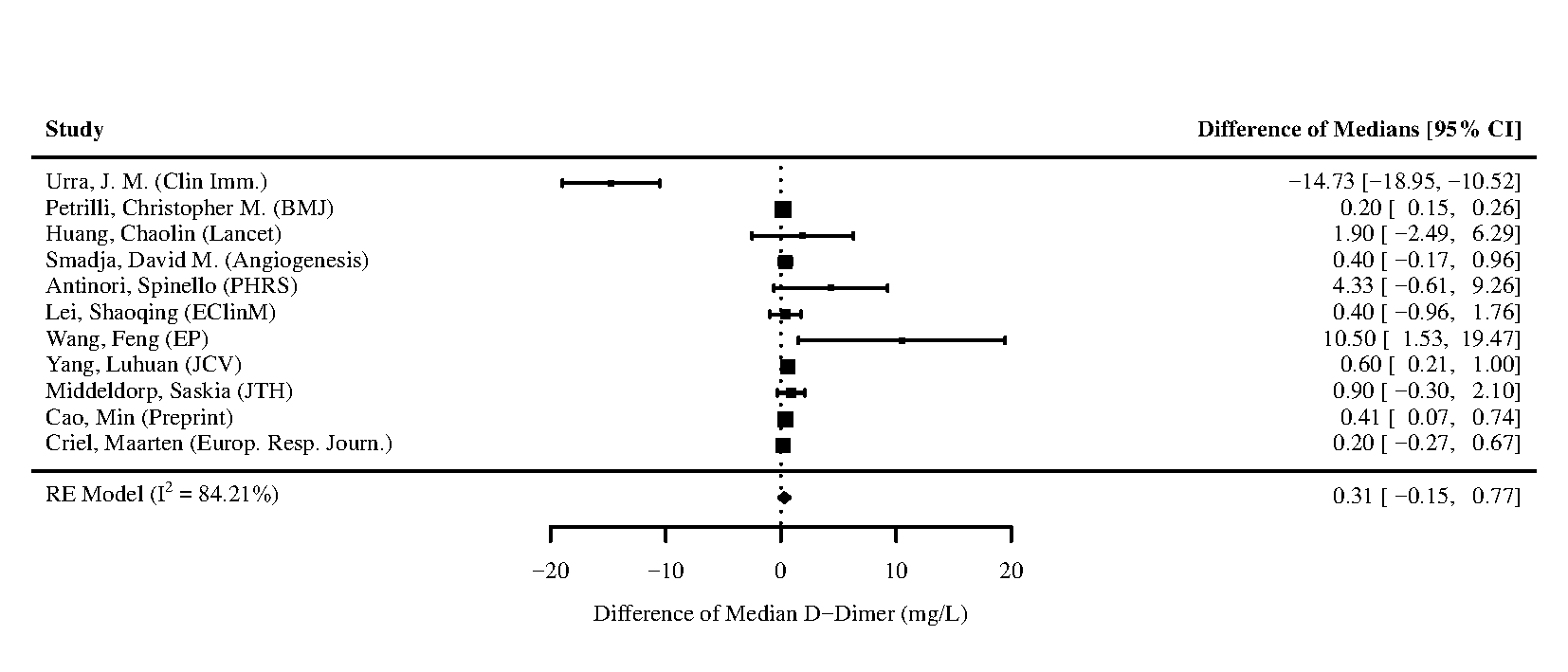

Figure 37 - Forest plot for difference of medians of **D-Dimer** for those who require ICU admission vs. non-ICU admission. Note that some studies used different laboratory assays leading to different ranges. The left panel contains the forest plot of the analysis including the studies by Liu, Yanli, Zhang, Guqin, and Wang, Dawei, and the right panel contains the forest plot of the analysis excluding the studies by Liu, Yanli, Zhang, Guqin, and Wang, Dawei.

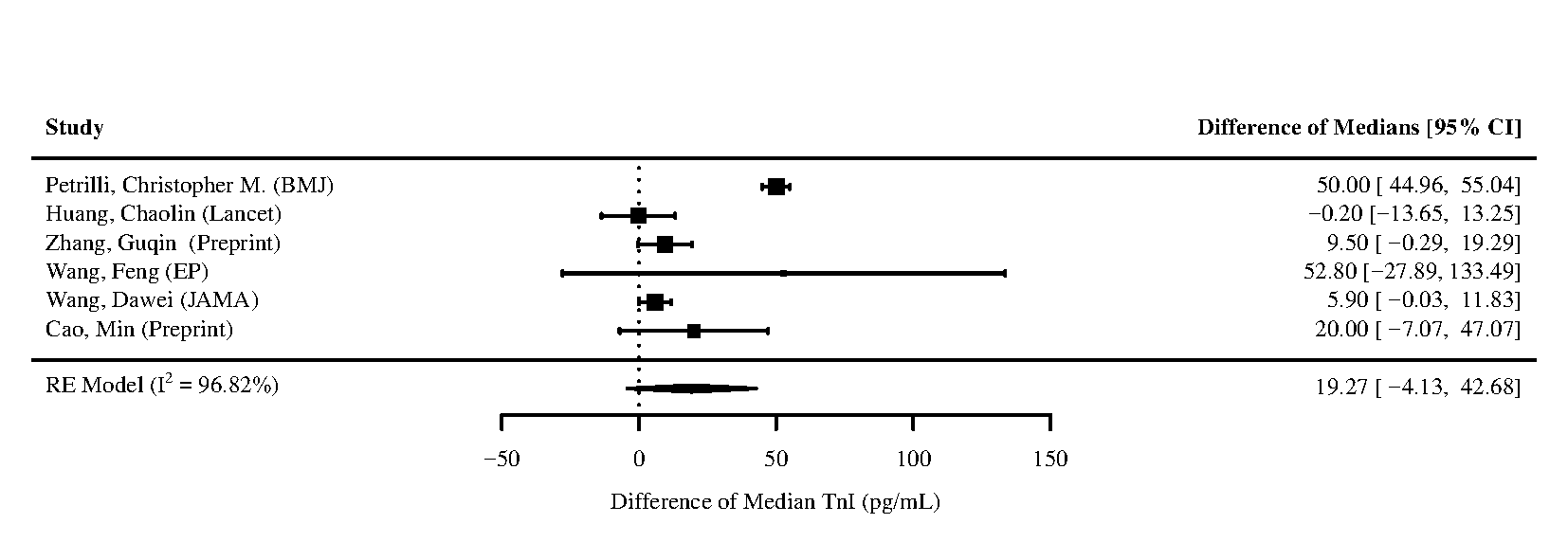

Figure 38 - Forest plot for difference of medians of **Troponin I (TnI)** for those who require ICU admission vs. non-ICU admission

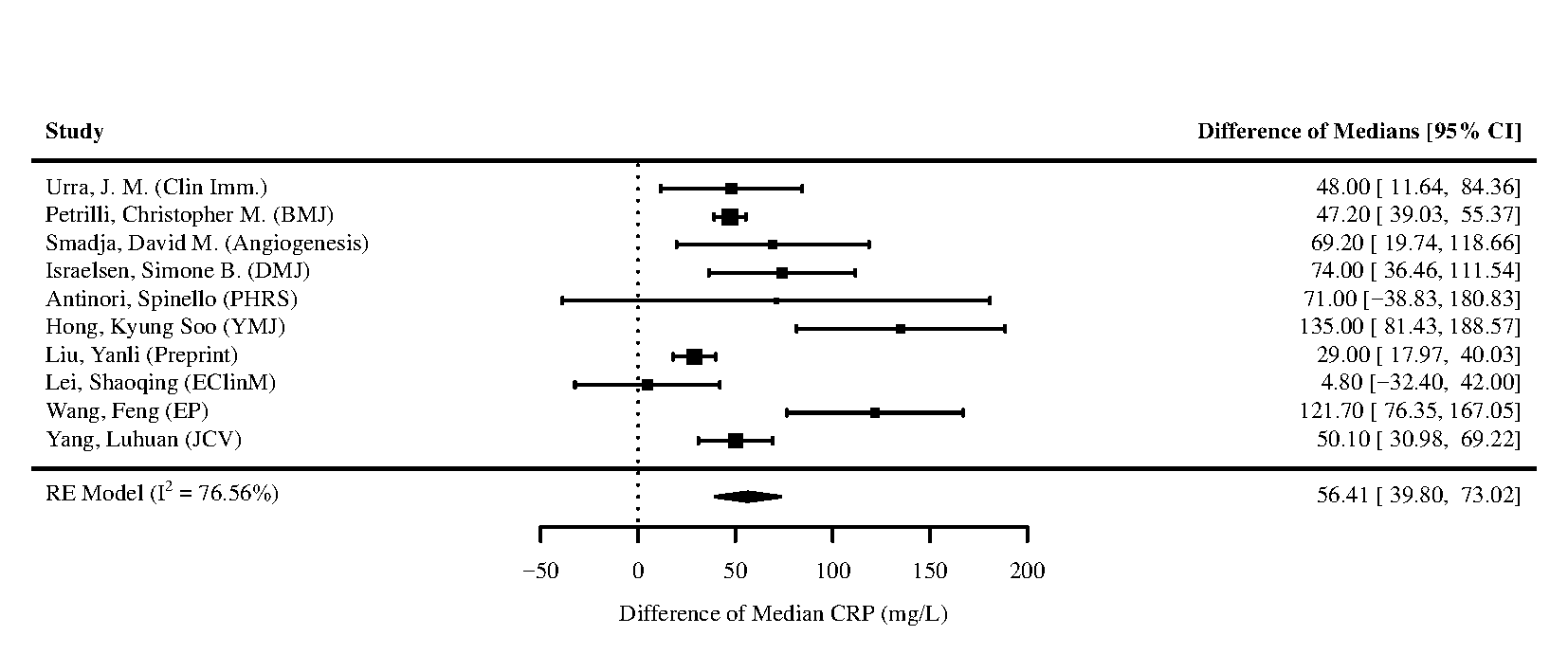

Figure 39 - Forest plot for difference of medians of **C-reactive protein (CRP)** for those who require ICU admission vs. non-ICU admission

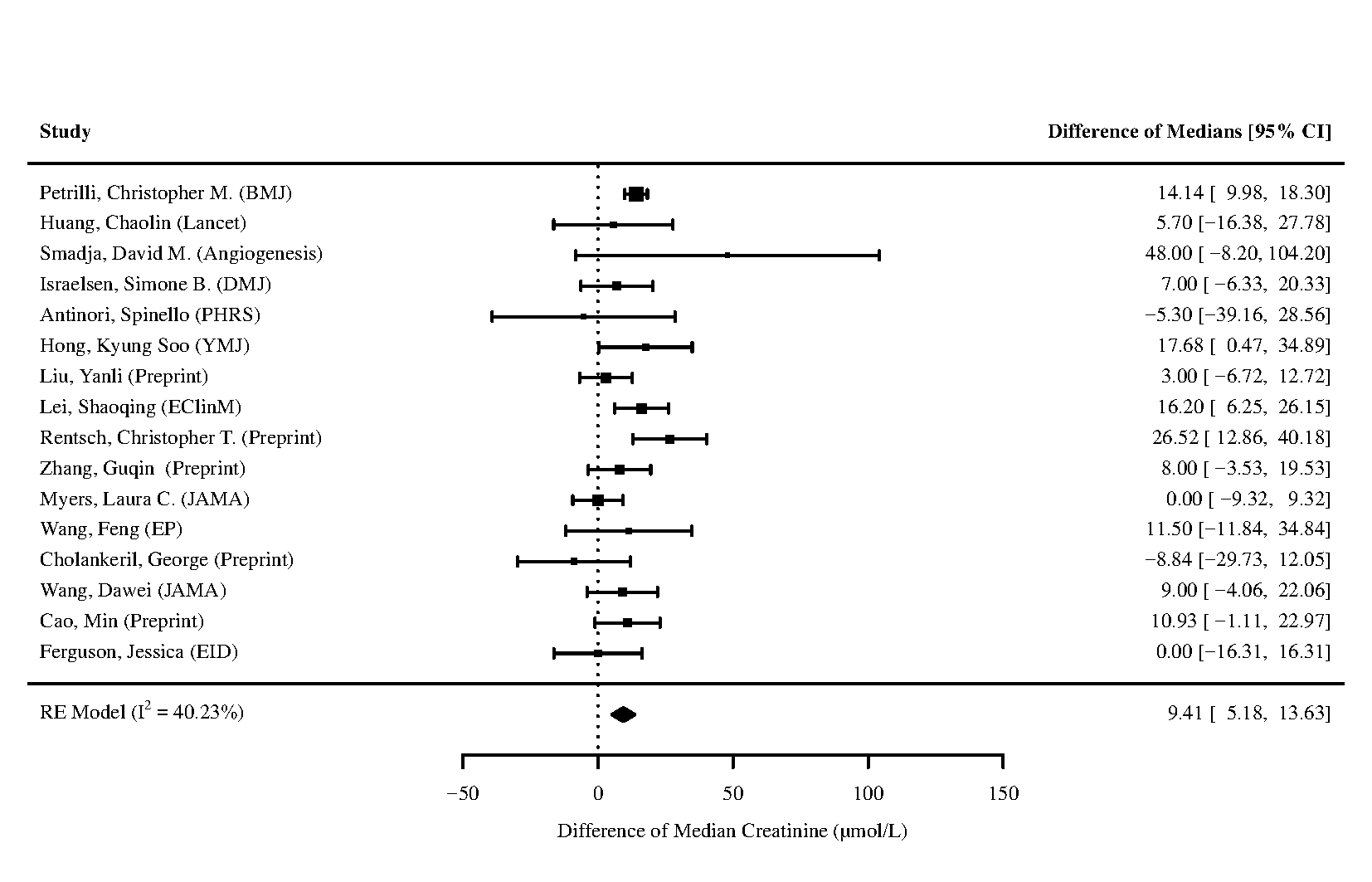

Figure 40 - Forest plot for difference of medians of **Creatinine** for those who require ICU admission vs. non-ICU admission

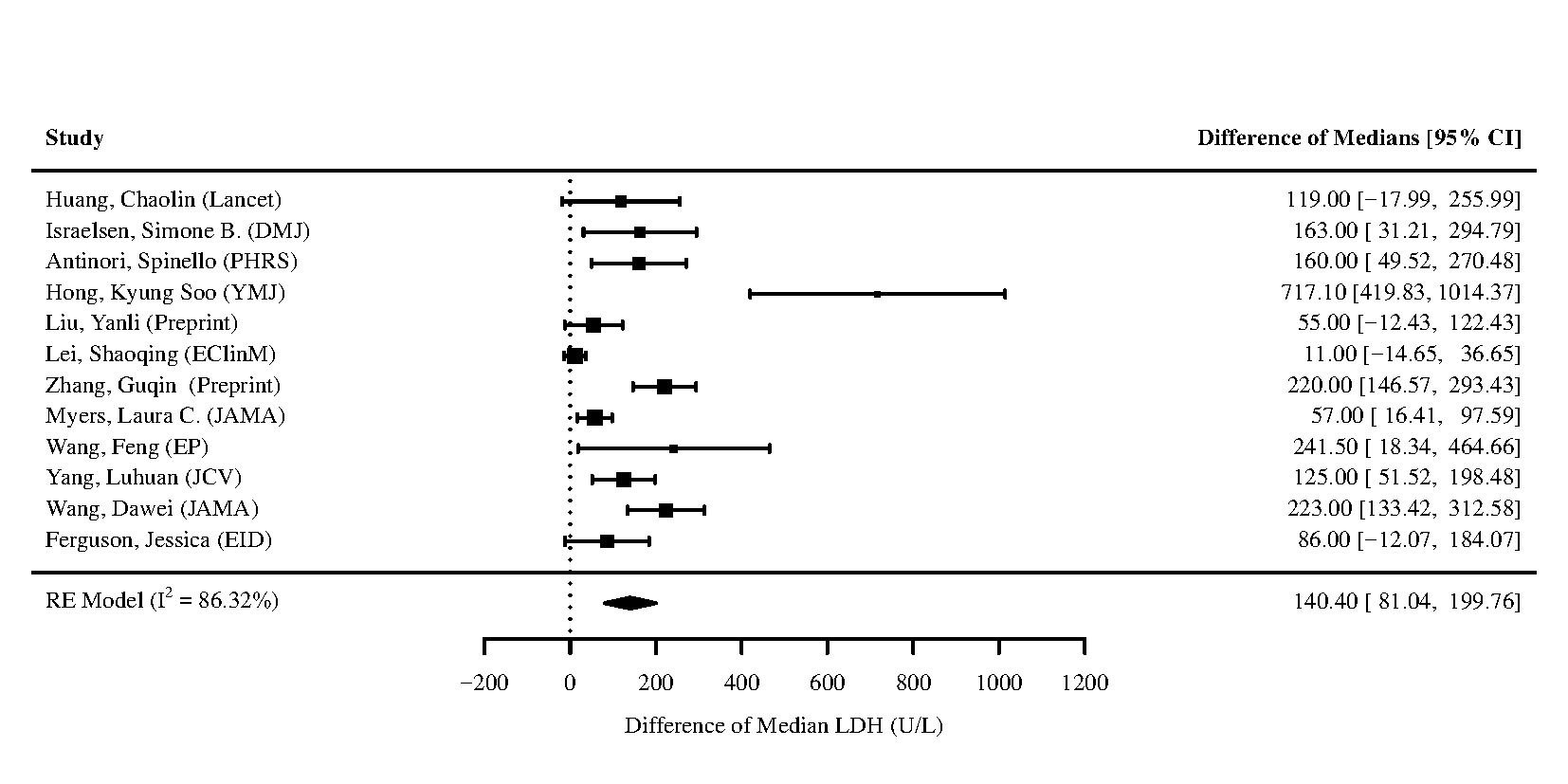

Figure 41 - Forest plot for difference of medians of **Lactate Dehydrogenase (LDH)** for those who require ICU admission vs. non-ICU admission

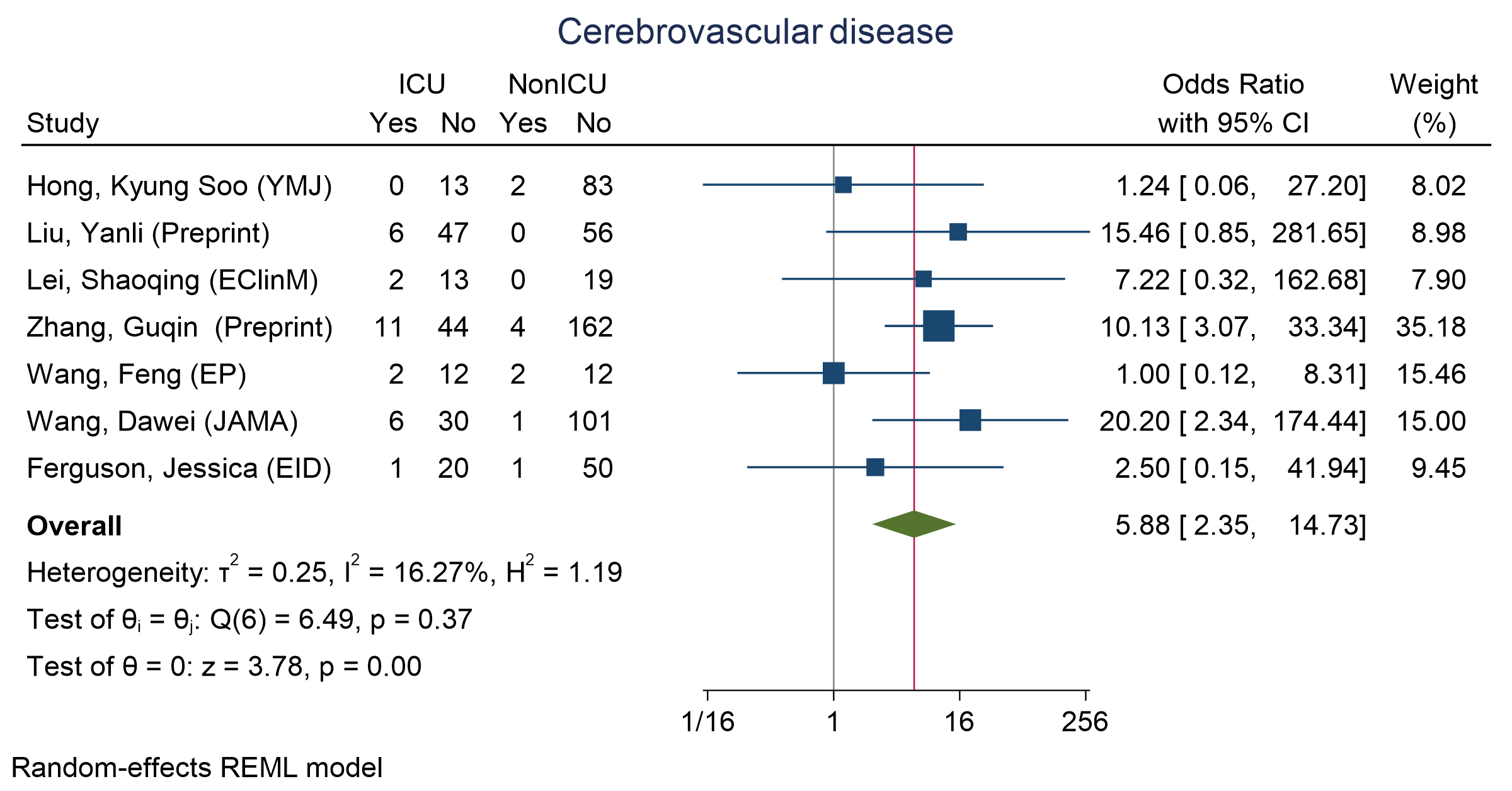

Figure 42 - Forest plot for Odds Ratio and 95% Confidence Interval (CI) of **Cerebrovascular disease** for those who require ICU admission vs. non-ICU admission

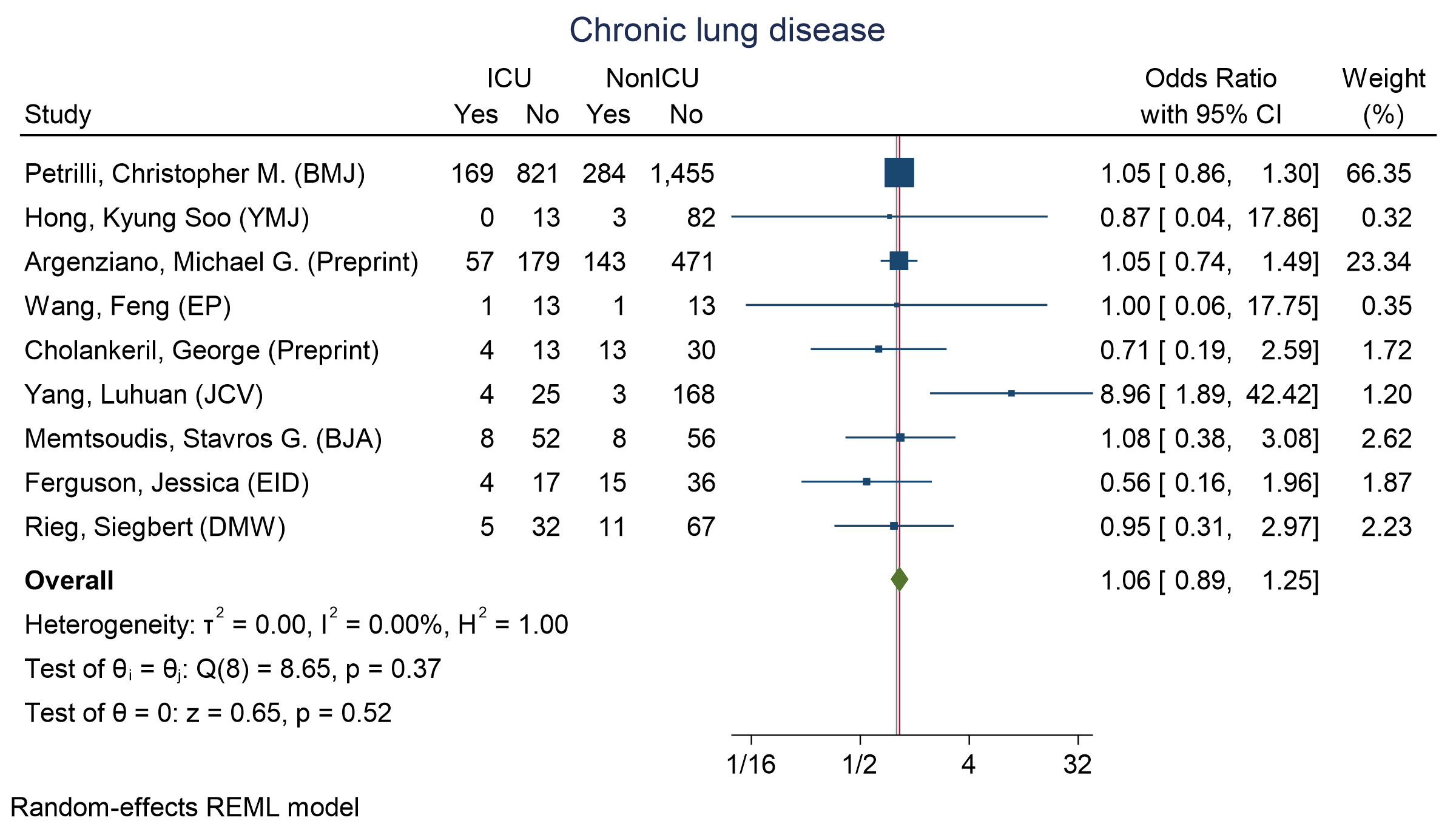

Figure 43 - Forest plot for Odds Ratio and 95% Confidence Interval (CI) of **Chronic Lung disease** for those who require ICU admission vs. non-ICU admission

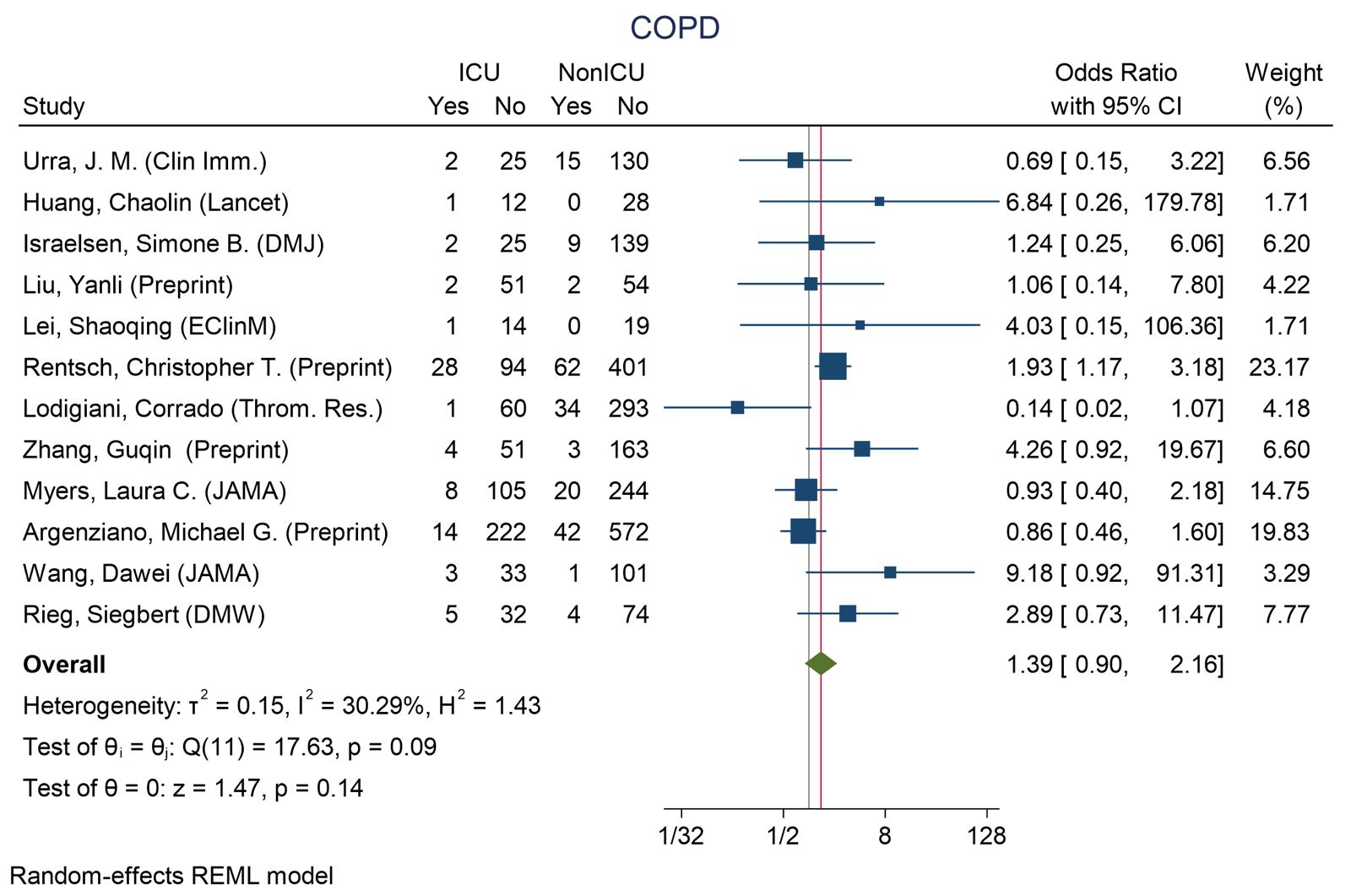

Figure 44 - Forest plot for Odds Ratio and 95% Confidence Interval (CI) of **COPD** for those who require ICU admission vs. non-ICU admission

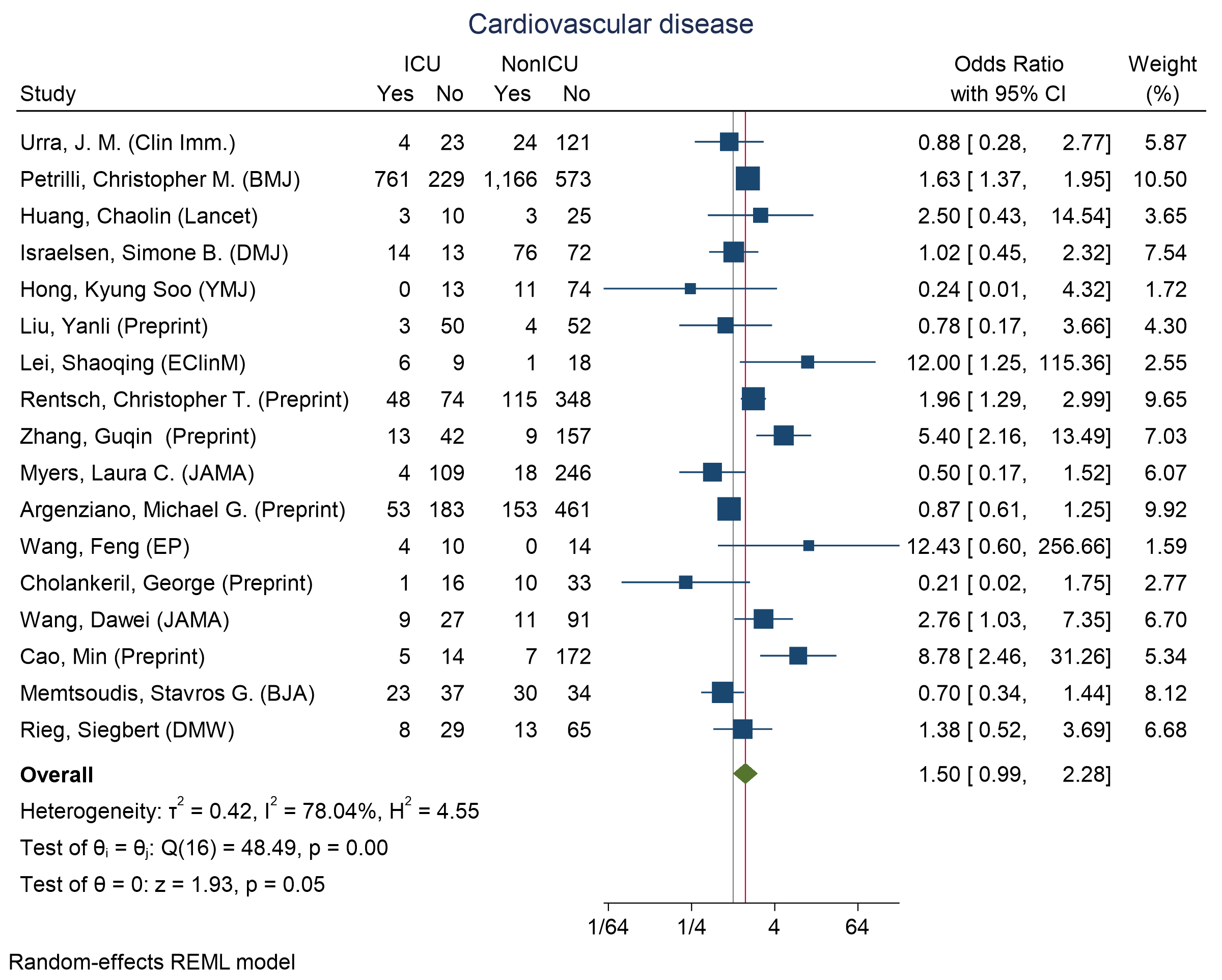

Figure 45 - Forest plot for Odds Ratio and 95% Confidence Interval (CI) of **Cardiovascular disease** for those who require ICU admission vs. non-ICU admission

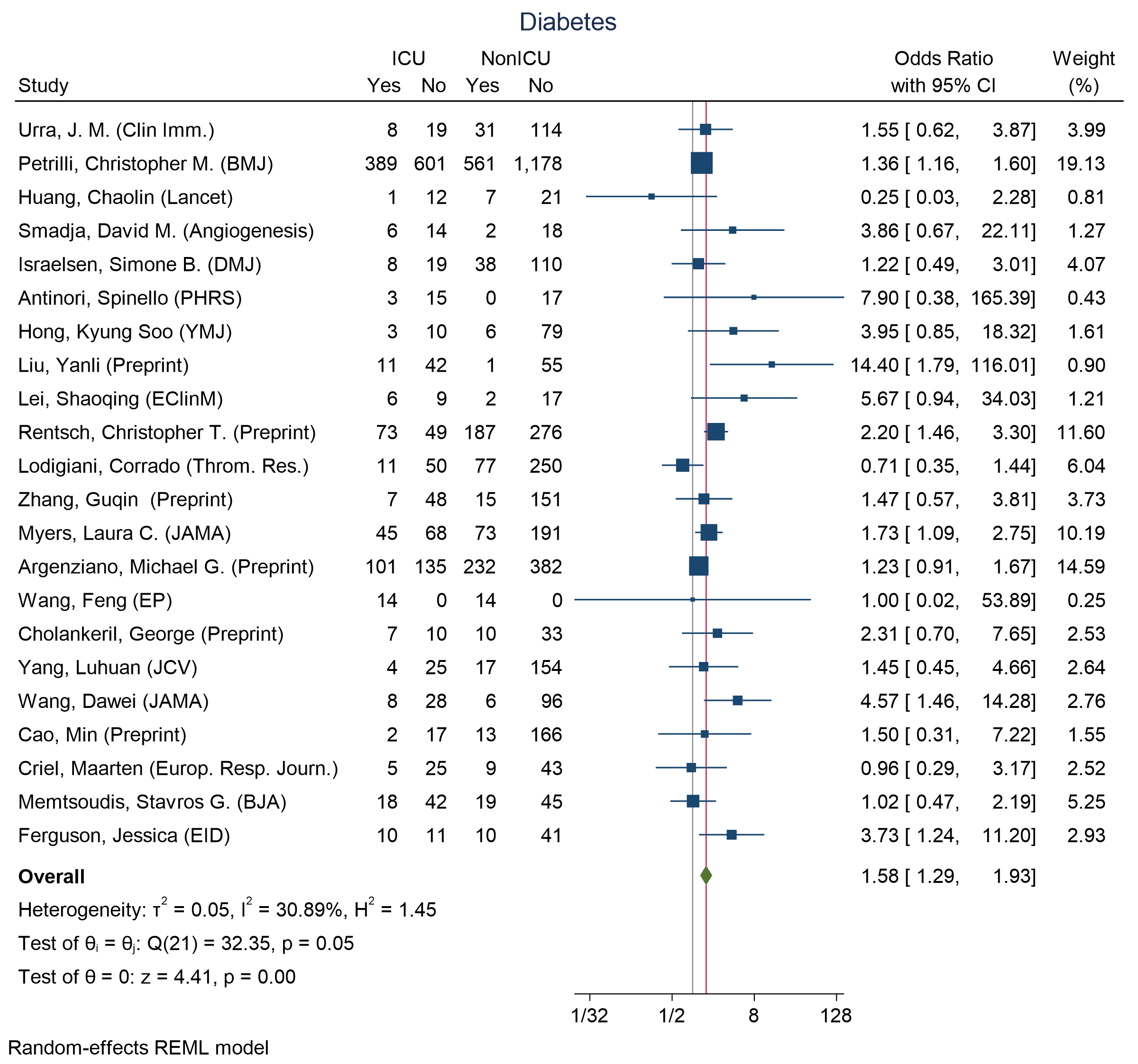

Figure 46 - Forest plot for Odds Ratio and 95% Confidence Interval (CI) of **Diabetes** for those who require ICU admission vs. non-ICU admission

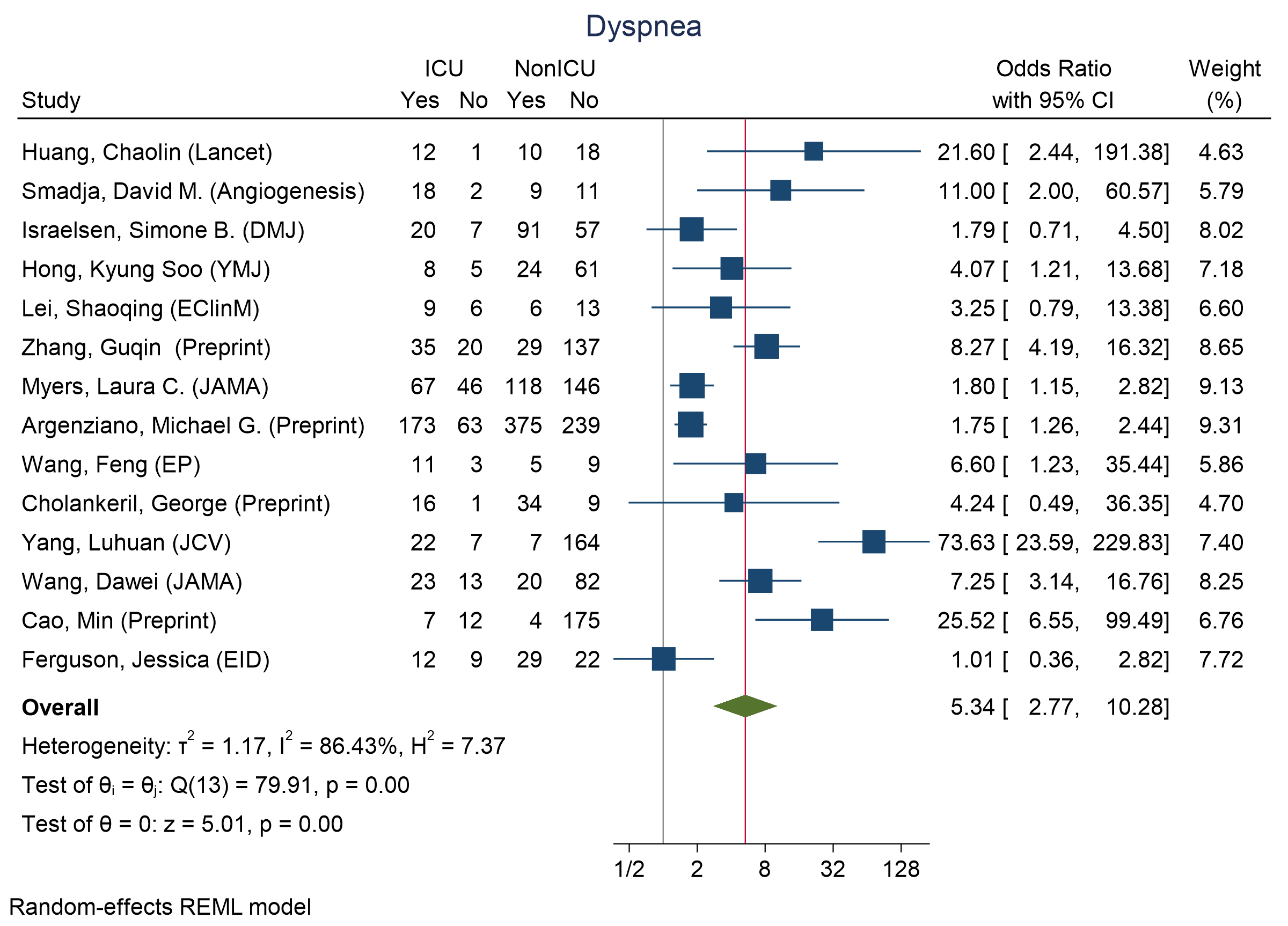

Figure 47 - Forest plot for Odds Ratio and 95% Confidence Interval (CI) of **Dyspnea** for those who require ICU admission vs. non-ICU admission

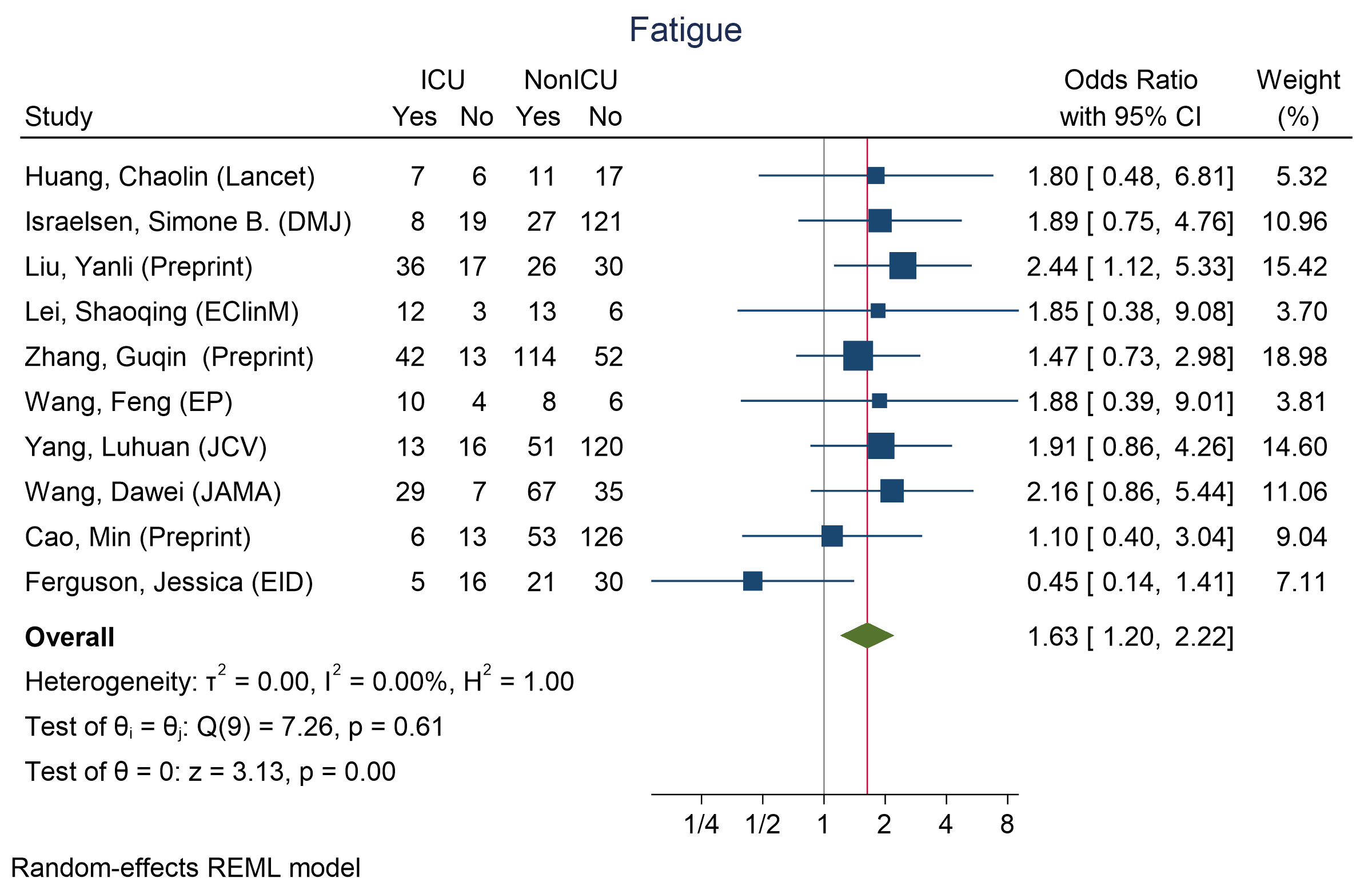

Figure 48 - Forest plot for Odds Ratio and 95% Confidence Interval (CI) of **Fatigue** for those who require ICU admission vs. non-ICU admission

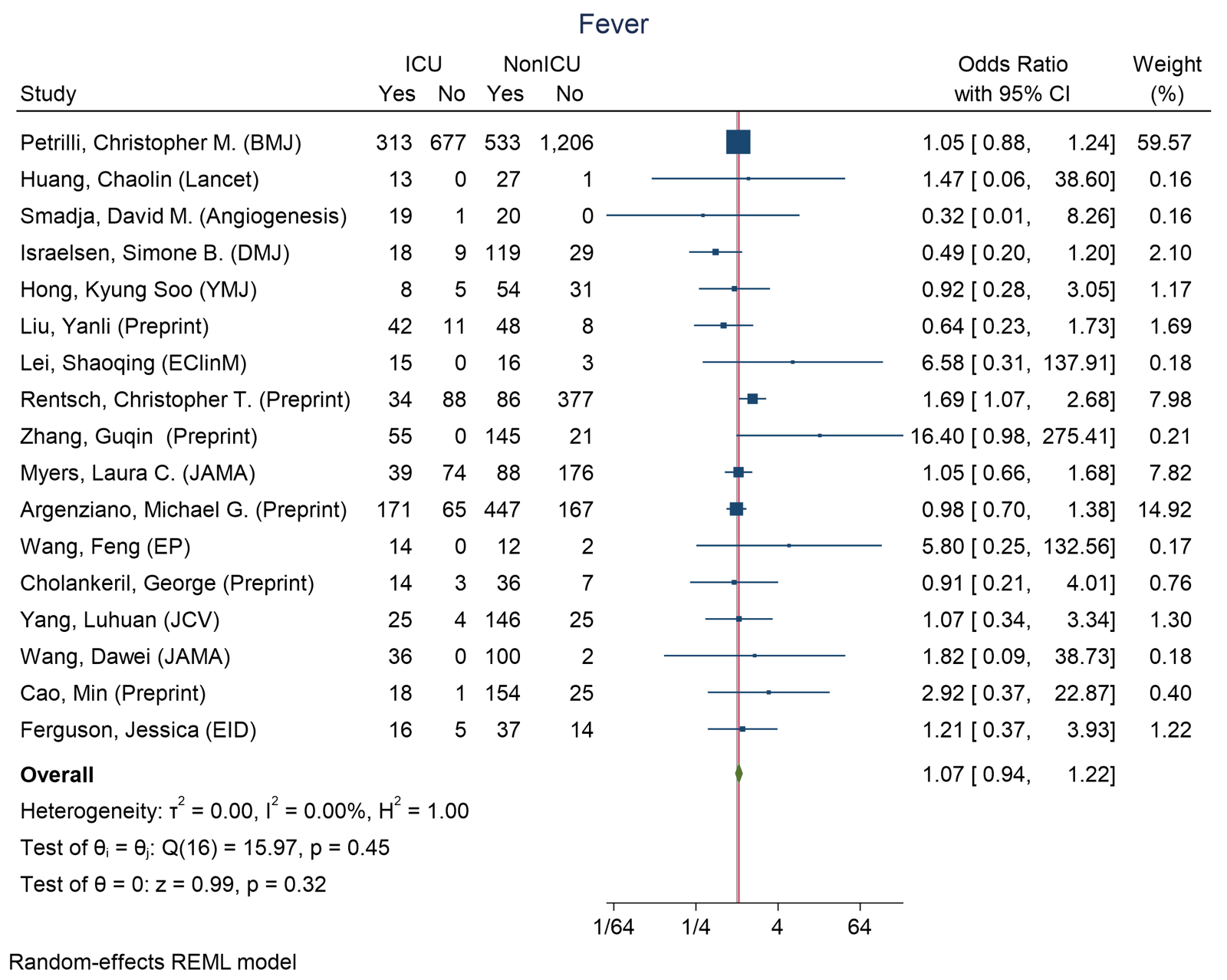

Figure 49 - Forest plot for Odds Ratio and 95% Confidence Interval (CI) of **Fever** for those who require ICU admission vs. non-ICU admission

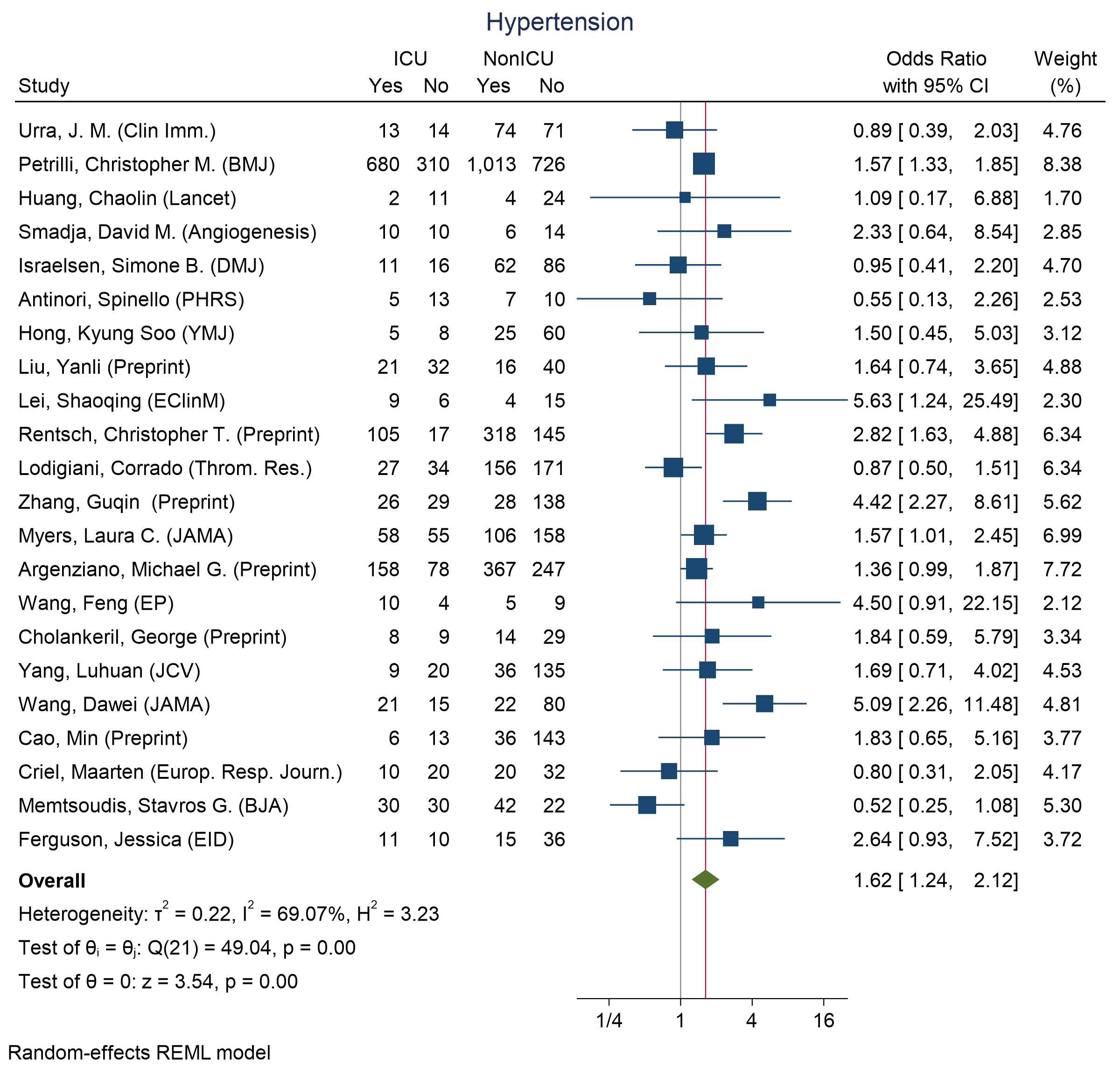

Figure 50 - Forest plot for Odds Ratio and 95% Confidence Interval (CI) of **Hypertension** for those who require ICU admission vs. non-ICU admission

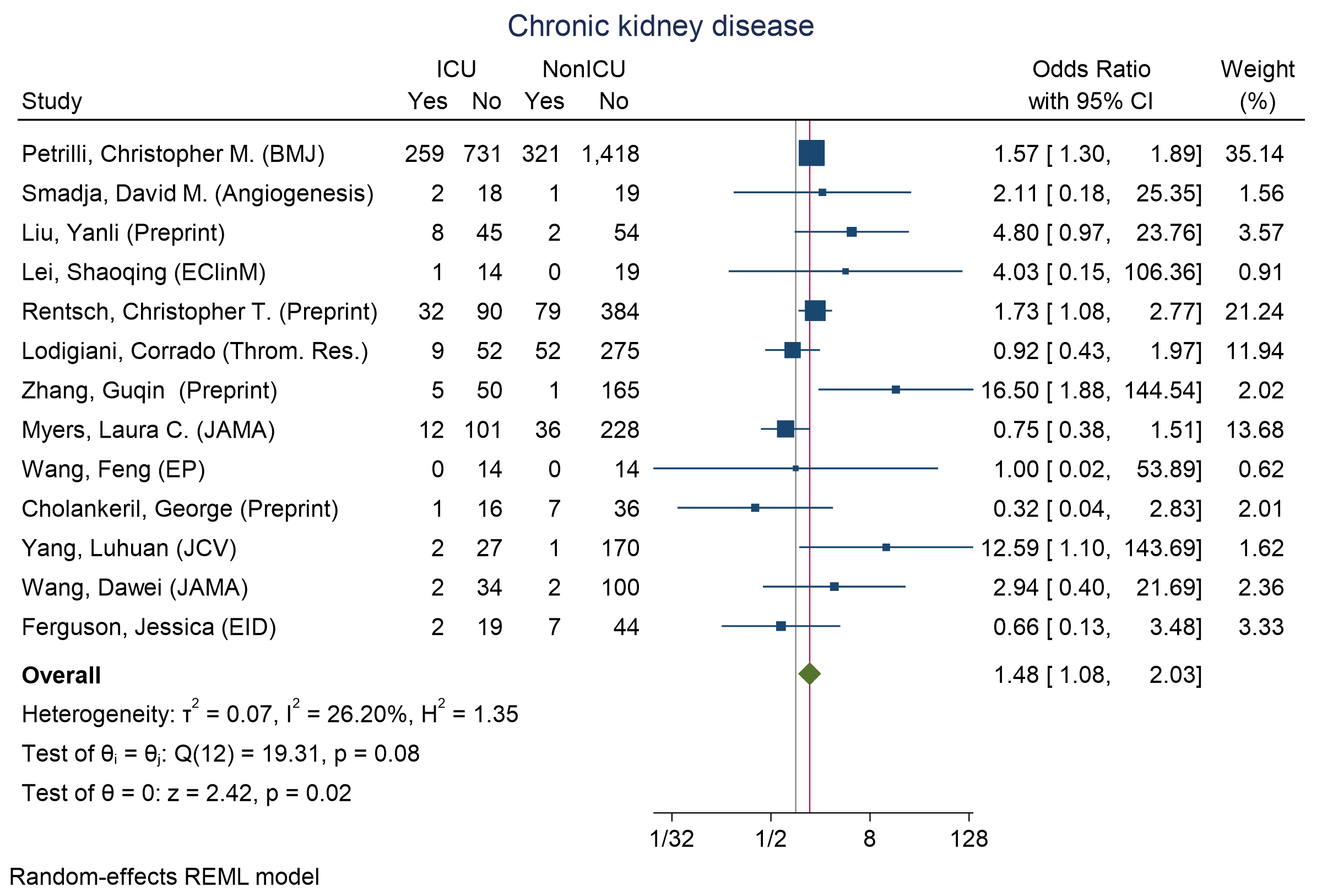

Figure 51 - Forest plot for Odds Ratio and 95% Confidence Interval (CI) of **chronic kidney disease** for those who require ICU admission vs. non-ICU admission

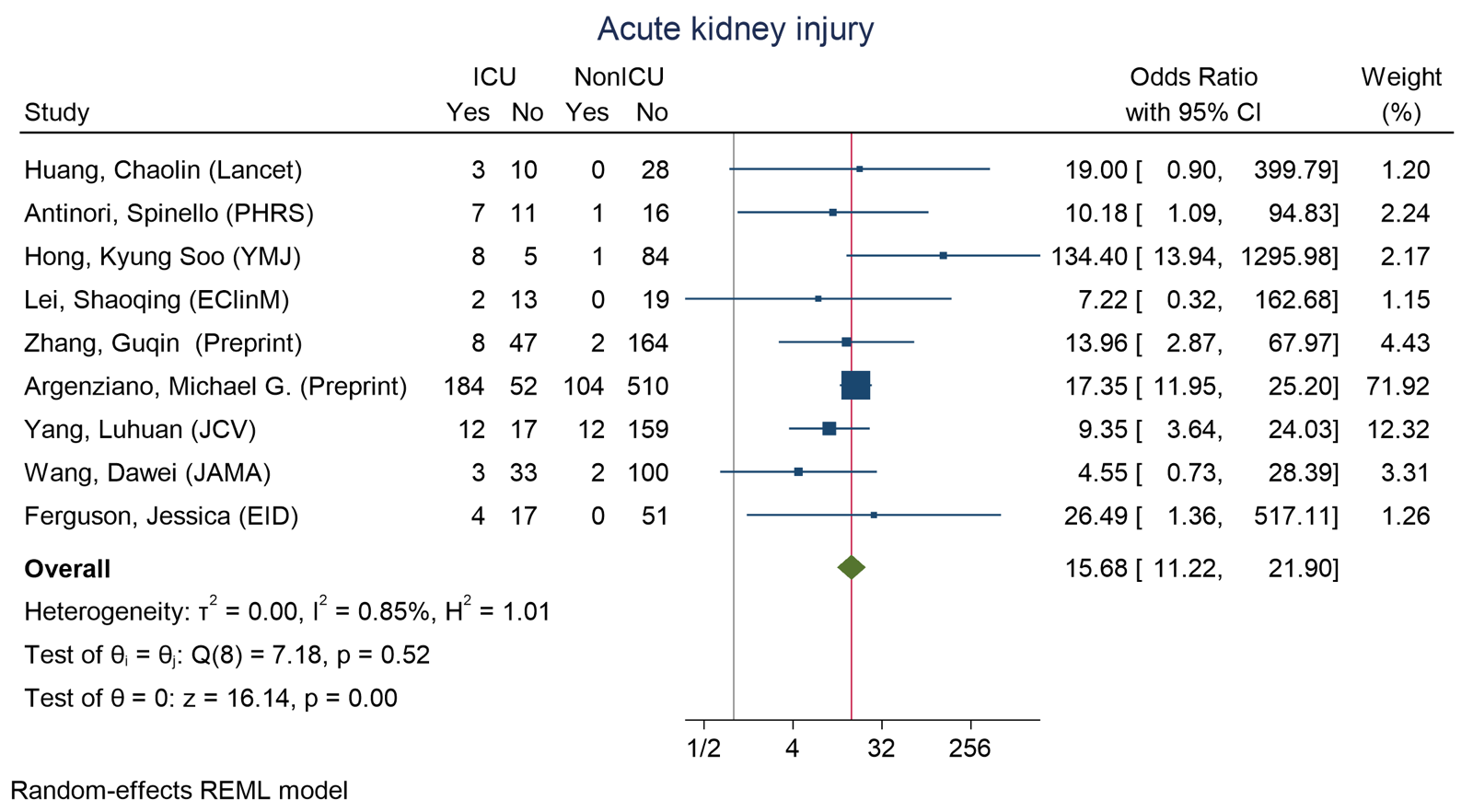

Figure 52 - Forest plot for Odds Ratio and 95% Confidence Interval (CI) of **acute kidney injury** for those who require ICU admission vs. non-ICU admission

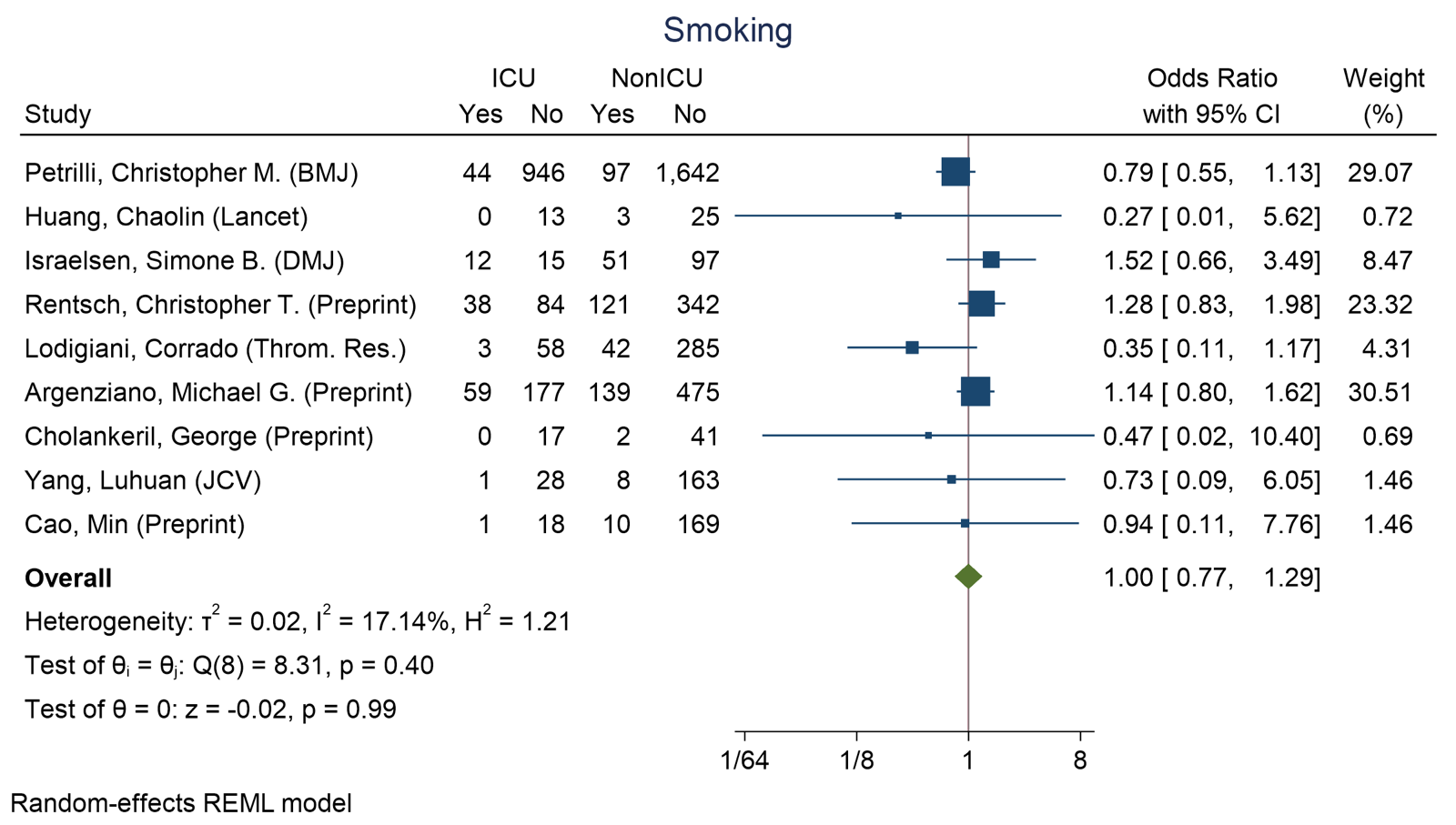

Smoking

Figure 53 - Forest plot for Odds Ratio and 95% Confidence Interval (CI) of **Smoking** for those who require ICU admission vs. non-ICU admission

### S7 Results for all available indicators in Intubated vs Non-Intubated analyses

| **Indicator** | **N. Studies** | **Pooled DoM** | **I^2^** |
| --- | --- | --- | --- |
| **Demographics** |  |  |  |
| Age (years) | 4 | 8.82 [3.06, 14.58] | 0 |
| **Laboratory Values** |  |  |  |
| Leukocyte (10^9^/L) | 4 | -1.04 [-5.78, 3.7] | 89.27 |
| Lymphocyte (10^9^/L) | 4 | -0.18 [-0.41, 0.05] | 63.75 |

Table 4 - Pooled difference of medians results for all indicators in Intubated vs Non-Intubated analyses

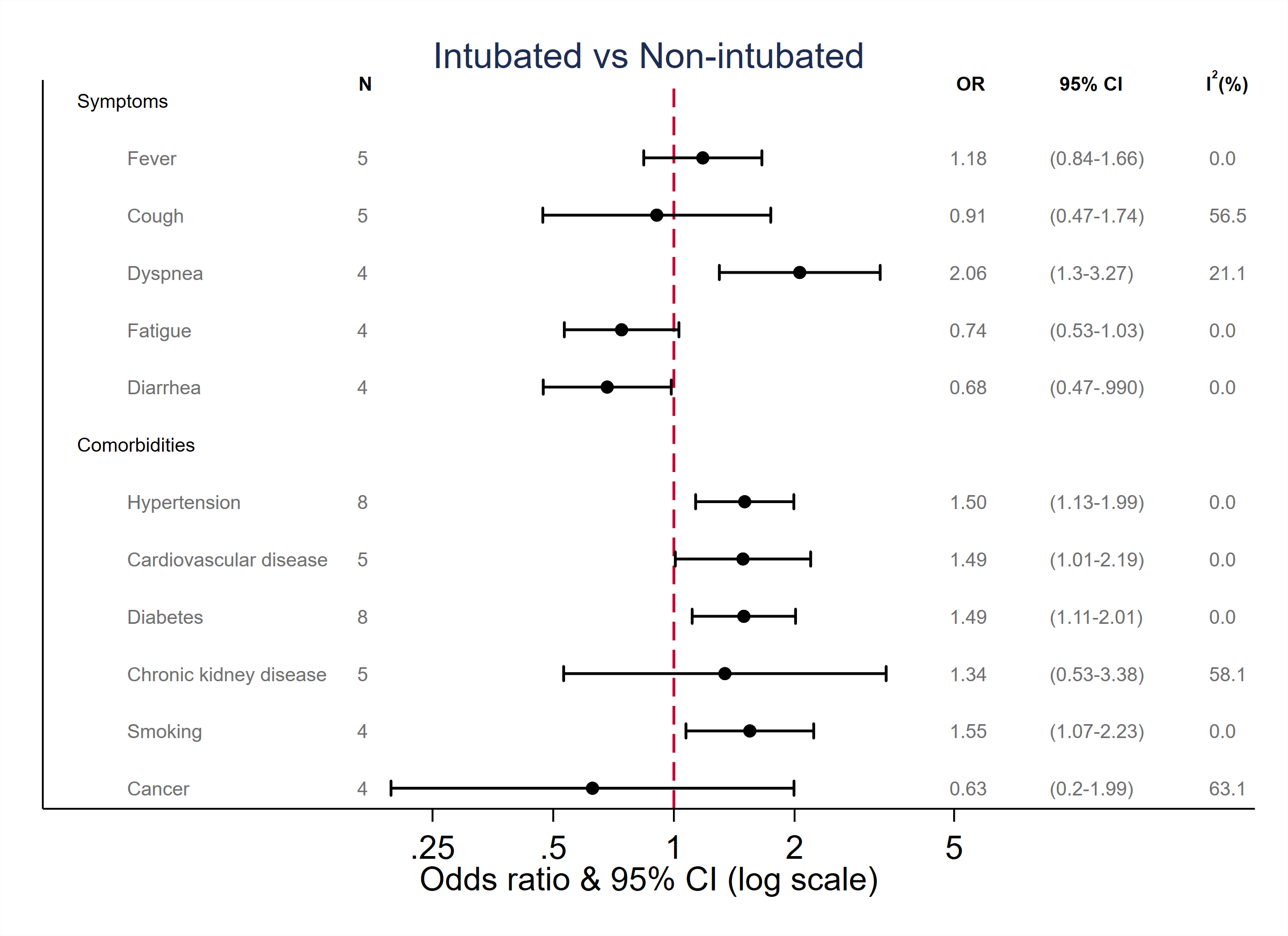

Figure 54 - Pooled Odds Ratios (OR) & 95% Confidence Interval (CI) for those who required intubation vs. non intubation

### S8 Results for all available indicators in hospitalization vs. non hospitalization

| **Indicator** | **N. Studies** | **Pooled DoM** | **I^2^** |
| --- | --- | --- | --- |
| **Demographics** |  |  |  |
| Age (years) | 10 | 13.16 [8.63, 17.68] | 94.74 |
| **Laboratory Values** |  |  |  |
| Leukocyte (10^9^/L) | 5 | -0.44 [-1.07, 0.19] | 39.44 |
| Lymphocyte (10^9^/L) | 4 | -0.40 [-0.62, -0.19] | 74.75 |
| Platelets (10^9^/L) | 4 | -28.68 [-39.61, -17.75] | 4.16 |
| Creatinine (µmol/L) | 4 | 7.67 [-2.98, 18.31] | 63.77 |

Table 5 - showing pooled difference of medians results for all biomarkers in Hospitalized vs Non-Hospitalized analyses

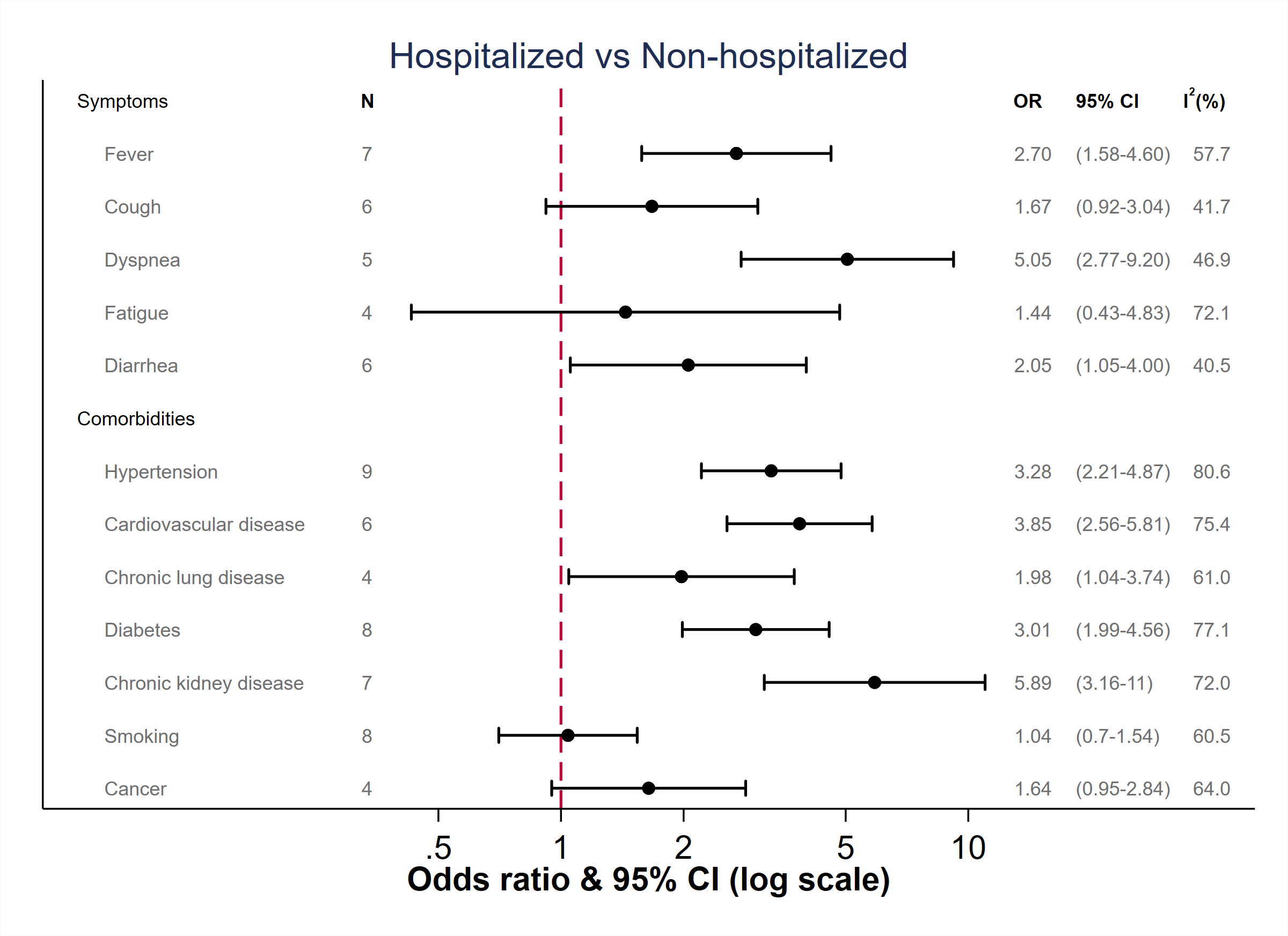

Figure 55 **-** Pooled Odds Ratios (OR) & 95% Confidence Interval (CI) of those who required hospitalization vs. non hospitalization

### S9 Sensitivity analyses

In the following sensitivity analyses, we considered excluding studies with outlier difference of medians values.

We considered removing the study of Yang, Xia (CCM) in the analysis of D-Dimer across those who died and those who survived. When including this study, we obtain a pooled difference of medians of 1.2914 [95% CI: 0.8964, 1.6865] and I^2^ = 81.53 (Fig. 11). When excluding this study, we obtain a pooled estimate of 1.2902 [95% CI: 0.8957, 1.6846] and I^2^ = 81.98.

We considered removing the studies of Liu, Yanli, Zhang, Guqin, and Wang, Dawei in the analysis of D-Dimer across those who required ICU and those who did not. When including this study, we obtain a pooled difference of medians of 0.3024 [95% CI: -0.2041, 0.8088] and I^2^ = 83.97 (Fig. 37). When excluding this study, we obtain a pooled estimate of 0.3123 [95% CI: -0.1472, 0.7718] and I^2^ = 84.21.

We considered removing the study of Xie, Jianfeng in the analysis of CRP across those who died and those who survived. When including this study, we obtain a pooled difference of medians of 69.1016 [95% CI: 50.4309, 87.7724] and I^2^ = 95.99 (Fig. 13). When excluding this study, we obtain a pooled estimate of of 59.2534 [95% CI: 41.5901, 76.9167] and I^2^ = 95.66.

### S10 Funnel plots for mortality vs. survival across different indicators

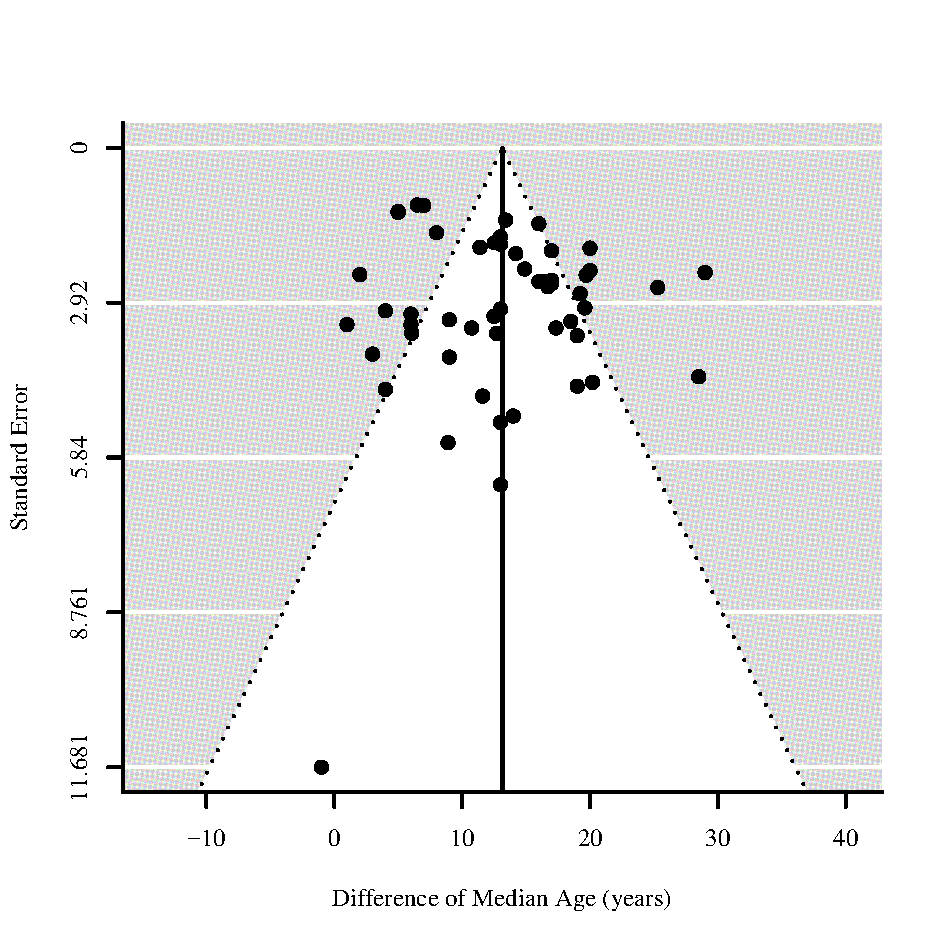

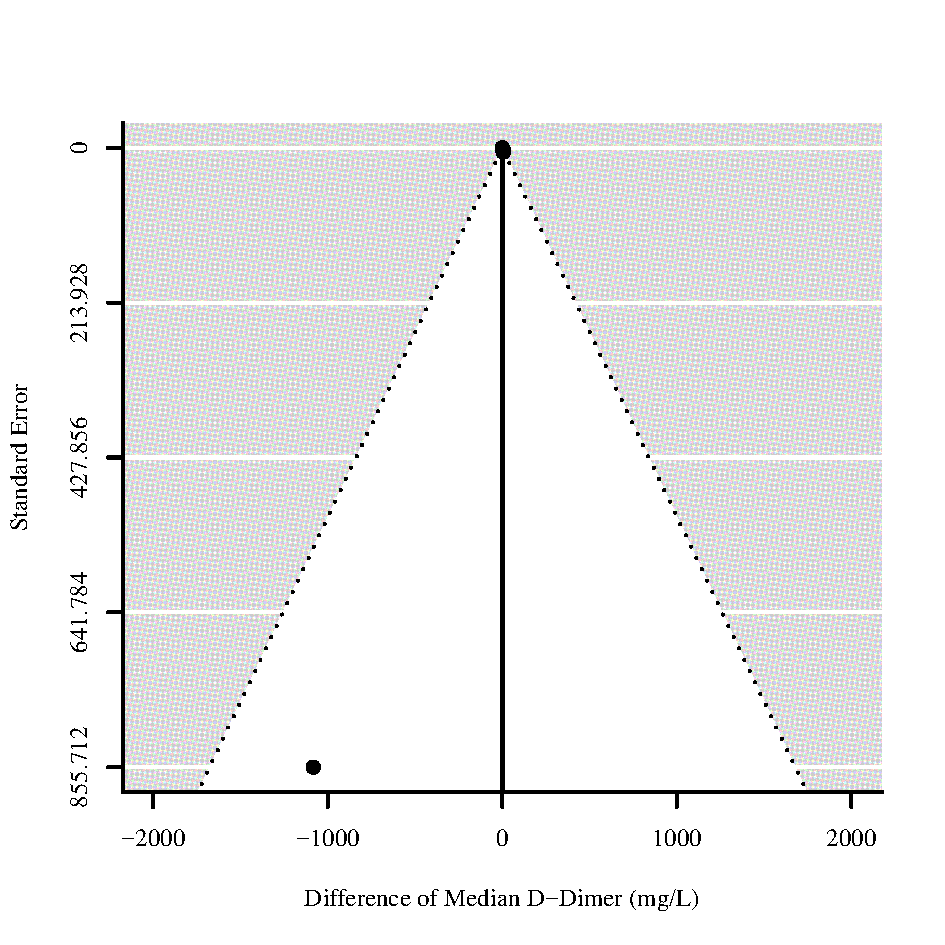

Figure 56 - Funnel plots for differences of medians of **Age** and **D-Dimer** in those who died vs. survived

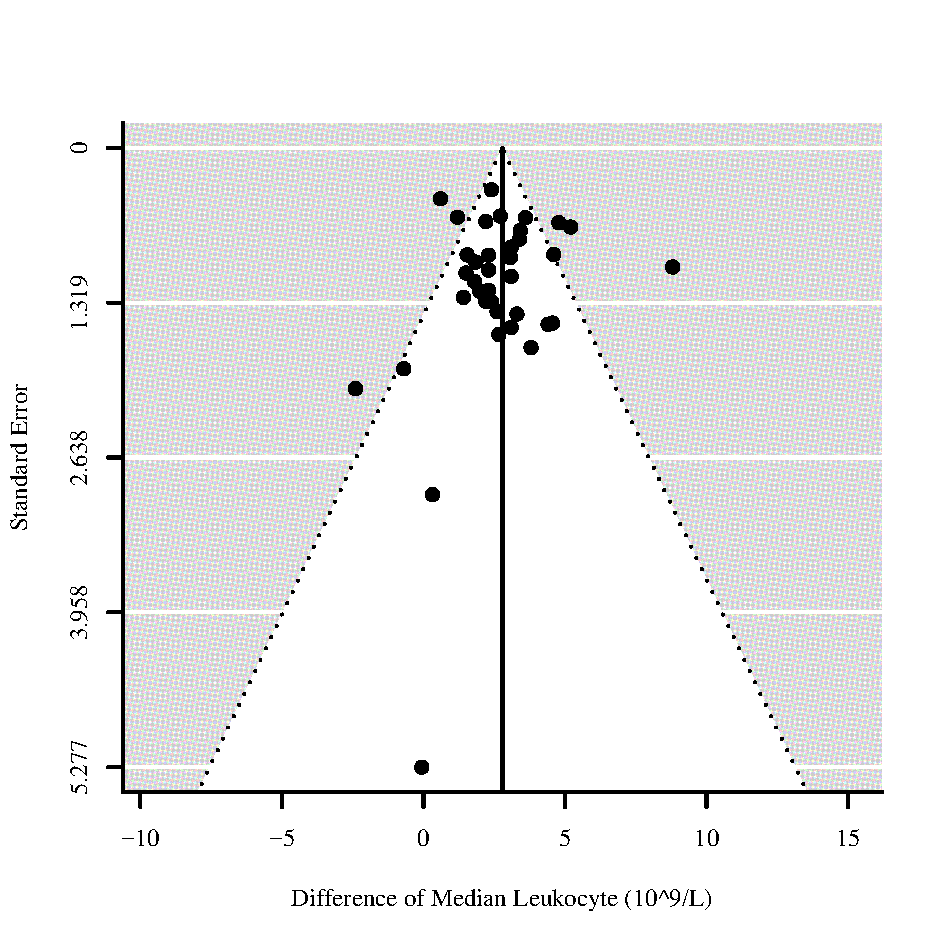

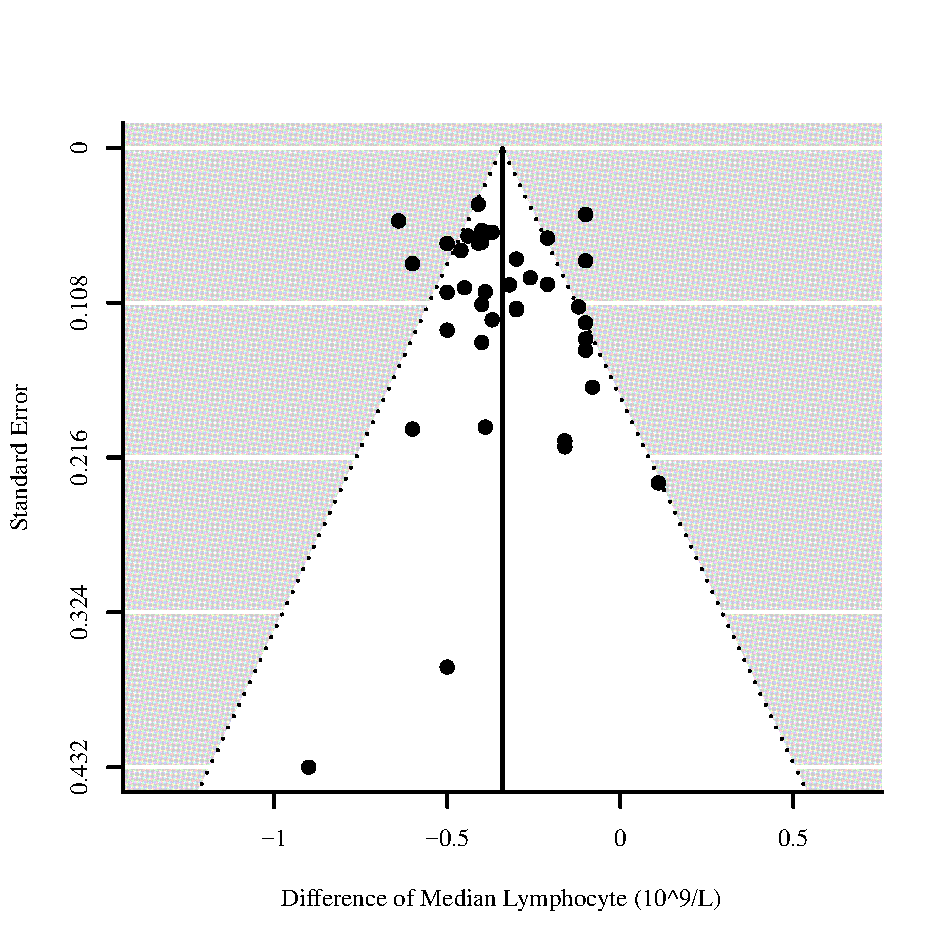

Figure 57 - Funnel plots for differences of medians of **Leukocyte** and **Lymphocyte** in those who died vs. survived

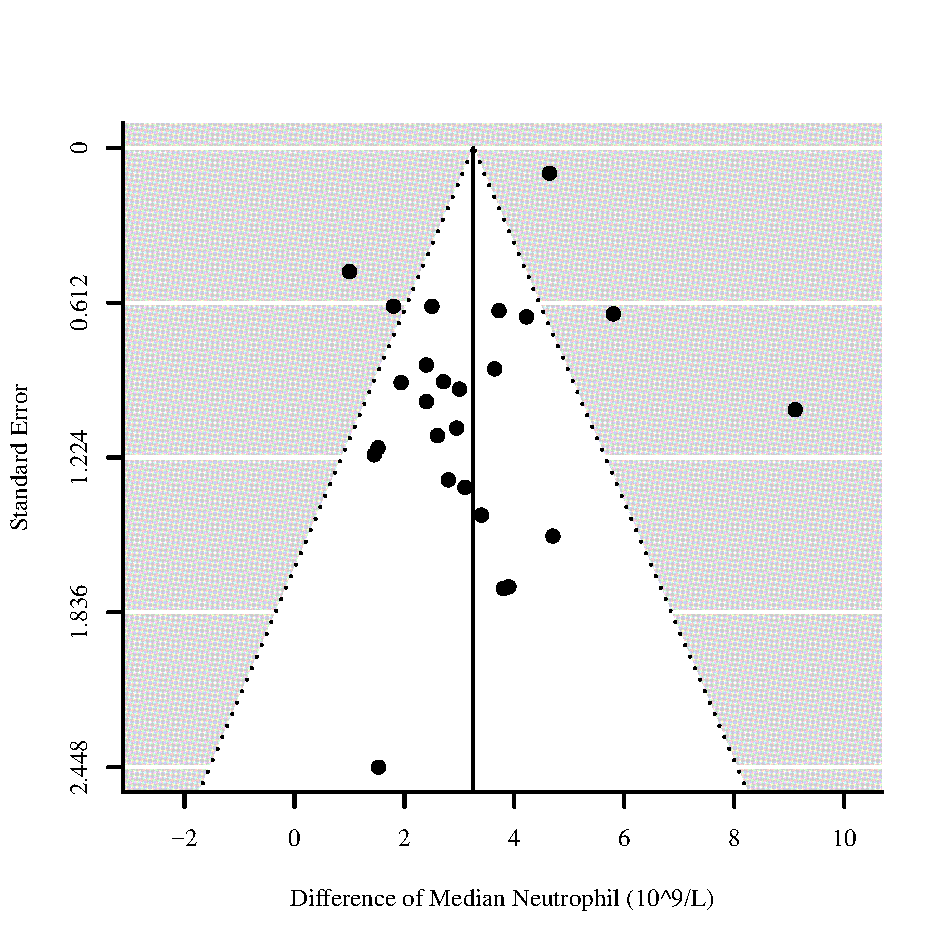

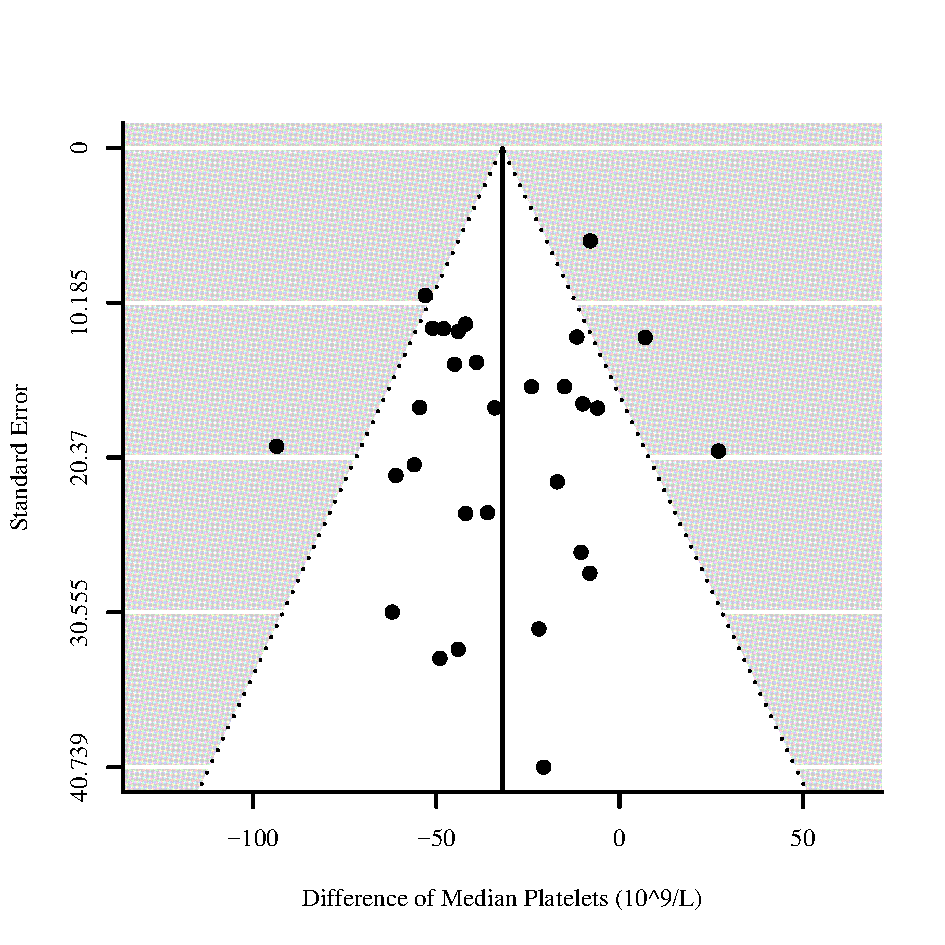

Figure 58 - Funnel plots for differences of medians of **Neutrophil** and **Platelets** in those who died vs. survived

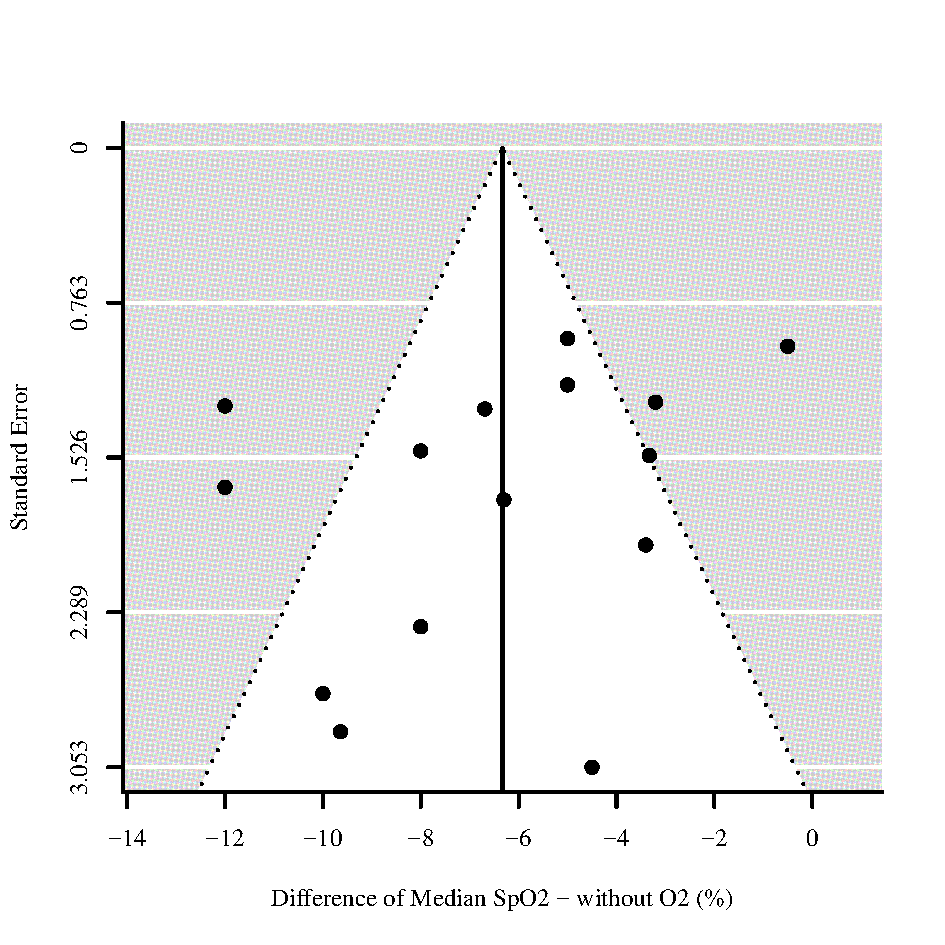

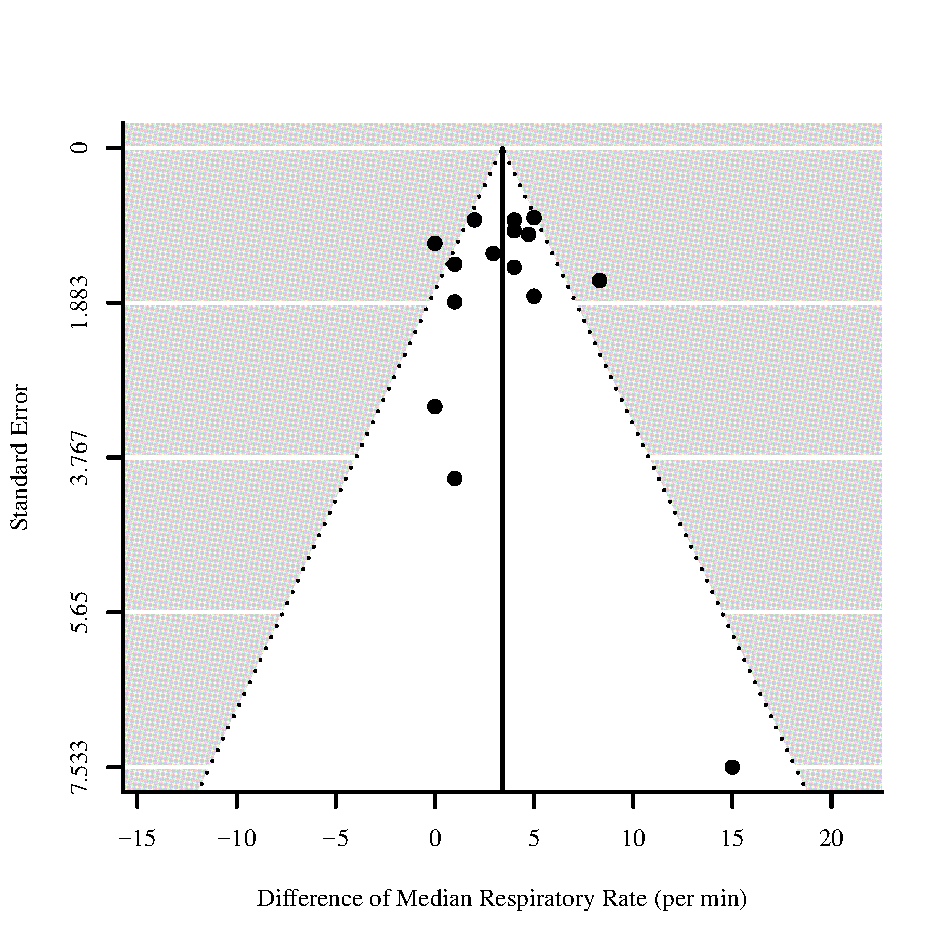

Figure 59 - Funnel plots for differences of medians of **Oxygen Saturation (SpO2)** without oxygen (O2) and **Respiratory Rate** in those who died vs. survived

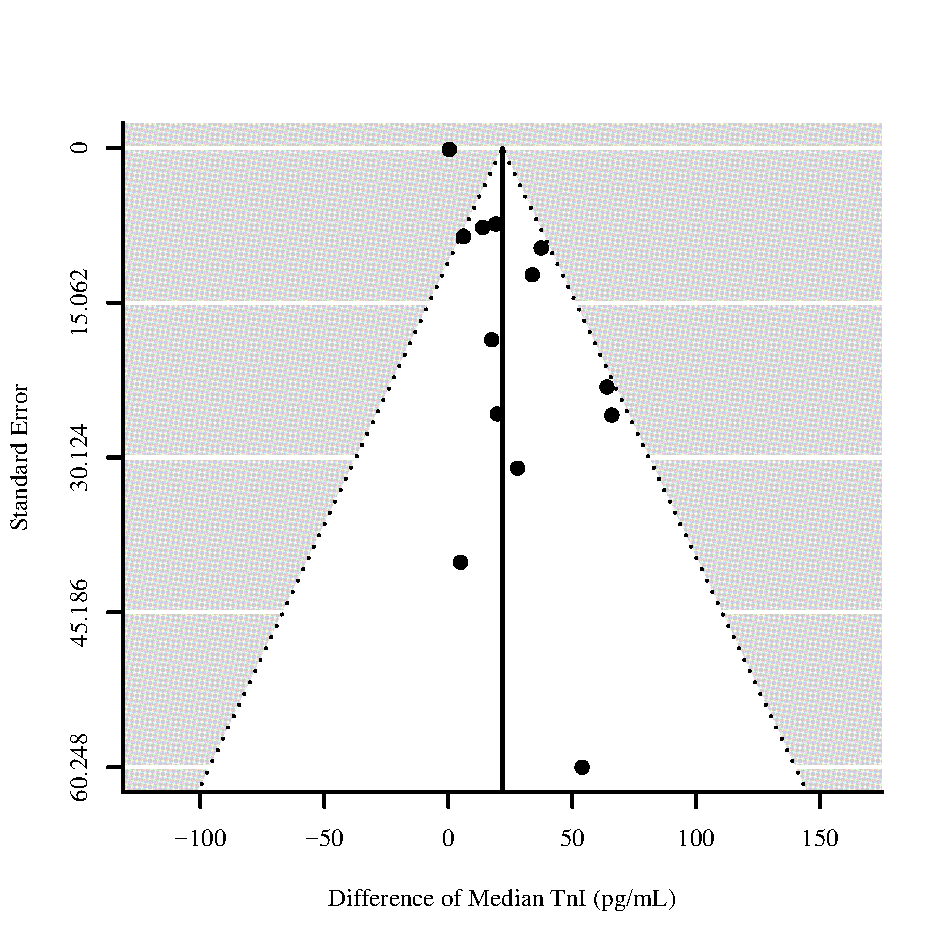

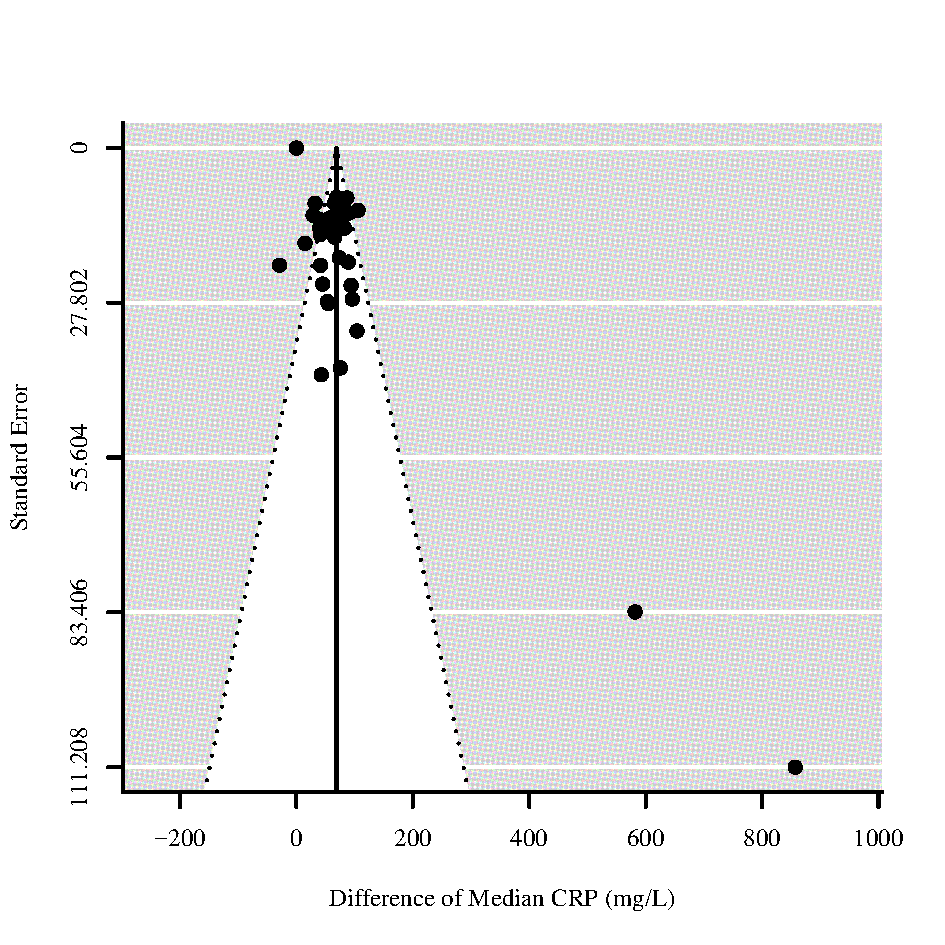

Figure 60 - Funnel plots for differences of medians of **Troponin I (TnI)** and **C-reactive protein (CRP)** in those who died vs. survived

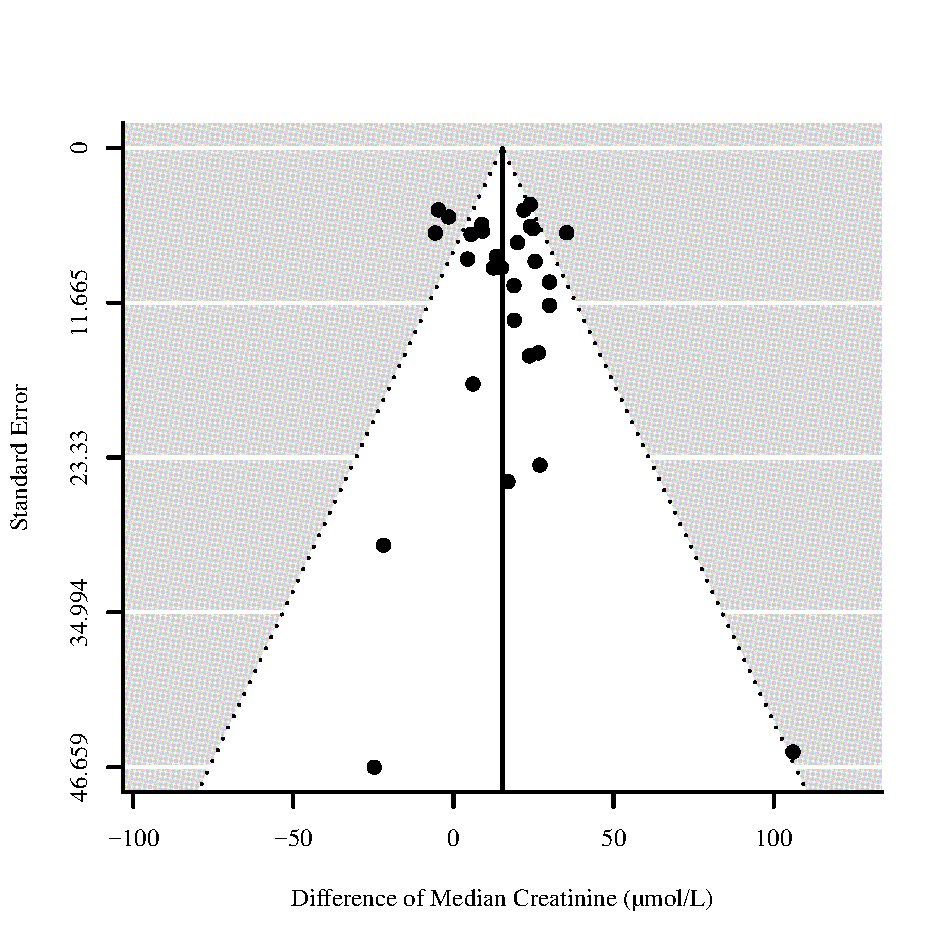

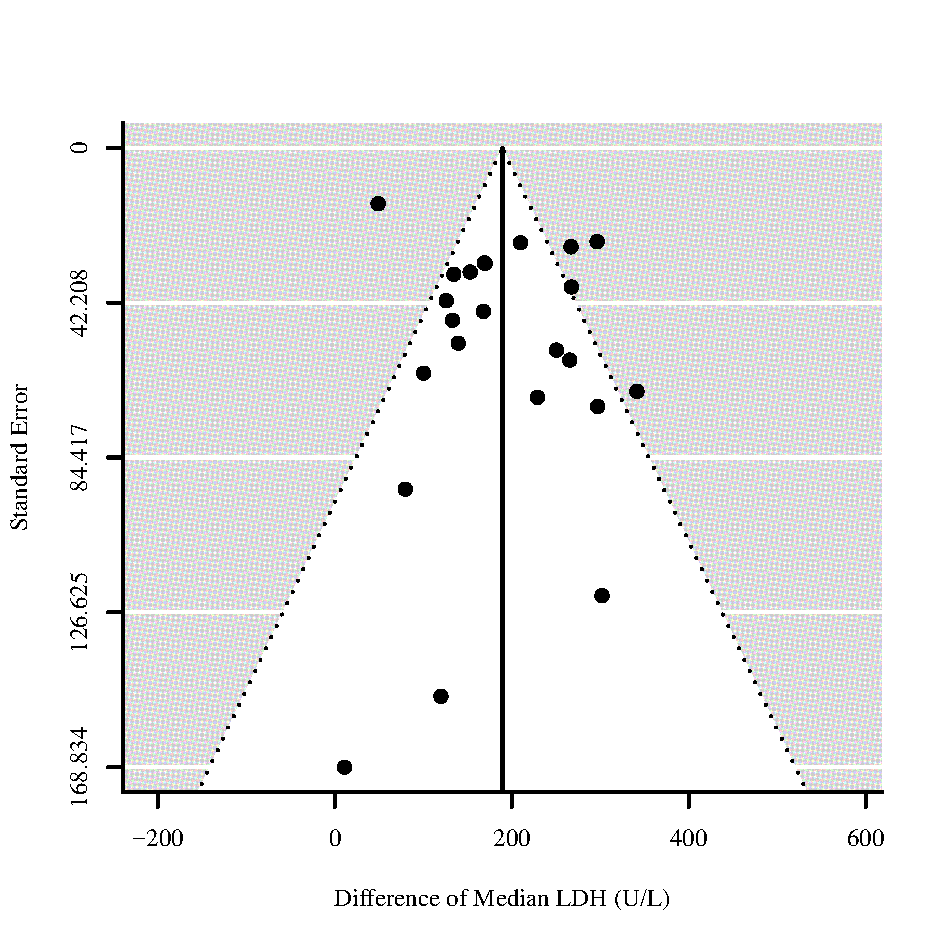

Figure 61 – Funnel plots for differences of medians of **Blood Urea Nitrogen (BUN)** and **Lactate Dehydrogenase (LDH)** in those who died vs. survived

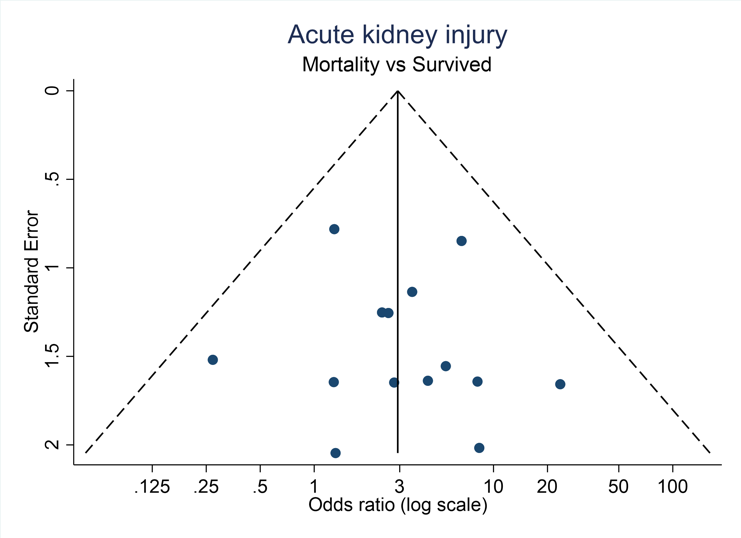

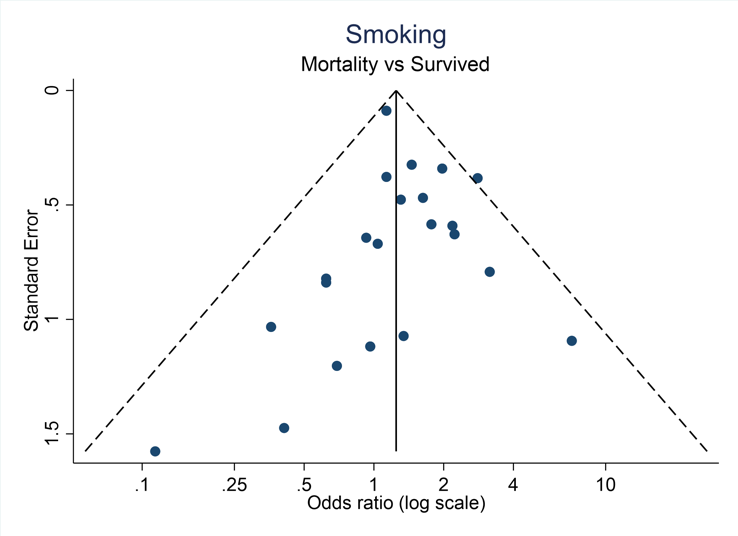

Figure 62 - Funnel plots for Odds ratios of **Acute kidney injury** and **Smoking** in those who died vs. survived

### S11 Funnel plots for ICU admission vs. non-ICU admission across different indicators

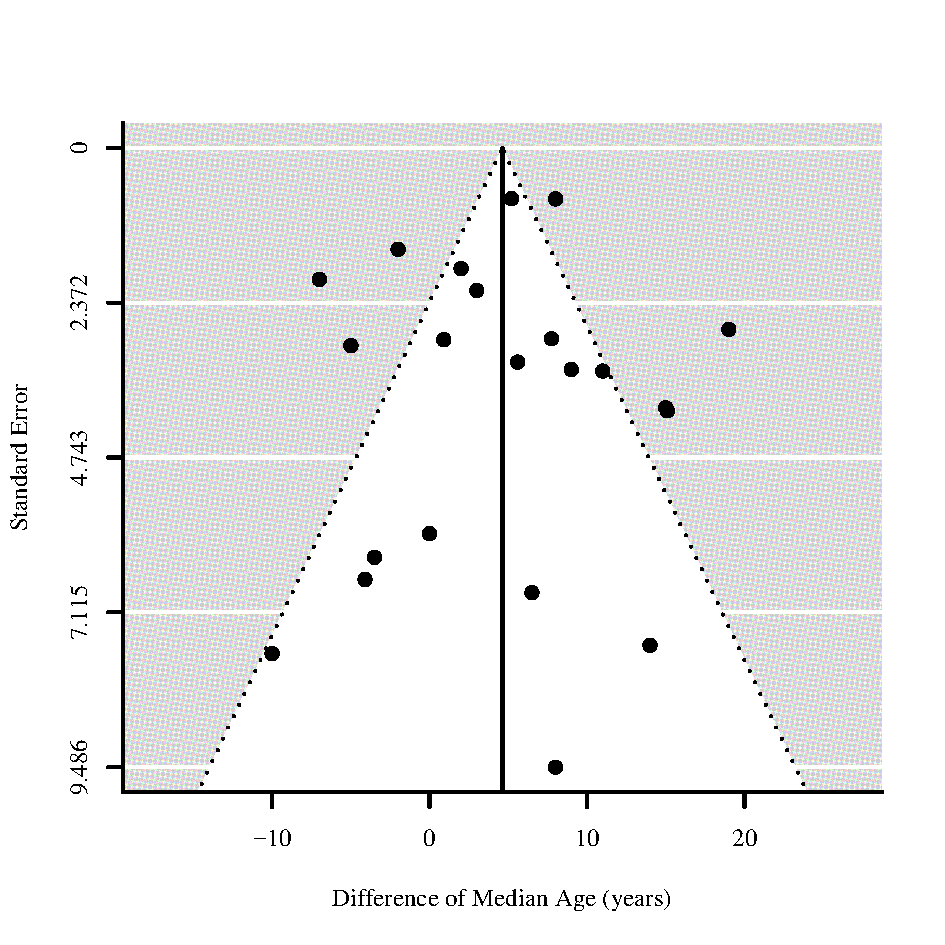

Figure 63 - Funnel plot for difference of medians of **Age** for those who require ICU admission vs. non-ICU admission

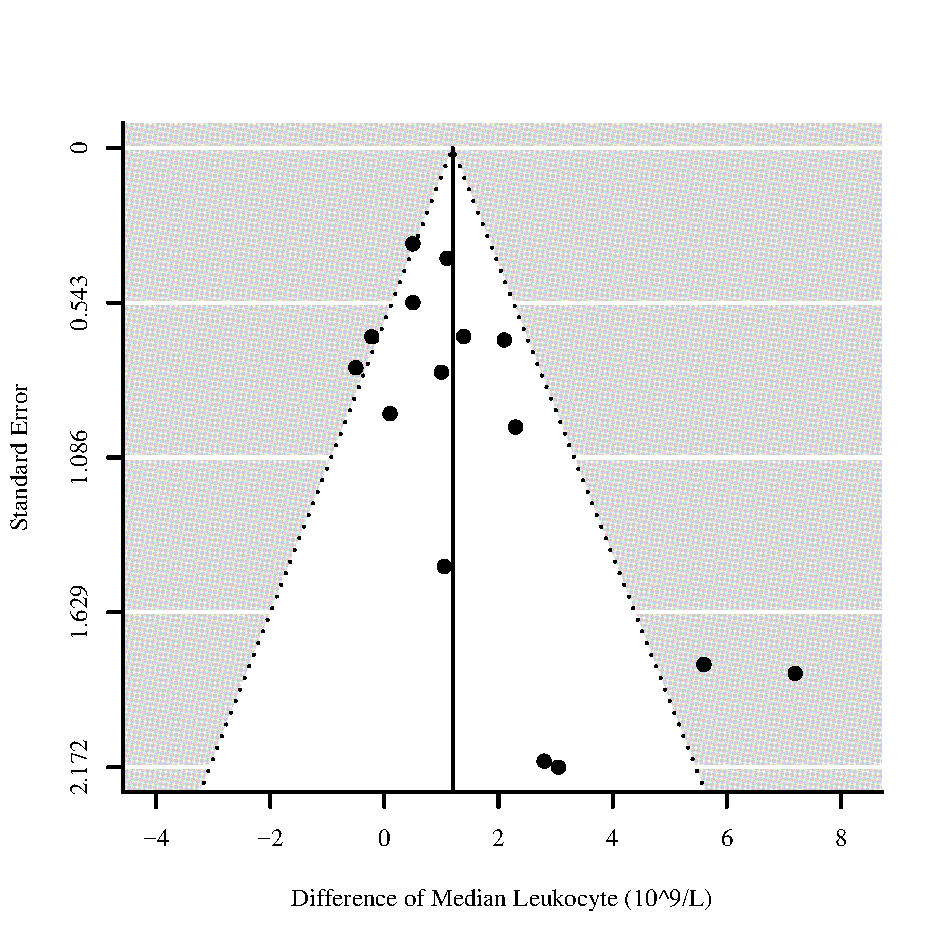

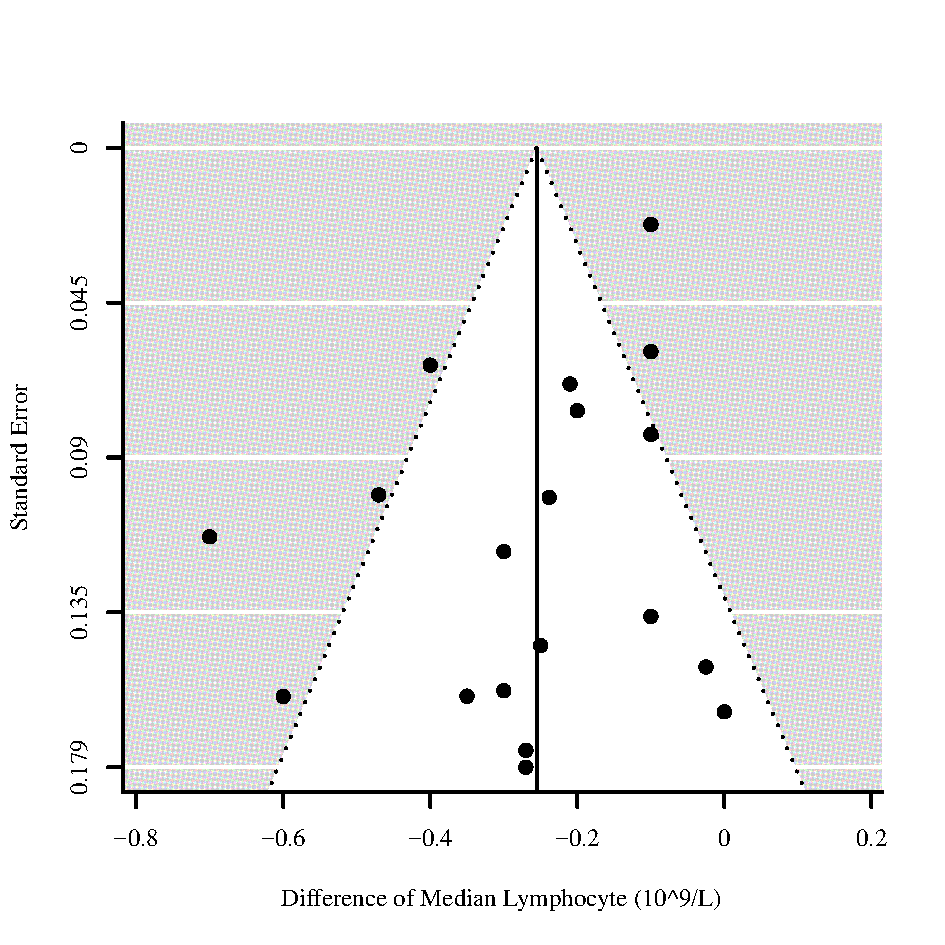

Figure 64 - Funnel plots for differences of medians of **Leukocyte** and **Lymphocyte** for those who require ICU admission vs. non-ICU admission

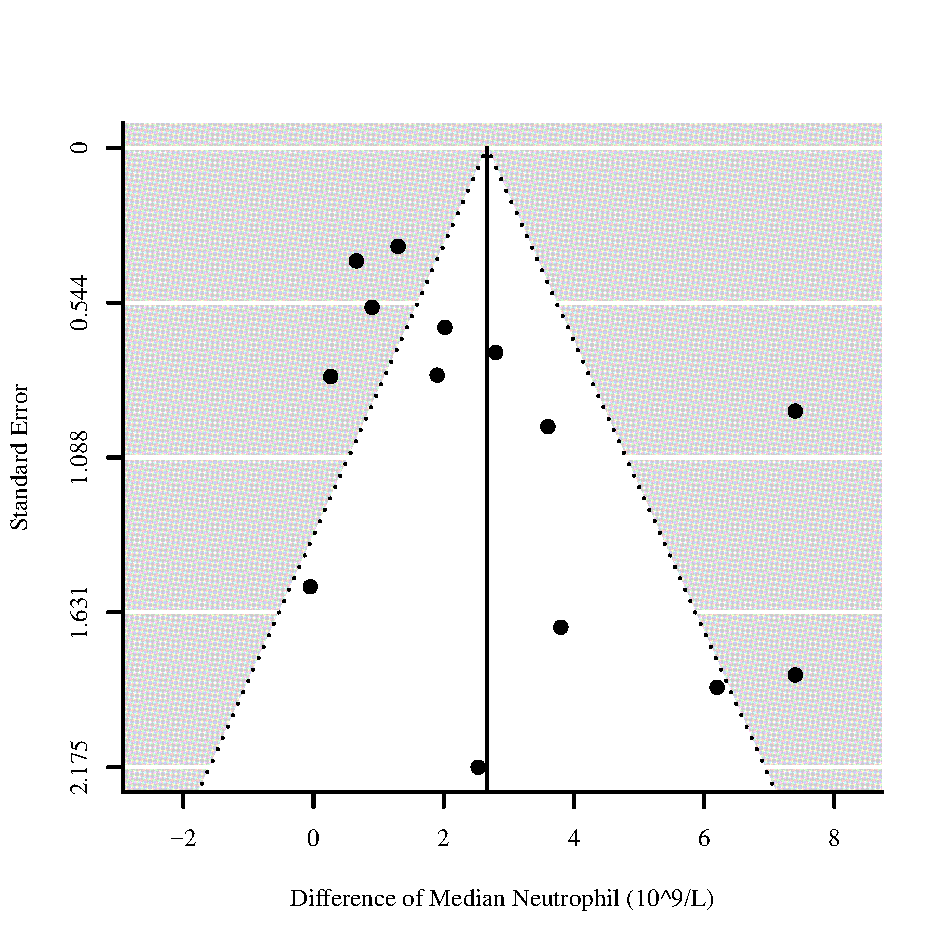

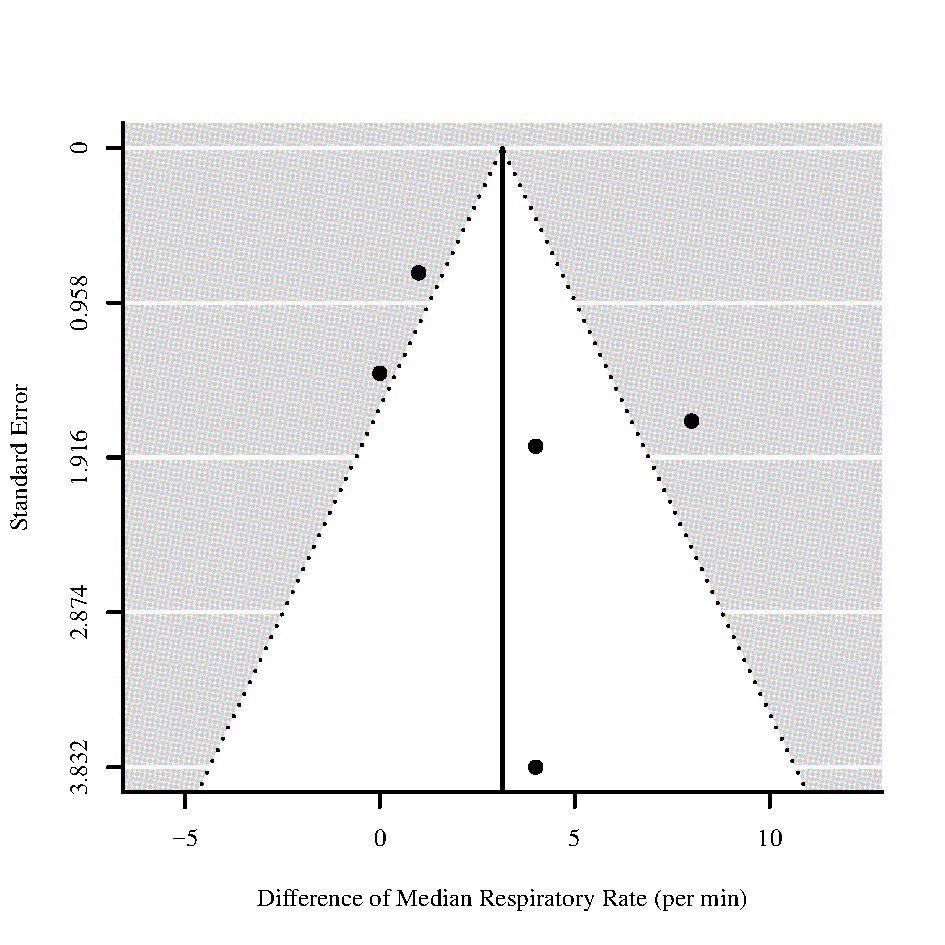

Figure 65 - Funnel plots for differences of medians of **Neutrophil** and **Respiratory Rate** for those who require ICU admission vs. non-ICU admission

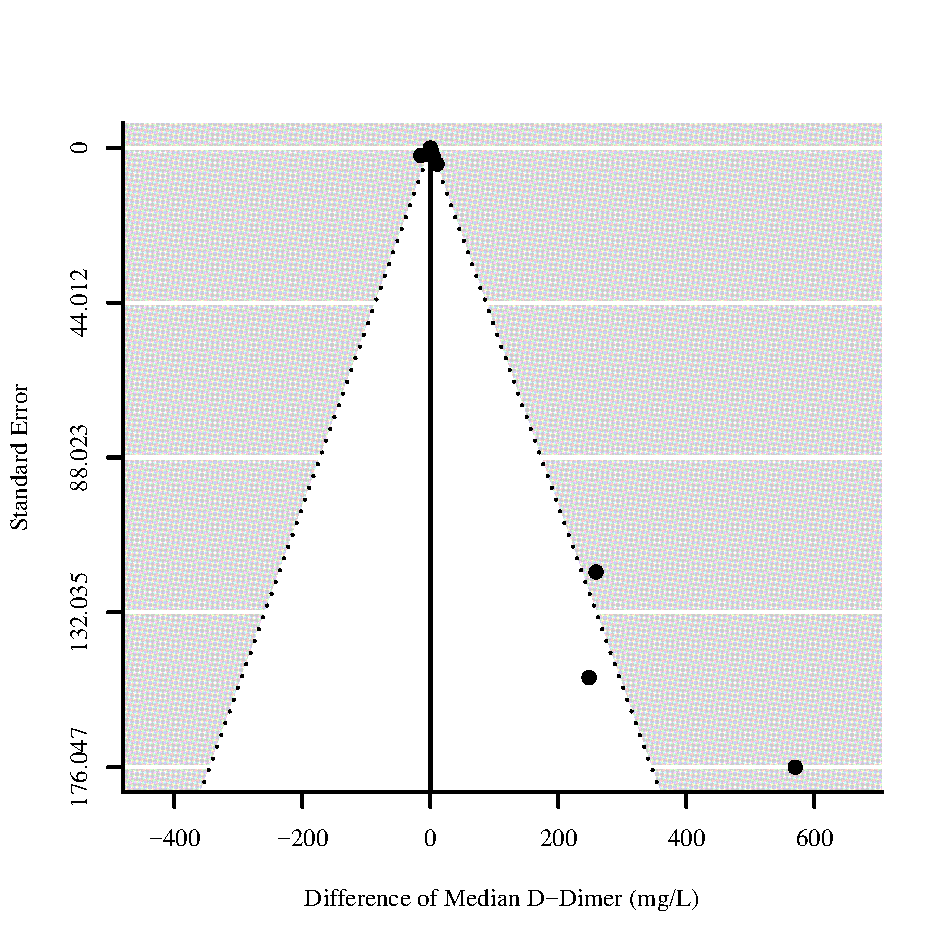

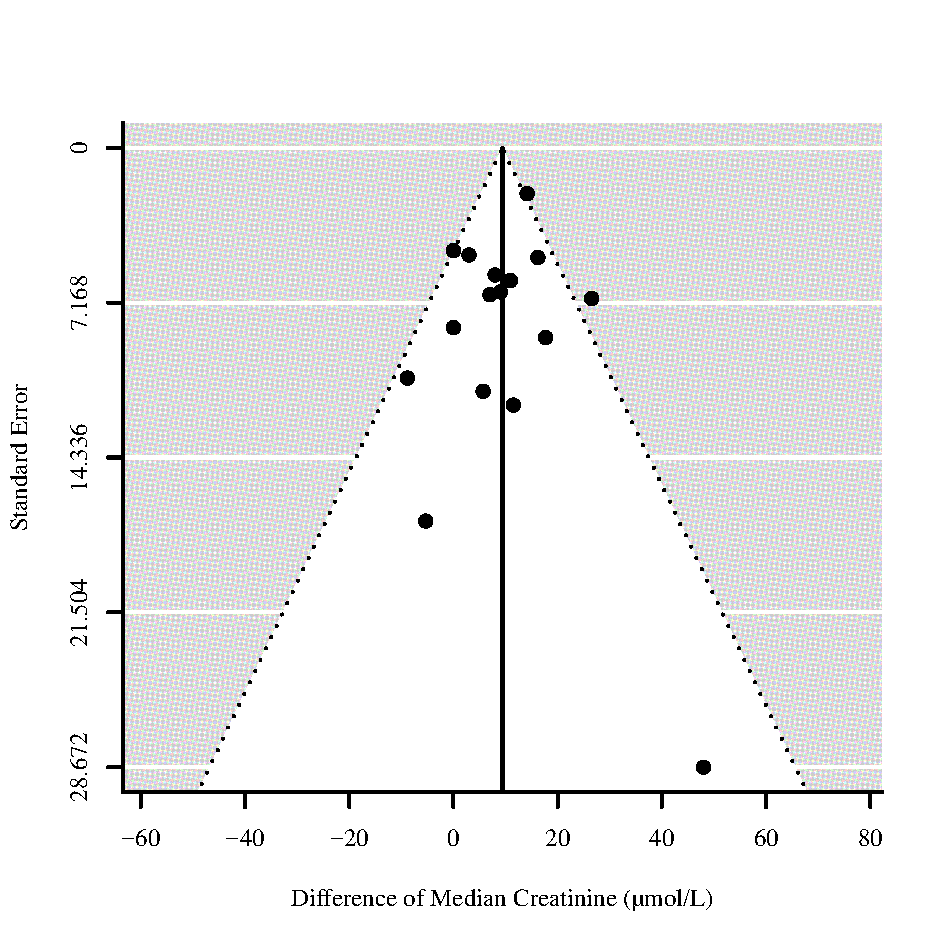

Figure 66 - Funnel plots for differences of medians of **D-Dimer** and **Creatinine** for those who require ICU admission vs. non-ICU admission

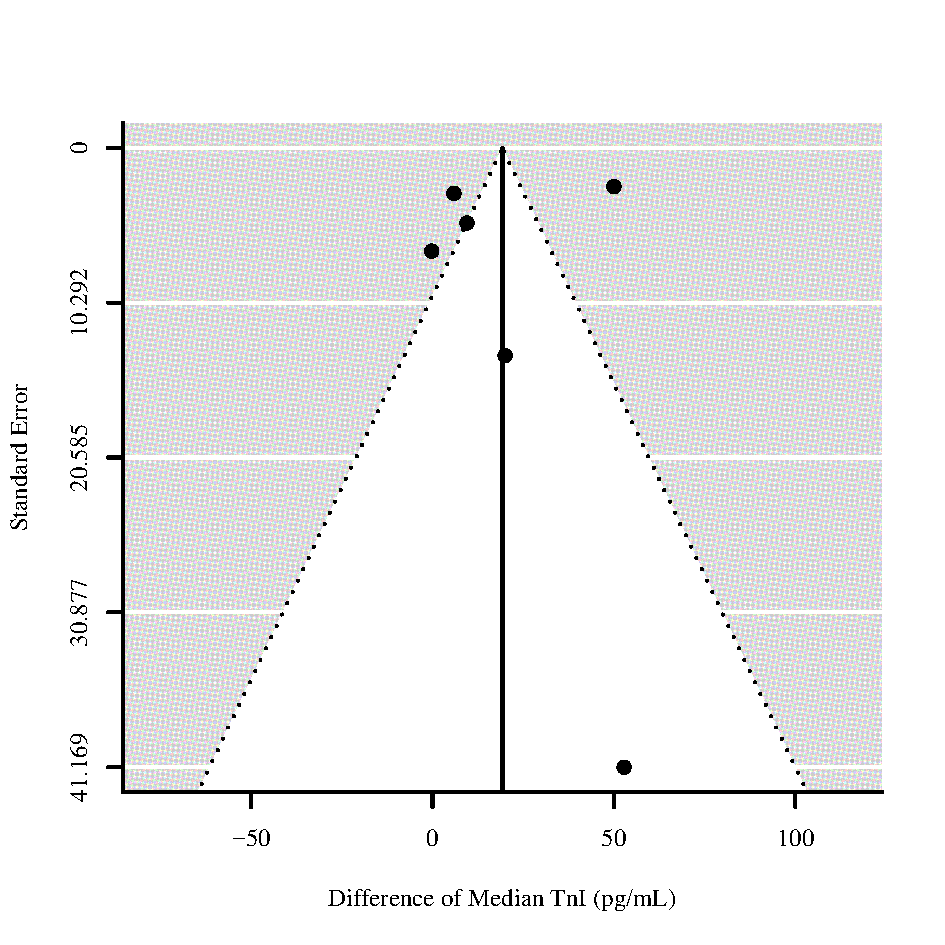

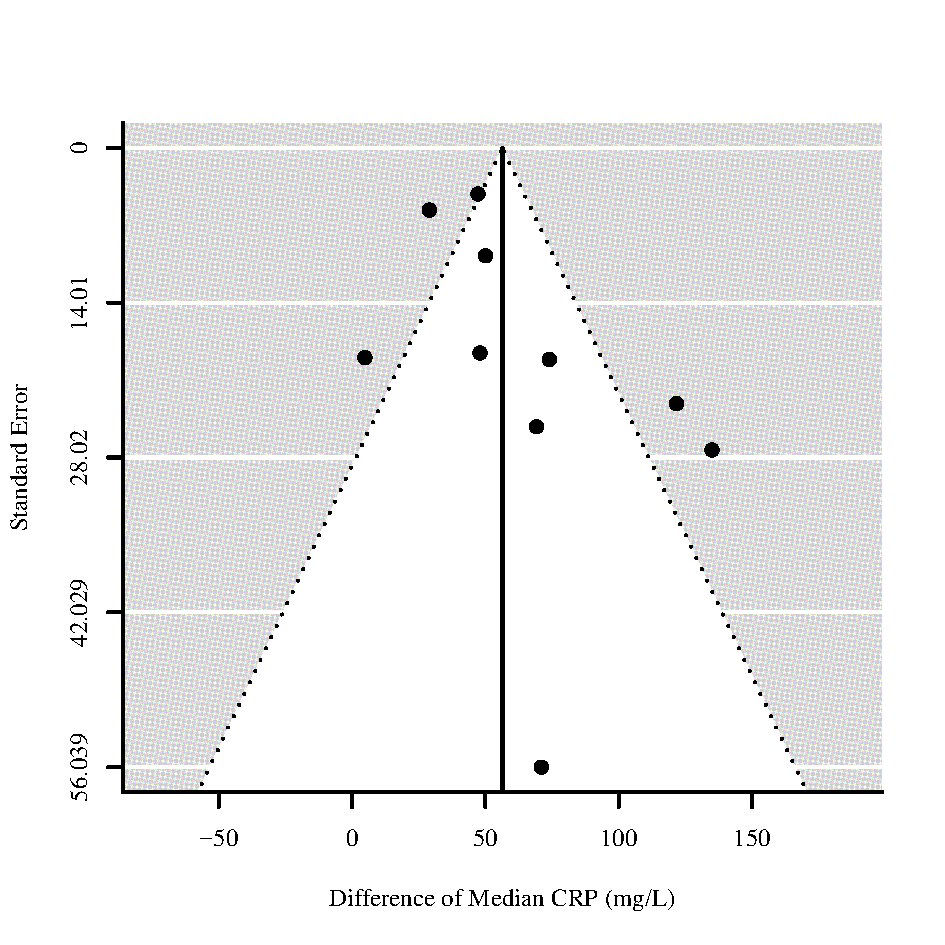

Figure 67 - Funnel plots for differences of medians of **Troponin I (TnI)** and **C-reactive protein (CRP)** for those who require ICU admission vs. non-ICU admission

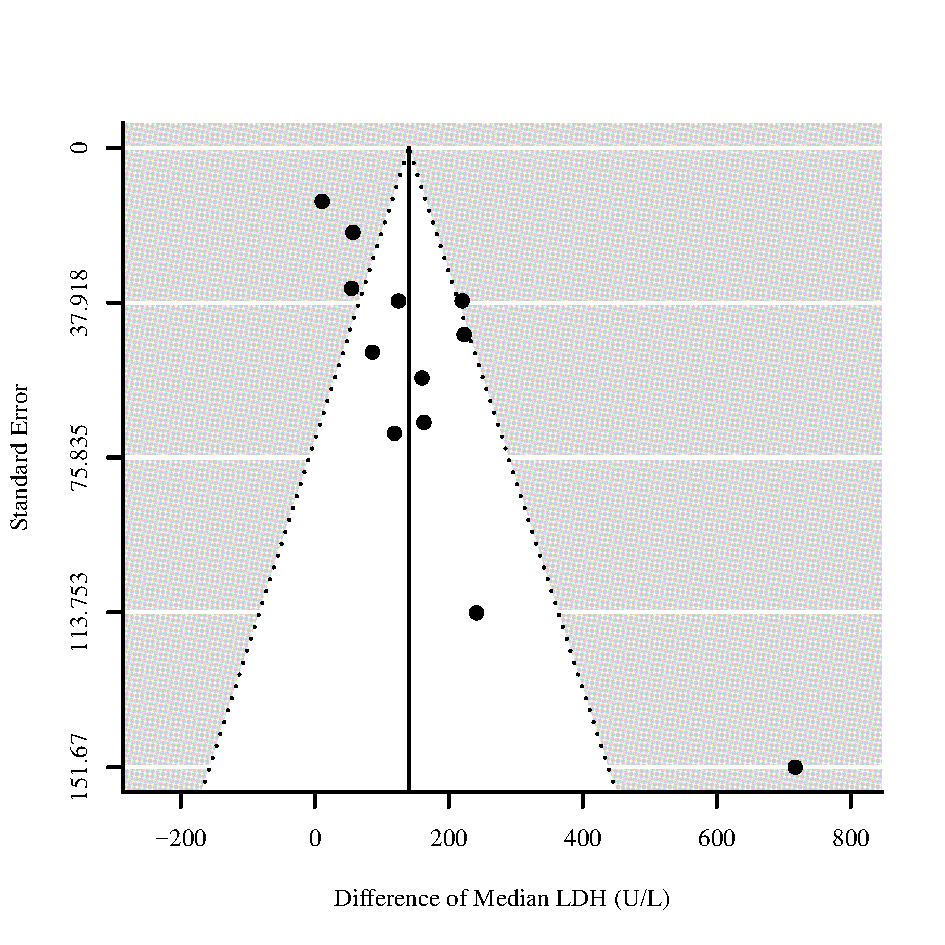

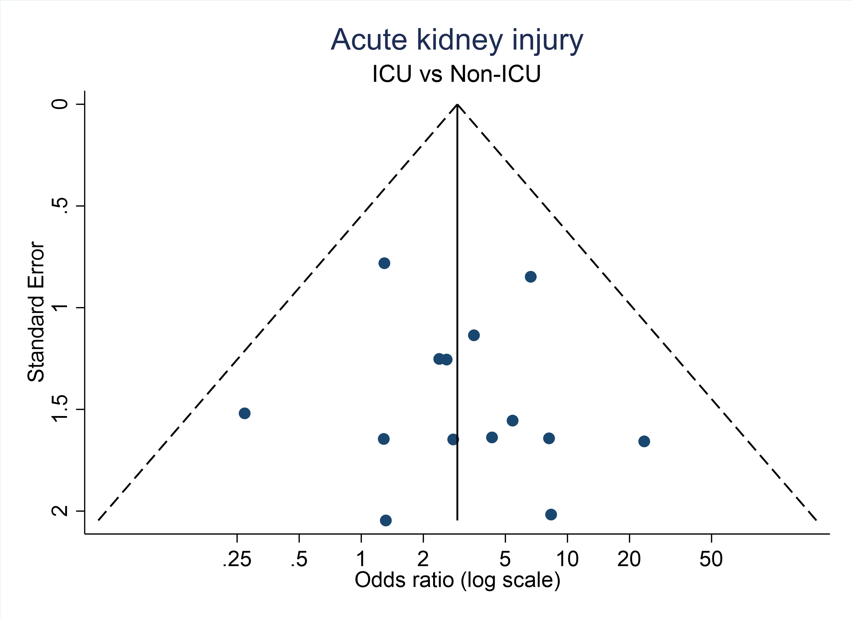

Figure 68 - Funnel plots for difference of medians of **Lactate Dehydrogenase (LDH)** and Odds ratio of **Acute kidney injury** for those who require ICU admission vs. non-ICU admission

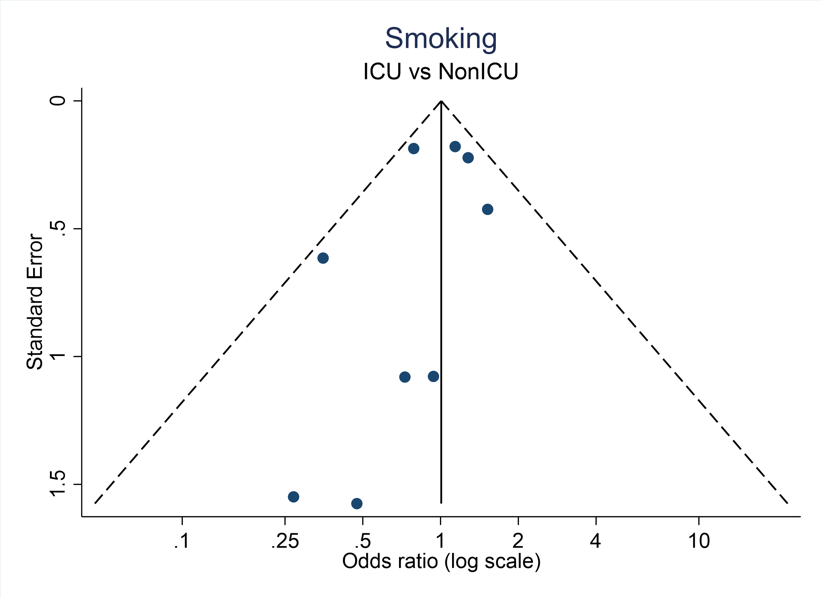

Figure 69 - Funnel plot for Odds of **Smoking** for those who require ICU admission vs. non-ICU admission

### S12 List of data items extracted from studies

**Demographics**

Age

Gender

**Laboratory values**

Platelets

Lymphocytes

Neutrophils

Hemoglobin

Hematocrit

Leukocytes

C-reactive protein

Procalcitonin

Interleukin-6

CD-4

Viral load

Creatinine

Blood Urea Nitrogen

Glomerular filtration rate

Lactate dehydrogenase

Albumine

Creatine Kinase

Creatine kinase – myocardial band

Troponin I

Troponin T

Brain natriuretic peptide

D-Dimer

Prothrombin

Activated partial thrombin time

Internationalized normalized ratio

Fibrinogen

Antithrombin activity

**Clinical**

Oxygen saturation without oxygen

Oxygen saturation with oxygen

Respiratory rate

CURB-65

SOFA

qSOFA

APACHE II

**Treatment**

Antiviral treatment

TNF-alpha

Non-invasive ventilation

Oxygen supplementation

ECMO

Renal replacement therapy

**Co-Morbidities**

Hypertension

Cardiomyopathy

Cardiovascular disease

Cerebrovascular disease

Previous Pneumonia

COPD

Asthma

Any chronic lung disease

Diabetes

Acute kidney injury

Smoker

Cancer

Chronic liver disease

Digestive system disease

Immunodeficiency

Tuberculosis

**Existing medication**

ACE Inhibitors

ARBS

Ibuprofen

ASS

DAPT

Diuretic

OAK

**Symptoms**

Fever

Cough

Dyspnea / Shortness of breath

Headache

Abdominal pain

Fatigue

Confusion

Pharyngalgia

Anosmia

Asymptomatic
